# How Many Time-Critical Emergencies? A Population-Based National Estimate of Attended Events per 100,000 US Residents

**DOI:** 10.64898/2026.11.09.26367629

**Authors:** Daniel Haight, Kelly A. Nisbet, Sunny Li

## Abstract

**Objectives:** To estimate how many time-critical emergencies fire and emergency medical services (EMS) crews attend each year per 100,000 US residents. We counted events in which a crew reached a living patient or a trapped person whose outcome plausibly depends on how soon effective treatment starts.

**Methods:** For 30 medical conditions, we took annual counts from US registries and datasets, or multiplied a national incidence count by the share of patients EMS attended. A condition was Tier A when a published study linked a measured delay, from minutes to a few hours, to death or major disability. Conditions whose time sensitivity rests on guidelines or expert consensus were Tier B. Fire and rescue events came from twelve months of National Emergency Response Information System public data, scaled to national totals. We removed double counting between conditions.

**Results:** Crews attended an estimated 589 time-critical events per 100,000 residents a year (494 to 697 with every condition at its low or high value), equal to 4.3% of 9-1-1 EMS activations. Medical conditions accounted for 583 after removing overlap. Of these, 350 were Tier A, led by ischemic stroke (120.2) and EMS-treated cardiac arrest (78.7). Opioid overdose (123.4) led the 233 Tier B events. Those three conditions made up 55% of the medical total. Fire and rescue events added 6.

**Conclusions:** At national rates, a US community of 100,000 can expect its fire and EMS crews to attend about 589 events a year in which time to treatment may change the outcome. That is roughly 11 a week, nearly all of them medical. The count measures how often response time can matter in the events crews attend, and it gives a denominator for valuing a faster response.

## Introduction

How fast a fire or emergency medical services (EMS) system needs to respond depends on how many of the people it serves have an emergency in which arrival time can change the outcome. Fire and EMS systems plan deployment against standards such as National Fire Protection Association (NFPA) 1750 (1). Its 2026 edition merged the career and volunteer standards, NFPA 1710 and 1720, into one document that covers both fire suppression and EMS. Studies that count time-critical interventions find one in about 7% of EMS incidents, and crews use lights and sirens on most (2–4).

Two lines of work come close to such a count. In US emergency departments, conditions that an expert panel judged sensitive to emergency care made up 14.0% of adult visits in 2016 (5). The European Emergency Data Project compared four EMS systems on patients with cardiac arrest, chest pain, or shortness of breath (6). Neither gives a population-based count of the time-critical patients that fire and EMS crews attend, and we found no US estimate of that number. Existing measures cannot supply it either. Dispatch priority over-triages by design. Final acuity misses patients whose condition improved during the call because treatment worked. Intervention counts depend on local scope of practice and on what crews document (2, 3). NFPA’s fire-loss estimates report fires, deaths, and injuries, but they do not say how often a crew arrived to find a person who needed rescue or care (7).

Without such a count, a department has no benchmark for its own tally of critical patients, and any estimate of what call prevention or a faster response is worth has no base number of events to scale from. A national rate gives departments an expected count to compare against their records, and gives planners a way to size how many patients an intervention could reach. We asked how many time-critical events fire and EMS crews attend per 100,000 US residents per year, across medical, fire, and rescue incidents. For each condition we labeled what its count rests on, and we checked the total against national acuity data and studies of time-critical interventions.

## Methods

### Definitions

The estimand is attended time-critical events per 100,000 residents per year. An event qualified when a fire or EMS crew reached a living patient, or a trapped person, with a condition whose outcome plausibly depends on how soon effective treatment starts. The window runs from minutes in cardiac arrest or a compromised airway to a few hours in stroke and myocardial infarction. We counted every attended event of each listed condition. The total therefore measures how often time to treatment can matter, which is a larger number than the events in which time changed the outcome.

For medical conditions, each event is one patient. A person with two such emergencies in a year counts twice, and so do two patients at one incident. An overdose that progresses to cardiac arrest and is worked (resuscitation attempted) counts once. Fire and rescue events are incidents with a patient treated or a person trapped, counted once each, so a fire with two patients treated, or a crash with two people trapped, is one event.

To build the list, we worked through the fire and EMS case mix by mechanism, population, and responder. Medical candidates came from the protocol and provider impression categories that NEMSIS uses and from the conditions that national guidelines treat as time sensitive. For the fire and rescue rows, the frame was the NERIS and NFIRS incident types. A condition entered the count only if a guideline, consensus statement, or outcome study established that it is time sensitive. Two entered instead on clinical reasoning set out in Supplement S1: burns with inhalation injury, and upper airway obstruction from causes other than a foreign body. A medical condition is Tier A when at least one published study found that a longer delay to the first effective treatment went with more deaths or more major disability. That study had to measure the delay itself, on a scale of minutes to a few hours, from onset, collapse, exposure, submersion, or arrival, and the delay could be prehospital or in hospital. Several kinds of evidence did not qualify: descriptions of the untreated course, case series without a comparison group, comparisons of treatment given before and after EMS arrival, studies that found no consistent association, consensus statements, and evidence that a treatment works without timing data. Table 1 names the qualifying study for each Tier A condition. Every other medical condition is Tier B. Fire and rescue events form a separate group, and the evidence for them is operational.

**Table 1.** Tier A medical conditions: a published study links delay to treatment with death or major disability.

| Condition as counted | Per 100,000 | Range | Net after overlap | Incidence basis | Anchor and sources | Time-to-outcome evidence |
|---|---|---|---|---|---|---|
| Ischemic stroke, EMS-transported | 120.2 | 118.2 to 122.0 | 120.2 | Measured shares | CDC 795,000 strokes a year x 87% ischemic x Coverdell EMS share 59.1% (30, 31) | Onset-to-treatment time and outcome after thrombolysis (32) |
| Cardiac arrest, EMS-treated, non-traumatic | 78.7 | 77.5 to 78.7 | 78.7 | Measured shares | CARES 2024 crude incidence in a 174.1 million catchment; national extrapolation 263,711 (27) | Collapse-to-CPR and collapse-to-defibrillation intervals and survival (33); response interval of 8 minutes or less and survival in a multinational prospective registry (34) |
| Septic shock, EMS-attended | 30 | 20 to 40 | 25.8 | Assumption-dependent | CDC 1.7 million adult sepsis cases a year x assumed shock share 15% x Seymour EMS share 40% (13, 14) | Time from ED arrival to bundle completion and in-hospital mortality (35) |
| STEMI, EMS-transported | 30 | 30 to 35 | 28.5 | Assumption-dependent | NIS 2016 to 2020 admissions, transfer-discounted, x Mathews EMS share 60% (36, 37) | Symptom-onset-to-balloon time and one-year mortality (38) |
| Presumed status epilepticus: EMS benzodiazepine-treated seizures | 29.6 | none | 29.0 | Source-derived proxy | Shtull-Leber NEMSIS 2010 to 2014 treatment share x 2024 activations (39) | Time to first benzodiazepine and outcome in pediatric refractory convulsive status epilepticus (40) |
| Intracerebral hemorrhage, EMS-transported | 19.8 | 17.3 to 20.0 | 19.4 | Source-derived proxy | NIS hospitalization rate x Coverdell hemorrhagic-stroke EMS share 75.3% (31, 41) | Time to sustained blood-pressure control and functional outcome in a pooled trial cohort (42) |
| Blunt trauma with hemorrhagic shock, EMS | 19.7 | 8.8 to 38.0 | 19.5 | Assumption-dependent | Hashmi NEMSIS 2019 hypotensive trauma x assumed blunt share 70%, scaled to 2024 volume (43) | Time from massive-transfusion activation to the first cooler and mortality in a randomized transfusion-ratio trial cohort (44); delay to tranexamic acid and loss of its survival benefit in pooled trial data (45) |
| Acute heart failure with respiratory failure (CPAP or ventilation), EMS-attended | 16.6 | 7.3 to 24.9 | 14.9 | Assumption-dependent | Heart failure hospitalizations x ventilated share 8.81% x assumed EMS share 50% (46, 47) | Door-to-furosemide time within 60 minutes and in-hospital mortality (48) |
| Penetrating trauma, exsanguinating subset, EMS | 5.3 | 2.9 to 7.1 | 5.3 | Assumption-dependent | Huebinger NEMSIS 2021 x assumed exsanguinating share 15%, scaled to 2024 volume (49) | Prehospital time and mortality in hypotensive penetrating trauma (50) |
| Severe traumatic brain injury (GCS 8 or less) without hypotension, EMS-transported | 4.2 | 4.2 to 8.5 | 4.2 | Assumption-dependent | CDC TBI hospitalizations x assumed severe, non-hypotensive, and EMS shares (51) | Prehospital time and outcome in traumatic brain injury (50) |
| Drowning, nonfatal, EMS-attended | 2.4 | 1.2 to 3.0 | 2.0 | Assumption-dependent | CDC nonfatal drowning ED visits, all taken as EMS-attended (52) | Submersion duration (time to rescue) and neurologic outcome (53) |
| Carbon monoxide poisoning, unintentional non-fire, EMS or fire department attended | 2.3 | 1.3 to 3.4 | 2.2 | Assumption-dependent | National ED-visit rates x assumed attended share 50% (54, 55) | Exposure-to-hyperbaric-oxygen delay and six-month neurocognitive outcome (56) |
| **Tier A total** | **359** | **318 to 410** | **350** |  |  |  |

**Table 1 notes.** Range is the low and high of the row's own inputs, and the totals add them; no probability attaches to a range. "None" means a single published figure carried without a range. Net after overlap is the row's value after the overlap deduction at the reference coefficients (Methods; Supplement Table S20). Incidence basis: measured shares, a US national count or population rate with every share applied to it measured in a US source; source-derived proxy, a US count of a treatment, procedure, or admission standing in for the condition, or a US count carried across years by a stated ratio; assumption-dependent, at least one share assumed within a published bracket or borrowed from a study of another population. Values are rounded half up; totals are computed from unrounded values and rounded once. Abbreviations: CARES, Cardiac Arrest Registry to Enhance Survival; CDC, Centers for Disease Control and Prevention; CPAP, continuous positive airway pressure; CPR, cardiopulmonary resuscitation; ED, emergency department; EMS, emergency medical services; GCS, Glasgow Coma Scale; NEMSIS, National EMS Information System; NIS, National Inpatient Sample; STEMI, ST-elevation myocardial infarction; TBI, traumatic brain injury.

Six conditions could be defined broadly or narrowly, and we used the narrow definition each time. Cardiac arrest includes only arrests that EMS treated, and septic shock leaves out other severe sepsis; the sensitivity table shows the broader forms of both. The other four are respiratory failure needing assisted ventilation, exsanguinating penetrating trauma, high-risk pulmonary embolism, and hypoglycemia treated by injection or infusion. Every condition, and every fire and rescue event type, is one row of a table we call the ledger. Each row has a point value, which goes into the total, and a range from a low to a high value. Supplements S1 and S2 derive each row. Where no study had measured a share we needed, we assumed one, placing the point usually at a central value and letting the range span the stated bounds. Medical rows also carry one of three labels for what their count rests on, defined in the Table 1 notes: measured shares, source-derived proxy, or assumption-dependent.

Every row needed a US starting point, which we call its anchor: a national count or population rate measured in the United States. A clinical share could come from a study of another population if we said so and gave the reason, but we never borrowed a whole incidence rate from another country. Five further conditions, which we call retained pathways, fit the definition but have no US anchor: severe foreign-body airway obstruction, severe hypothermia, electrical injury, dialysis-access hemorrhage, and device-dependent emergencies. They stay out of the total, and one sensitivity line adds them (Supplement Table S21). Supplement Tables S23 and S24 list the candidates we considered and excluded, with reasons.

### Medical conditions

Thirty medical conditions met these rules. For each, the annual national count of attended events came either from a national registry or dataset, or from a national incidence count multiplied by the share of patients that EMS attended or transported. We preferred national registries (the Cardiac Arrest Registry to Enhance Survival, CARES), then national datasets (the National EMS Information System, NEMSIS, and the National Inpatient Sample, NIS), then Centers for Disease Control and Prevention (CDC) surveillance, and last, peer-reviewed cohorts. Rates use the Census Bureau Vintage 2024 population of 340,100,000 (8).

Eight counts came from NEMSIS or ESO Data Collaborative samples from earlier years. We turned each into a share of its sample’s activations, where an activation is one EMS unit’s response, and applied that share to the 60,298,684 NEMSIS activations of 2024 (9–12). This assumes that each condition’s share of activations held steady since its source year.

Readers should weigh several rows with care, because they join sources of different kinds or rest on assumed shares; rows with at least one assumed or borrowed share are labeled assumption-dependent in Tables 1 and 2. For septic shock, we applied an assumed shock share of 15% (10% to 20%) to the CDC count of adult sepsis, then a 40% EMS arrival share from a single 2012 cohort (13, 14). The anaphylaxis row multiplies a population incidence from a meta-analysis (15) by the share of surveyed patients who said they called 9-1-1 (16), joining two different study designs. An assumed severity fraction converts national emergency department visits for non-opioid overdose into the poisoning row (17). Alcohol intoxication borrows its intubation share from a Swiss emergency department cohort (18) and applies it to US emergency department visits for alcohol-specific diagnoses (19, 20). Opioid overdose is the selected adult nonfatal cohort of Taylor and colleagues. They matched records across agencies by keeping the final responding agency’s record, and they acknowledged that some duplication remained (21).

**Table 2.** Tier B medical conditions: time-critical by guideline or consensus, with no qualifying study of delay and outcome found.

| Condition as counted | Per 100,000 | Range | Net after overlap | Incidence basis | Anchor and sources | Consensus basis |
|---|---|---|---|---|---|---|
| Adult nonfatal opioid-overdose EMS encounters, selected dispositions | 125.5 | 104.4 to 125.5 | 123.4 | Source-derived proxy | NEMSIS 2021 to 2022 encounters, agency-reconciled, scaled to 2024 volume (21) | Respiratory depression progresses to hypoxic arrest without ventilation or naloxone (57); pre-EMS naloxone and fatality at contact (58); no treatment-interval study outside cardiac arrest; EMS response interval and fatality at contact, no overall association (58) |
| Adult asthma or COPD encounters with assisted ventilation, EMS treated and transported | 36.1 | 17.0 to 36.1 | 35.0 | Source-derived proxy | Peters NEMSIS 2018 to 2019 encounters x 9% ventilated, scaled to 2024 volume (59) | Prehospital and in-hospital CPAP compared, mortality, a setting comparison (60); UK national audits found higher mortality only when noninvasive ventilation started after 24 hours (61) |
| Hypoglycemia treated with dextrose or glucagon, EMS | 25.4 | none | 25.4 | Measured shares | Kaufmann NEMSIS 2013 events x 31.9% treated (62) | Untreated severe hypoglycemia progresses to seizure, coma, and death; a crew reverses it (63) |
| Acute alcohol intoxication with airway compromise requiring intubation, EMS-attended | 16.9 | 7.4 to 41.5 | 14.4 | Assumption-dependent | NCHS alcohol-specific ED visits x acute share x Swiss intubation share 2.3% x assumed EMS share (18–20) | Loss of airway protection is fatal without airway management (64) |
| Non-opioid poisoning with respiratory or cardiovascular compromise, EMS | 7.0 | 2.9 to 15.2 | 6.7 | Assumption-dependent | NEDS 2017 non-opioid overdose visits x assumed critical and EMS shares (17) | 2023 AHA focused update on life-threatening poisoning (57) |
| Subarachnoid hemorrhage, EMS-transported | 6.1 | 6.1 to 8.7 | 6.0 | Source-derived proxy | NIS hospitalization rate x Coverdell hemorrhagic-stroke EMS share (31, 65) | Urgent specialist assessment; no timing estimate used (66) |
| Anaphylaxis, EMS-attended | 5 | 4 to 12 | 4.9 | Assumption-dependent | Meta-analytic incidence x survey share who called 9-1-1 (cross-design) (15, 16) | Median time from exposure to arrest in fatal reactions 5 to 30 minutes, an untreated course (67) |
| Unstable arrhythmia needing pacing or cardioversion, EMS | 4.9 | 3.9 to 9.5 | 4.4 | Assumption-dependent | ESO Data Collaborative 2018 to 2021 pacing rate, non-arrest share, applied to 2024 activations (68, 69) | 2020 AHA guidance on unstable bradycardia and tachycardia (70) |
| Pediatric airway-procedure workload proxy | 2.8 | 2.1 to 6.2 | 2.5 | Assumption-dependent | NEMSIS 2019 pediatric airway procedures, arrest and injury removed, scaled to 2024 volume (71, 72) | NAEMSP pediatric airway statement (73); field-managed pediatric respiratory failure outcomes (74) |
| Heat stroke, EMS-attended | 2.6 | 2.6 to 3.5 | 2.6 | Assumption-dependent | NEMSIS heat-related activations x heat-stroke share 17.1% (75, 76) | Cooling within 30 minutes of collapse (77) |
| Acute aortic dissection, EMS-transported | 2.0 | 1.3 to 3.0 | 1.9 | Assumption-dependent | NIS 2012 admissions and the pooled incidence of Gouveia e Melo and colleagues x assumed EMS share 60% (78, 79) | Untreated type A dissection mortality of 1% to 2% per hour, an untreated course (80, 81) |
| Gastrointestinal bleeding presenting in hemorrhagic shock, EMS-attended | 2.0 | 1.4 to 5.6 | 2.0 | Assumption-dependent | NIS 2012 upper GI hemorrhage rate plus assumed lower GI rate x assumed shock and EMS shares (82, 83) | Resuscitation and urgent endoscopy in the ACG guideline (84) |
| High-risk pulmonary embolism, EMS-transported | 1.4 | 1.0 to 1.8 | 1.1 | Assumption-dependent | NIS high-risk rate x assumed EMS share 60% (85, 86) | Reperfusion urgency in the 2019 ESC guideline (87) |
| Obstetric and neonatal emergencies attended in the field (delivery complications, high-acuity hemorrhage, eclamptic seizure, neonatal resuscitation) | 1.4 | 1.0 to 4.2 | 1.1 | Assumption-dependent | NEMSIS 2018 to 2019 counts, scaled to 2024 volume (88–90) | 2020 AHA neonatal resuscitation guideline (91); ACOG postpartum hemorrhage bulletin (92) |
| Ruptured abdominal or thoracic aortic aneurysm, EMS-transported | 1.0 | 0.7 to 1.6 | 1.0 | Assumption-dependent | NIS 2019 admissions x assumed EMS share 75% (93) | Door-to-intervention targets in the SVS guideline (94) |
| Major burn with inhalation injury or airway involvement, EMS-transported | 0.5 | 0.3 to 0.8 | 0.5 | Assumption-dependent | NIS 2020 burn admissions x inhalation share 7.5% x EMS share 85% (95, 96) | Early airway control before supraglottic edema (97) |
| Ruptured ectopic pregnancy with hemodynamic instability, EMS-attended | 0.3 | 0.1 to 1.0 | 0.3 | Assumption-dependent | Medicaid 2004 to 2008 ectopic incidence x complication, instability, and EMS shares (98, 99) | Surgical management in the ACOG bulletin (100) |
| Upper airway obstruction other than foreign body (angioedema, epiglottitis, deep neck infection) requiring airway intervention | 0.3 | 0.1 to 0.7 | 0.3 | Assumption-dependent | Angioedema and epiglottitis visits x intervention and assumed EMS shares (101–103) | Complete upper airway obstruction is fatal within minutes; clinical reasoning |
| **Tier B total** | **241** | **182 to 302** | **233** |  |  |  |

**Table 2 notes.** Columns and rounding as in Table 1. Retained pathways outside the sum (severe foreign-body airway obstruction, severe hypothermia, electrical injury, dialysis-access hemorrhage, device-dependent emergencies) are in Supplement Table S21; their scenario values sum to 5.9 per 100,000. Abbreviations: ACG, American College of Gastroenterology; ACOG, American College of Obstetricians and Gynecologists; AHA, American Heart Association; COPD, chronic obstructive pulmonary disease; CPAP, continuous positive airway pressure; ED, emergency department; EMS, emergency medical services; ESC, European Society of Cardiology; GI, gastrointestinal; NAEMSP, National Association of EMS Physicians; NCHS, National Center for Health Statistics; NEDS, Nationwide Emergency Department Sample; NEMSIS, National EMS Information System; NIS, National Inpatient Sample; SVS, Society for Vascular Surgery.

### Fire and rescue incidents

We counted fire and rescue events in the National Emergency Response Information System (NERIS) public incident dataset, a record-level feed from participating US fire departments (22). We drew aggregate counts by incident type and recorded tactics for the latest complete twelve months, 1 August 2025 to 31 July 2026, a window holding 16,997,003 incidents from 17,103 departments. An incident counted when its type put a person in immediate danger (the thirteen incident types in Table 3) and crews recorded emergency medical care as a tactic. We took that tactic as evidence that a patient was present and treated. Crews recorded that tactic on 70.5% of NERIS medical incidents, so our counts may understate patients treated. We made no adjustment, because we do not know how often crews record it at fires.

**Table 3.** Fire and rescue events with a patient treated or trapped (NERIS public data, August 2025 to July 2026, scaled to national totals), with cross-checks.

| Event type | NERIS incidents (12 months) | Per 100,000 | Range | Net after overlap | NFIRS 2024 analogue, per 100,000 | NFPA casualties x 16% serious, per 100,000 |
|---|---|---|---|---|---|---|
| Motor vehicle crash with a physically trapped patient | 12,659 | 2.3 | 1.6 to 9.0 | 1.1 | Vehicle extrication (NFIRS incident type 352) with an EMS action 2.5; extrication actions on collision or extrication incidents 15.6 |  |
| Structure fire with a patient treated | 3,351 | 2.2 | 2.0 to 2.4 | 2.0 | Structure fires with a civilian casualty 2.16 | 1.4 |
| Other extrication or entrapment (structure, machinery, outside) with a patient treated | 2,194 | 1.4 | 1.3 to 1.6 | 1.1 | Technical rescue with an EMS action 1.12 |  |
| Vehicle fire with a patient treated | 1,095 | 0.7 | 0.7 to 0.8 | 0.6 | Vehicle fires with an EMS action 0.75 | 0.2 |
| Person in water (standing or swift) with a patient treated | 1,028 | 0.7 | 0.6 to 0.7 | 0.3 | Water rescue with an EMS action 0.73 |  |
| Backcountry rescue with a patient treated | 747 | 0.5 | 0.4 to 0.5 | 0.4 |  |  |
| Rope rescue (high, steep, low angle) with a patient treated | 506 | 0.3 | 0.3 to 0.4 | 0.2 |  |  |
| Aviation crash with a patient treated | 274 | 0.2 | 0.2 to 0.2 | 0.1 |  |  |
| Hazardous materials release with a patient treated | 218 | 0.1 | 0.1 to 0.2 | 0.1 |  |  |
| Structural collapse with a patient treated | 194 | 0.1 | 0.1 to 0.1 | 0.1 |  |  |
| Watercraft in distress with a patient treated | 157 | 0.1 | 0.1 to 0.1 | 0.1 |  |  |
| Confined space or trench rescue with a patient treated | 130 | 0.1 | 0.1 to 0.1 | 0.1 |  |  |
| Explosion or overpressure rupture with a patient treated | 130 | 0.1 | 0.1 to 0.1 | 0.1 |  |  |
| **Fire and rescue total** | **22,683** | **9** | **8 to 16** | **6** |  |  |
| Context, not counted: carbon monoxide incident with a patient treated | 783 | 0.5 | 0.5 to 0.6 |  | CO incidents with an EMS action 0.50 |  |
| Context, not counted: incidents typed extrication or entrapment, any position | 45,517 | 29.9 | 27.4 to 32.4 |  |  |  |
| Context, not counted: civilian fire deaths (NFPA 2024) |  | 1.15 |  |  |  | 3,920 deaths |

**Table 3 notes.** Per 100,000 = NERIS incidents x 2.233 / 3,401, the US population in hundred-thousands (Methods); the low value uses 2.047 and the high value 2.418; no completeness adjustment. Net after overlap deducts the stated intersection with the medical rows (reference fractions 0.10 for fires, 0.50 for vehicle entrapment and water rescue, 0.25 for the other rescue rows; Supplement Table S20). For the motor vehicle crash row the NERIS count shown (12,659) is the number of incidents typed extrication or entrapment on which emergency medical care was recorded, which sets the high value; the point applies the 10% physical-entrapment share to the central value (22.7) between the NFIRS reading of extrications performed (15.6) and the NERIS typed reading (29.9), and the low value applies it to the NFIRS reading. Because the NERIS column total includes those typed incidents, the formula does not convert it to the total rate. NFIRS analogues are 2024 counts after dropping aid-given records, scaled to NFPA totals (all incidents x1.296; fires x1.439; structure fires x1.485). The casualty-based column is (NFPA 2024 civilian deaths + injuries x 0.16) / 3,401 by property class. Carbon monoxide poisoning is counted on the medical side (Table 1). Abbreviations: CO, carbon monoxide; EMS, emergency medical services; NERIS, National Emergency Response Information System; NFIRS, National Fire Incident Reporting System; NFPA, National Fire Protection Association.

Not every department reports to NERIS, and participation rose during the window, so we scaled counts up to national totals. NFPA’s 2024 incident total, net of mutual aid, was 2.418 times the NERIS incident count (23). Structure fires gave a lower factor, 2.047, from NFPA’s total over the NERIS count (7). The point uses the midpoint, 2.233, and the range spans both factors. Scaling assumes that departments joining later saw the same mix of events.

A patient treated at an incident typed as an extrication may not have been trapped, so we built the motor vehicle crash row from extrications performed. Extrications per 100,000 lie between a floor of 15.6 and a ceiling of 29.9. The floor comes from extrication actions in the 2024 National Fire Incident Reporting System (NFIRS), where that code is optional. The ceiling comes from NERIS incidents typed as extrication or entrapment, a type assigned before crews arrive. For the point, we applied the 10% physical-entrapment share from a scoping review of UK and European extrication studies (24) to the central value of 22.7. We counted carbon monoxide poisoning once, on the medical side.

To cross-check the NERIS rows, we used the 2024 NFIRS public data release, scaled to NFPA totals. We took the civilian casualty field as marking a patient at a fire, and EMS action codes as marking one at a rescue (25). The fire rows have a second check based on casualties, built from NFPA’s 2024 civilian deaths and injuries and the US Fire Administration’s finding that 16% of residential fire injuries are serious or life-threatening (7, 26).

### Overlap

One patient can appear in more than one row. An overdose can end in a cardiac arrest that crews work, and a crash victim can be trapped and in hemorrhagic shock at the same time. We removed this overlap in a fixed order (an ordered union; Supplement S3.6): cardiac arrest first, then the other medical rows, then the fire and rescue rows. Each row’s qualifying events are its rate multiplied by two factors. Retention is the share of counted records that are distinct events, and eligibility is the share of the row’s events that fit our definition. From the qualifying events we deducted a stated fraction, the share taken to fall inside rows already counted. For opioid overdose, the deduction is half of the CARES treated arrests with an overdose cause, which make up 5.3% of all treated arrests (27).

Every headline figure uses the reference coefficients, in which retention is 1.0, so each record counts as a distinct event; the opioid source had already matched records across agencies (21). A New York City study of assault responses matched 32,202 response records to 26,451 presumed patient encounters, a retention of 82.1% in one city (28). With that result as a guide, the joint lower-count scenario treats 75% of activation records and 80% of hospital admissions as distinct events, and a single-change line uses 85% for activation records. Eligibility is 1.0 for every row except septic shock, which takes the 87.9% community-onset share from Rhee and colleagues, because the CDC sepsis count includes cases that began in hospital, where no crew attends (29). Every fraction is a stated assumption. Supplement Table S20 gives the reference values by row, and the row entries in Supplement S1 and Table S9 give the alternatives. The joint scenarios in Table 4 are internally consistent sets of assumptions applied together, with no probability attached.

**Table 4.** Attended time-critical events: totals and sensitivity (per 100,000 residents per year)

| **Component or variant** | **Per 100,000** | **Range** |
|---|---|---|
| Tier A medical, before overlap | 359 | 318 to 410 |
| Tier B medical, before overlap | 241 | 182 to 302 |
| Fire and rescue, before overlap | 9 | 8 to 16 |
| Medical rows, net after overlap | 583 | 489 to 687 |
| Fire and rescue, net after overlap | 6 | 6 to 10 |
| **Attended time-critical events, net after overlap** | **589** | **494 to 697** |
| Joint lower-count assumptions: activation records 75% and admissions 80% distinct events, stricter eligibility on six rows, overlap fractions at their high values (Supplement S3.6) | 474 |  |
| Joint higher-count assumptions: retention 1.0, every row fully eligible, every overlap fraction zero | 609 |  |
| Activation records at 85% distinct events | 571 |  |
| Stricter eligibility on six rows (opioid, septic shock, status epilepticus, carbon monoxide, hypoglycemia, nonfatal drowning) | 527 |  |
| Overlap fractions at their high values | 570 |  |
| Eight NEMSIS or ESO sample rows at their source-year counts, no scaling to 2024 volume | 515 |  |
| Severe sepsis (97.5, the midpoint of 75 to 120) in place of septic shock (30) | 648 |  |
| EMS-assessed cardiac arrest (110.8, the 2015 ROC extrapolation) in place of EMS-treated | 621 |  |
| Retained pathways added at their scenario values | 595 |  |
| Structure fires with a search performed and no patient treated added | 601 |  |

**Table 4 notes.** Ranges for the components are the totals with every row at its low or high value under the reference coefficients; no probability attaches to them. The joint scenarios are coherent assumption sets applied together (Supplement S3.6 and Table S22). Single-change lines recompute the attended count with one change from the reference. The joint higher-count total equals the sum before overlap by construction, because it sets retention and eligibility to 1.0 and every overlap fraction to zero. Totals are computed from unrounded values and rounded once, half up. Abbreviations: EMS, emergency medical services; NEMSIS, National EMS Information System; ROC, Resuscitation Outcomes Consortium.

### Comparison with other measures

We checked whether the medical count after overlap had the right order of magnitude by comparing it with two other measures. The first was the acuity that EMS clinicians recorded in the 2024 NEMSIS public-release research dataset (12). We counted records coded Critical per 100,000 residents at two points: initial assessment (Initial Patient Acuity, eSituation.13) across all service types, and the end of the call (Final Patient Acuity, eDisposition.19), both across all service types and for 9-1-1 responses only (eResponse.05 = 2205001). We adjusted only for records coded Not Recorded (7701003), scaling the observed counts up on the assumption that those values were missing at random, and left records coded Not Applicable (7701001) out of that adjustment. Supplement Tables S17 and S18 give observed and adjusted rates. Acuity coding alone defined the comparison group, with no filter for patient contact. For the second measure, we turned the population and incident volume that Jarvis and colleagues report into a rate per population (3). Their denominator leaves out cancelled calls, and their list of interventions includes stroke and ST-elevation myocardial infarction (STEMI) alerts.

### Ethics

This study analyzed publicly available, de-identified data (aggregate national statistics and counts from public-use record files) and published literature. It does not involve human subjects as defined in 45 CFR 46.102. The University of Alberta Research Ethics Board (REB 2) reviewed a description of the project and determined on 5 November 2026 that it does not require research ethics board review under Chapter 2 of the Tri-Council Policy Statement (TCPS 2, 2022) and is outside the Board’s mandate. NERIS public data were used under the NERIS Public terms of use; only aggregate counts were queried. This work used the NEMSIS Public-Release Research Dataset. National Highway Traffic Safety Administration, National Emergency Medical Services Information System. The content reproduced from the NEMSIS Database remains the property of the National Highway Traffic Safety Administration. The National Highway Traffic Safety Administration is not responsible for any claims arising from works based on the original Data, Text, Tables, or Figures.

## Results

### Medical events

The twelve Tier A conditions came to 359 events per 100,000 residents per year before overlap and 350 after it (Table 1). Six of them made up 86% of the Tier A total after overlap, led by ischemic stroke (120.2) and EMS-treated non-traumatic cardiac arrest (78.7). Two Tier A rows rest on measured shares, and the other ten are source-derived proxies or depend on assumptions.

The eighteen Tier B conditions added 241 before overlap and 233 after (Table 2). Adult nonfatal opioid overdose encounters with EMS before overlap adjustment (125.5) made up the largest row in either tier (21). Three rows followed at 16.9 to 36.1, and the other fourteen were each 7.0 or less. Of the 583 medical events after overlap, rows built on measured shares contributed 224, source-derived proxies 213, and assumption-dependent rows 146.

### Fire and rescue events

Fire and rescue events with a patient treated or a person trapped came to 9 per 100,000 before overlap (range 8 to 16) and 6 after it (Table 3). Nearly all of the 589 attended events are therefore medical. Motor vehicle crashes with a physically trapped patient (2.3; range 1.6 to 9.0) and structure fires with a patient treated (2.2; range 2.0 to 2.4) were the largest of the thirteen event types. Each of the other eleven came to 1.4 or less. Four event types have a like-for-like NFIRS counterpart, and for each of them the NERIS and NFIRS rates fall within 0.4 per 100,000 of each other.

Incidents typed as extrication or entrapment scaled to 29.9 per 100,000. Crews recorded emergency medical care on 30% of those with that type in the first position. A count of treated patients at these incidents would measure how broadly each system uses the type, so the point rests on the physical-entrapment share instead. Structure fires where crews searched but treated no patient would add 10.2 to 12.1 per 100,000. We left them out, because a search does not show that anyone was inside. For scale, NFPA counted 3,920 civilian fire deaths in 2024 (1.15 per 100,000) (7).

### Totals and sensitivity

Before overlap, the rows summed to 609 per 100,000. Eligibility brought the qualifying events to 605, and the overlap deduction removed 16 (2.6% of the qualifying sum). Attended time-critical events came to 589 per 100,000 residents per year (Table 4). With every row at its low value the total was 494, and with every row at its high value it was 697. The joint lower-count and higher-count scenarios gave 474 and 609. Leaving the eight sample-based rows at their source-year counts, without scaling to 2024 volume, gave 515. Of the other single changes, stricter eligibility on six rows lowered the total most, to 527. Broader definitions raised it: severe sepsis in place of septic shock to 648, and EMS-assessed in place of EMS-treated cardiac arrest to 621 (104). Adding the retained pathways at their scenario values gave 595. The total fell to 571 when 85% of activation records counted as distinct events.

### Comparison with acuity data and system volume

In NEMSIS 2024, EMS clinicians coded 460 activations per 100,000 residents as Critical at initial assessment across all service types, 291 at final acuity, and 240 at final acuity for 9-1-1 responses (12). Leaving Not Applicable aside, Not Recorded made up 26.5%, 35.8%, and 39.2% of those three groups. Adjusting for the missing values on a missing-at-random assumption gave 626, 453, and 395 per 100,000, in the same order. Our medical total of 583 after overlap sits above all three recorded counts, between adjusted final acuity for all service types (453) and adjusted initial acuity (626). The Tier A total of 350 falls between recorded final and recorded initial Critical acuity.

Jarvis and colleagues report a service population of 650,000, about 40,000 incidents a year, and a time-critical intervention in 6.9% to 7.6% of those incidents (3). That works out to 425 to 468 intervention incidents per 100,000 residents a year. The figure falls between our Tier A and medical totals, though it uses a different definition and denominator.

For every 100,000 residents in 2024, US fire departments handled 12,083 incidents, net of mutual aid (23), including 138 structure fires (7). EMS activations numbered 17,730 per 100,000 across all service types and 13,741 for 9-1-1 responses, and 1,944 of those 9-1-1 activations were coded Emergent or Critical at final acuity (12). The 589 attended events equal 4.3% of 9-1-1 EMS activations, 3.3% of all EMS activations, and 30% of 9-1-1 activations coded Emergent or Critical (Figure 1). Because each activation is one unit’s response and one event can draw several units, the share of patients who are time-critical is higher than these shares of activations. The Tier A total after overlap equals 2.5% of 9-1-1 activations.

**Figure 1.**
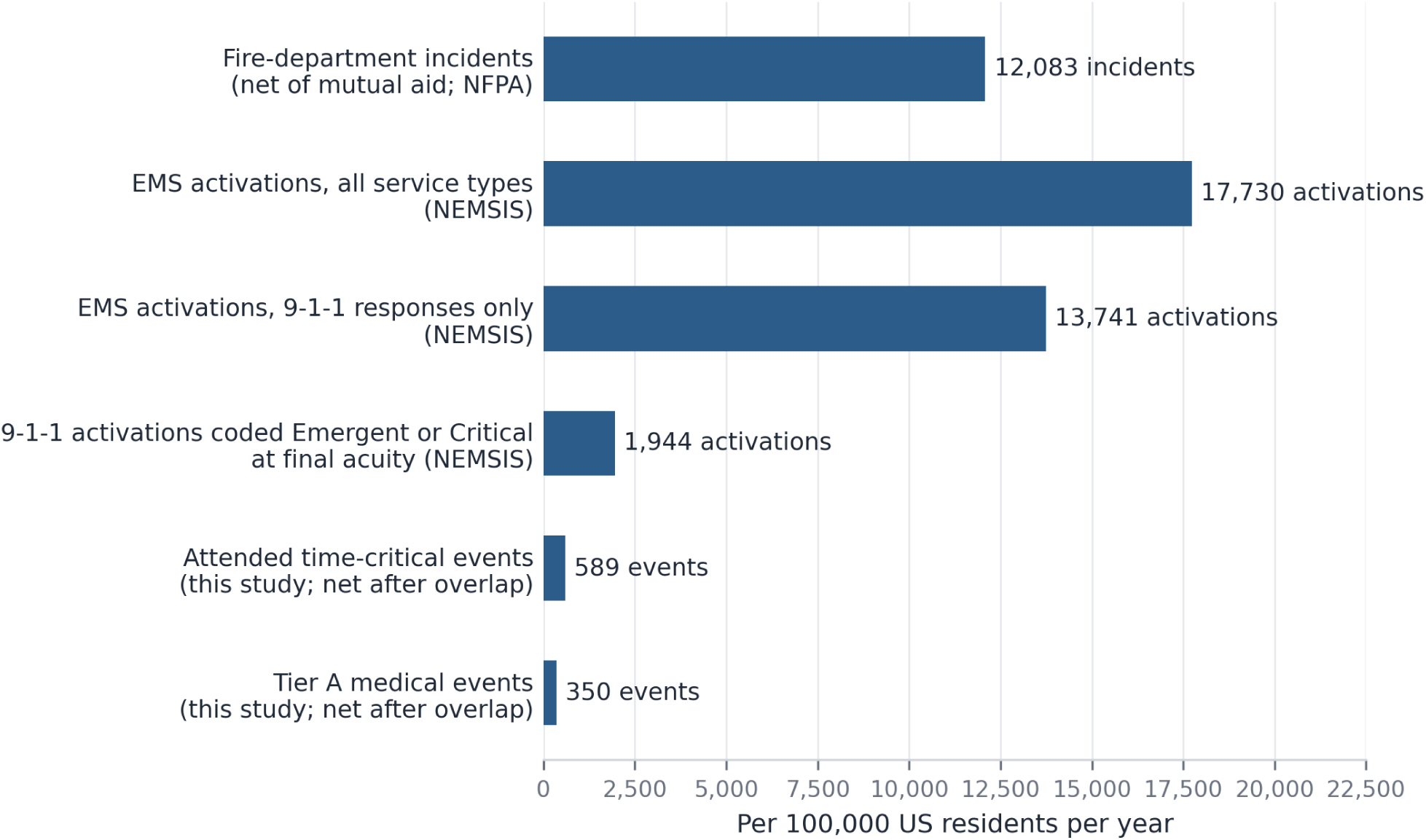
Funnel per 100,000 US residents per year. System volumes are for 2024: fire department incidents; emergency medical services (EMS) activations (all service types and 9-1-1 only); 9-1-1 activations coded Emergent or Critical at final acuity, as recorded. Attended time-critical events net after overlap (fire and rescue events with a patient treated or trapped included) and Tier A medical events net after overlap come from Tables 1 to 4, whose medical anchors span 2001 to 2024 and whose fire and rescue rows use NERIS data from August 2025 to July 2026. Bases labeled on each bar. Abbreviations: EMS, emergency medical services; NEMSIS, National EMS Information System; NFPA, National Fire Protection Association.

## Discussion

At national rates, a US community of 100,000 people can expect its fire and EMS crews to attend about 589 time-critical emergencies a year, or roughly 11 a week. Of these, 350 are in conditions for which a published study ties delay to death or major disability. Opioid overdose, ischemic stroke, and cardiac arrest account for 55% of medical events after overlap. Two other measures give figures of the same order of magnitude, which is consistent with the scale of our estimate: the acuity EMS clinicians record in NEMSIS, and studies that count time-critical interventions.

For fire departments that run EMS, almost all time-critical patients are medical. Medical incidents made up 62% of the incidents departments reported to NERIS in the study window, and fires made up 4% (22). Fire and rescue events account for about 6 of the 589 attended events after overlap, and about one in six of those involve a person trapped in a crashed vehicle. That vehicle row is the least certain on the fire side. Its point applies one borrowed share to the midpoint of two extrication counts that differ about twofold. Because the count covers people, it leaves out the value of fast suppression in limiting fire spread and property loss.

Wherever a definition could be drawn broadly, we used the narrow one, so each looser definition in Table 4 raises the total. Severe sepsis raises it most, to 648. Two other choices push the count up: we treated every activation record and admission as a distinct event, and we scaled earlier samples up to 2024 volume. Discounting duplicate records and admissions, together with stricter eligibility and larger overlap, gives 474. Leaving the samples unscaled gives 515.

Opioid overdose, at 125.5 per 100,000, is the largest row in either tier and also the largest gap in the evidence. Guidance describes opioid-induced respiratory depression progressing to hypoxic arrest, which naloxone reverses (57). Naloxone given before EMS arrival goes with lower fatality at EMS contact (58). For overdose short of cardiac arrest, we found no study relating time to treatment to outcome, and the one analysis of EMS response time and fatality at contact found no overall association (58).

Time to treatment is made up of the response, scene, transport, and in-hospital intervals, and a faster response shortens only the first. The Tier A studies for stroke and sepsis relate outcome to delays of hours, of which the response is a small part. Cardiac arrest runs on minutes. Survival falls with each minute before cardiopulmonary resuscitation and defibrillation (33), and in a Pan-Asian registry a response of 8 minutes or less went with higher survival (34). Putting a value on a minute cut from the response requires knowing, for each condition, how much of the total delay the response controls. We leave that estimate to later work. Deaths in which no responder reached a living patient fall outside this count by design and need their own estimate.

## Limitations

Several EMS shares come from single studies done in different years. Where no study existed, we assumed the share, and those rows carry the assumption-dependent label and wide ranges. The anchors span 2001 to 2024, and we did not adjust them for trends over time. Rows also count on different bases: patients transported by EMS for stroke and STEMI, patients attended by EMS for most rows, and EMS encounters for opioid overdose.

We scaled eight sample-based rows to 2024 activation volume from source years between 2010 and 2022. Activation counts grow both because demand rises and because more agencies contribute to data, and the scaling carries both. Overdose deaths fell in 2024 (105), so scaling the opioid row probably overstates 2024 encounters. The septic shock row starts from the CDC count of adult sepsis, which includes cases that began in hospital, where no crew attends. The 87.9% eligibility share removes them. That share comes from a study that treated sepsis as community-onset if it appeared in the first two hospital days, and some of those patients were not yet in shock when the crew reached them, so the row counts patients who went into shock only after they reached hospital.

The list of conditions is only as complete as the frames we searched. A time-critical condition that none of those frames surfaced is outside the count, and the total is low by whatever it would add. Three Tier A conditions qualified on delays that occurred in hospital (septic shock, heart failure, and intracerebral hemorrhage), so for them the link between prehospital time and outcome is inferred. The status epilepticus evidence comes from a cohort of children with refractory status epilepticus, a narrower group than the benzodiazepine-treated seizures the row counts. Blunt trauma qualifies on two trial cohorts enrolled in hospital, while a systematic review of prehospital time found no consistent association for trauma as a whole (50). The rates are national averages. A community’s own count will move with its age structure, opioid burden, and rurality, and we offer no local adjustment. The cardiac arrest row rests on the CARES extrapolation from its catchment to the whole country.

NEMSIS calls its 2024 file a near complete census covering 99% of US counties, and its manual says the file is not a population-based dataset (12). Both are true: the file carries no population denominator, so our rates use the Census population. Aggregate acuity counts cannot confirm patient contact or show which way the bias runs, and several medical rows draw on the same NEMSIS data. Jarvis and colleagues studied a single agency with its own list of interventions.

NERIS public data come from departments that chose to take part, and they carry no casualty fields. Departments also record tactics inconsistently. The scaled values depend on which departments report. Participation rose during our window, and the window does not line up with 2024, the year of the NFPA totals we scaled to. The entrapment row cannot exclude occupants who were dead when freed, and NFIRS fills its casualty fields only for fires.

## Conclusions

Fire and EMS crews in the United States attend an estimated 589 time-critical events per 100,000 residents per year. After overlap, 350 are in conditions where a published study ties delay to outcome, 233 are in conditions that guidelines or expert consensus treat as time-critical, and 6 are fire and rescue events. National acuity data and studies of time-critical interventions give figures of the same order of magnitude. A system that applies these case definitions to its own records can compare its count with this national rate, allowing for its age structure, opioid burden, and rurality. The count also gives a denominator for estimating what a faster response is worth.

## Data Availability

The row inputs (JSON), the ledger table (CSV and JSON), the NEMSIS acuity counts, the NERIS aggregate counts with the query specification, the NFIRS count file with its counting script, and the code that generates every table in the paper and its supplement are deposited on Zenodo (https://doi.org/10.5281/zenodo.24024025), with a mirror on the Open Science Framework (https://osf.io/b9e76/).

https://osf.io/b9e76/

## Declarations

### Declaration of generative AI in scientific writing

During the preparation of this work, the authors used large language model tools, including Claude (Anthropic), for literature retrieval, citation checking, arithmetic reproduction, aggregate data queries, and drafting. After using these tools, the authors reviewed and edited the content as needed. Deposited code generates every number in the paper from inputs that cite their sources, and each row’s check calculation reproduces its value from the source figures. Daniel Haight checked the anchors, shares, and tier evidence of the ten largest rows, 86% of the total, against the primary sources. A research assistant checked the remaining rows. The authors take full responsibility for the content of this publication.

### Data sharing statement

The following are deposited on Zenodo (https://doi.org/10.5281/zenodo.24024025), with a mirror on the Open Science Framework (https://osf.io/b9e76/): the row inputs (JSON); the ledger table (CSV and JSON); the NEMSIS acuity counts; the NERIS aggregate counts with the query specification; the NFIRS count file with its counting script; and the code that generates every table in this paper and its supplement.

### Authorship statement (CRediT)

Daniel Haight: conceptualization, methodology, data curation, formal analysis, writing (original draft), supervision. Kelly Nisbet: data curation (NEMSIS 2024 acuity extraction and validation), methodology, validation, writing (review and editing). Sunny Li: investigation (condition incidence anchors and source development), validation, writing (review and editing). All authors approved the final manuscript and accept accountability for the work.

