## Supplementary material for "How Many Time-Critical Emergencies? A Population-Based National Estimate of Attended Events per 100,000 US Residents": Online Supplement

Every table in this supplement is generated from the deposited inputs: the ledger (the row table of anchors, shares, arithmetic, and sources, deposited as CSV and JSON), the NERIS Public aggregate file, the NFIRS 2024 count file with the counting script that produced it. The code that generates the tables is deposited with them. The supplement carries its own reference list. Citations are numbered in order of first appearance within the supplement, independently of the main text, and are given as full citations at the end.

**Tables**


- Table S1. Incidence basis definitions.
- Table S2. The 30 medical rows.
- Table S3. NERIS Public provenance.
- Table S4. Type_1 values counted in each fire and rescue row.
- Table S5. All 85 type_1 values in the window, sorted by count.
- Table S6. Incident counts by type token in any type position.
- Table S7. Tactic counts on the type_1 basis.
- Table S8. Scaling factors.
- Table S9. Fire and rescue events with a patient treated or trapped, NERIS Public 1 August 2025 to 31 July 2026, scaled.
- Table S10. Context rows, reported and not counted.
- Table S11. Fire and rescue operational events per 100,000: NERIS rows, NFIRS analogues, and NFPA casualty-based estimates.
- Table S12. NFIRS 2024 records, coverage, and scaling.
- Table S13. Field positions in basicincident.txt used by the counting script.
- Table S14. Definitions applied by the counting script.
- Table S15. All counts in the NFIRS 2024 count file.
- Table S16. NFIRS 2024 cross-check rows, scaled to NFPA totals.
- Table S17. NEMSIS 2024 acuity-code eligibility and missingness by extraction scope.
- Table S18. Recorded rates and missing-at-random scenarios excluding Not Applicable.
- Table S19. Funnel per 100,000 residents, bases labeled.
- Table S20. Ordered union at the reference coefficients (per 100,000).
- Table S21. Retained pathways: no US incidence anchor; scenario values shown outside the sum.
- Table S22. Sensitivity of the attended count (net after overlap, per 100,000).
- Table S23. Candidates considered and excluded.
- Table S24. NERIS incident types considered for the fire and rescue rows and excluded.
- Table S25. Natural-history notes for rows admitted on the untreated course and related rows.


### S1. Condition-by-condition derivation for the 30 medical rows

Each subsection gives one row of the medical ledger: the anchor it starts from, the shares applied to that anchor with their sources, the arithmetic that reproduces the point and the range, the value under another definition of the row, the time-to-outcome evidence (Tier A) or consensus basis (Tier B) that admits the row, the coefficients the ordered union applies to it (S3.6), the sources with the figure taken from each, and the caveats carried in the ledger. Rates are per 100,000 US residents using the Census Bureau Vintage 2024 population of 340,100,000, so a national annual count divides by 3,401 [1]. A row's range runs from the low to the high of its own inputs, and the totals add them; the ranges are not confidence intervals. Each row is shown at the precision the ledger tables use. Totals are computed from the unrounded row values and rounded once, half up, so the displayed row values can sum to a figure that differs from the displayed total in the last digit. Rows appear in the order of the main text's Tables 1 and 2. The check calculation in each subsection is the machine-checked expression that reproduces the point from the source figures; the ledger asserts it at every rebuild.

A row is Tier A when at least one published study measured the interval from onset, collapse, exposure, submersion, or arrival to the first effective intervention (prehospital or in hospital), on the time scale of the definition (minutes to a few hours), and found that a longer interval went with more deaths or more major functional disability; the subsection names the study. Untreated time courses, case series without a comparison group, comparisons of treatment before and after EMS arrival, studies that found no consistent association, consensus statements, and evidence of treatment efficacy alone do not qualify. Tier B rows are time-critical by consensus, and the subsection names the consensus basis. Each row also carries an incidence basis, one of the labels defined in Table S1, which says what kind of evidence the rate rests on. Two rules govern what enters the sum. A row needs a US incidence anchor: a national count or population rate measured in the United States. A clinical share may be borrowed from a study of another population when the row says so and gives the reason (the Swiss intubation share on the alcohol row, the UK and European physical-entrapment share on the vehicle entrapment row); a whole incidence rate from another country is never transferred into the sum. Pathways that fail the first rule are listed with their scenario values in Table S21 and enter the total only in one sensitivity line.

**Table S1. Incidence basis definitions**

| Incidence basis | Definition |
|---|---|
| US source count or rate with measured shares | a US national count or population rate, and every share applied to it is measured in a US source |
| US source-derived proxy | a US count of a treatment, procedure, or admission standing in for the condition, or a US count carried across years or populations by a stated ratio |
| Assumption-dependent scenario | at least one share is assumed within a published bracket or borrowed from a study of another population; the low and high values are scenario endpoints, not confidence limits |
| US operational count, scaled by incident share | a NERIS incident count with a patient treated, scaled to national fire-department incident totals by the structure-fire share (low) and the all-incident share (high) |

Every medical row whose count comes from a NEMSIS or ESO Data Collaborative sample expresses that count as a share of the sample's activations in its source year or years and applies the share to the activations of 2024. The activation totals are 22,532,890 in 2018 and 34,203,087 in 2019 [2], 48,982,990 in 2021 [3], 51,379,493 in 2022 [4], and 54,190,579 in 2023 and 60,298,684 in 2024 [5]. The factors are 60,298,684 / 22,532,890 = 2.676 for 2018 counts; 60,298,684 / 34,203,087 = 1.763 for 2019 counts, which carries the blunt trauma row; 2 x 60,298,684 / (22,532,890 + 34,203,087) = 2.126 a year for counts pooled over 2018 and 2019; 60,298,684 / 48,982,990 = 1.231 for 2021 counts; and 2 x 60,298,684 / (48,982,990 + 51,379,493) = 1.202 a year for counts pooled over 2021 and 2022. A row that starts from a published share applies that share straight to the 60,298,684 activations of 2024. The ratio assumes the event's share of activations held from the source year, and it carries coverage growth and demand growth together. One sensitivity line (Table S22) carries all eight sample-based rows at their source-year counts with the rows' point shares and no scaling, a floor, because the samples covered part of the country in their source years.

**Table S2. The 30 medical rows**

| Section | Row | Tier | Per 100,000 | Range | Net after overlap | Incidence basis | Counting unit |
|---|---|---|---:|---|---:|---|---|
| S1.1 | Ischemic stroke, EMS-transported | A | 120.2 | 118.2 to 122.0 | 120.2 | US source count or rate with measured shares | condition-defined patient events or encounters |
| S1.2 | Cardiac arrest, EMS-treated, non-traumatic | A | 78.7 | 77.5 to 78.7 | 78.7 | US source count or rate with measured shares | condition-defined patient events or encounters |
| S1.3 | Septic shock, EMS-attended | A | 30 | 20 to 40 | 25.8 | Assumption-dependent scenario | condition-defined patient events or encounters |
| S1.4 | STEMI, EMS-transported | A | 30 | 30 to 35 | 28.5 | Assumption-dependent scenario | condition-defined patient events or encounters |
| S1.5 | Presumed status epilepticus: EMS benzodiazepine-treated seizures | A | 29.6 | none | 29.0 | US source-derived proxy | condition-defined patient events or encounters |
| S1.6 | Intracerebral hemorrhage, EMS-transported | A | 19.8 | 17.3 to 20.0 | 19.4 | US source-derived proxy | condition-defined patient events or encounters |
| S1.7 | Blunt trauma with hemorrhagic shock, EMS | A | 19.7 | 8.8 to 38.0 | 19.5 | Assumption-dependent scenario | condition-defined patient events or encounters |

| Section | Row | Tier | Per 100,000 | Range | Net after overlap | Incidence basis | Counting unit |
|---|---|---|---:|---|---:|---|---|
| S1.8 | Acute heart failure with respiratory failure (CPAP or ventilation), EMS-attended | A | 16.6 | 7.3 to 24.9 | 14.9 | Assumption-dependent scenario | condition-defined patient events or encounters |
| S1.9 | Penetrating trauma, exsanguinating subset, EMS | A | 5.3 | 2.9 to 7.1 | 5.3 | Assumption-dependent scenario | condition-defined patient events or encounters |
| S1.10 | Severe traumatic brain injury (GCS 8 or less) without hypotension, EMS-transported | A | 4.2 | 4.2 to 8.5 | 4.2 | Assumption-dependent scenario | condition-defined patient events or encounters |
| S1.11 | Drowning, nonfatal, EMS-attended | A | 2.4 | 1.2 to 3.0 | 2.0 | Assumption-dependent scenario | condition-defined patient events or encounters |
| S1.12 | Carbon monoxide poisoning, unintentional non-fire, EMS or fire department attended | A | 2.3 | 1.3 to 3.4 | 2.2 | Assumption-dependent scenario | condition-defined patient events or encounters |
| S1.13 | Adult nonfatal opioid-overdose EMS encounters, selected dispositions | B | 125.5 | 104.4 to 125.5 | 123.4 | US source-derived proxy | condition-defined patient events or encounters |
| S1.14 | Adult asthma/COPD encounters with assisted ventilation, EMS treated and transported | B | 36.1 | 17.0 to 36.1 | 35.0 | US source-derived proxy | condition-defined patient events or encounters |
| S1.15 | Hypoglycemia treated with dextrose or glucagon | B | 25.4 | none | 25.4 | US source count or rate with measured shares | condition-defined patient events or encounters |
| S1.16 | Acute alcohol intoxication with airway compromise requiring intubation, EMS-attended | B | 16.9 | 7.4 to 41.5 | 14.4 | Assumption-dependent scenario | condition-defined patient events or encounters |
| S1.17 | Non-opioid poisoning with respiratory or cardiovascular compromise, EMS | B | 7.0 | 2.9 to 15.2 | 6.7 | Assumption-dependent scenario | condition-defined patient events or encounters |
| S1.18 | Subarachnoid hemorrhage, EMS-transported | B | 6.1 | 6.1 to 8.7 | 6.0 | US source-derived proxy | condition-defined patient events or encounters |
| S1.19 | Anaphylaxis, EMS-attended | B | 5 | 4 to 12 | 4.9 | Assumption-dependent scenario | condition-defined patient events or encounters |

| Section | Row | Tier | Per 100,000 | Range | Net after overlap | Incidence basis | Counting unit |
|---|---|---|---:|---|---:|---|---|
| S1.20 | Unstable arrhythmia needing pacing or cardioversion, EMS | B | 4.9 | 3.9 to 9.5 | 4.4 | Assumption-dependent scenario | condition-defined patient events or encounters |
| S1.21 | Pediatric airway-procedure workload proxy | B | 2.8 | 2.1 to 6.2 | 2.5 | Assumption-dependent scenario | procedure-derived encounter proxy |
| S1.22 | Heat stroke, EMS-attended | B | 2.6 | 2.6 to 3.5 | 2.6 | Assumption-dependent scenario | condition-defined patient events or encounters |
| S1.23 | Acute aortic dissection, EMS-transported | B | 2.0 | 1.3 to 3.0 | 1.9 | Assumption-dependent scenario | condition-defined patient events or encounters |
| S1.24 | Gastrointestinal bleeding presenting in hemorrhagic shock, EMS-attended | B | 2.0 | 1.4 to 5.6 | 2.0 | Assumption-dependent scenario | condition-defined patient events or encounters |
| S1.25 | High-risk pulmonary embolism, EMS-transported | B | 1.4 | 1.0 to 1.8 | 1.1 | Assumption-dependent scenario | condition-defined patient events or encounters |
| S1.26 | Obstetric and neonatal emergencies attended in the field (delivery complications, high-acuity intrapartum or postpartum hemorrhage, eclamptic seizure, neonatal resuscitation) | B | 1.4 | 1.0 to 4.2 | 1.1 | Assumption-dependent scenario | condition-defined patient events or encounters |
| S1.27 | Ruptured abdominal or thoracic aortic aneurysm, EMS-transported | B | 1.0 | 0.7 to 1.6 | 1.0 | Assumption-dependent scenario | condition-defined patient events or encounters |
| S1.28 | Major burn with inhalation injury or airway involvement, EMS-transported | B | 0.5 | 0.3 to 0.8 | 0.5 | Assumption-dependent scenario | condition-defined patient events or encounters |
| S1.29 | Ruptured ectopic pregnancy with hemodynamic instability, EMS-attended | B | 0.3 | 0.1 to 1.0 | 0.3 | Assumption-dependent scenario | condition-defined patient events or encounters |
| S1.30 | Upper airway obstruction other than foreign body (angioedema, epiglottitis, deep neck infection) requiring airway intervention | B | 0.3 | 0.1 to 0.7 | 0.3 | Assumption-dependent scenario | condition-defined patient events or encounters |

Gross sums of the rows: Tier A 359 (318 to 410); Tier B 241 (182 to 302); medical together 600 (500 to 712). After the ordered union (S3.6) the medical rows give 583 (489 to 687 at the rows' low and high values). The range column reads "none" where the row carries no range.

#### S1.1. Ischemic stroke, EMS-transported (Tier A; US source count or rate with measured shares)

| Item | Value |
| --- | --- |
| Anchor | CDC: approximately 795,000 strokes annually, 87% ischemic. |
| Shares applied | Ischemic-stroke EMS use 59.1% in the Coverdell 2014 to 2019 cohort. Scenario endpoints use the annual shares 58.1% and 60.0%. |
| Arithmetic | 795,000 x 0.87 x (221,293 / 374,428) / 3,401 = 120.2. Scenarios use EMS shares 0.581 and 0.600. |
| Check calculation (machine-checked) | 795000 * 0.87 * (221293 / 374428) / 3401 |
| Other definition | Intracerebral and subarachnoid hemorrhage are separate rows. |
| Time-to-outcome evidence | Onset-to-treatment time and mortality or disability after thrombolysis, Get With The Guidelines-Stroke, 58,353 patients (Saver 2013) [6]. |
| Point and range (per 100,000) | 120.2 (118.2 to 122.0) |
| Counting unit and union coefficients | Counting unit patient events or a population rate; retention 1.0 in every scenario. Eligibility 1, 1, 1 (lower-count, reference, higher-count). Intersection with the union of the preceding rows: overlap fraction 0.01, 0.00, 0.00 (lower-count, reference, higher-count). Reference: qualifying 120.19, deduction 0.00, net 120.19 per 100,000. Ischemic stroke is retained as an eligible presentation. A 1% intersection with earlier arrest is an assumed lower-count stress, not a measured rate. |
| Sources | [7] (data used: More than 795,000 strokes a year; about 87% ischemic); [8]; [6] (data used: Get With The Guidelines-Stroke, 58,353 patients; each 15-minute reduction in onset-to-treatment time associated with lower in-hospital mortality and less disability) |
| Caveats | Repeated strokes count as events. The interval varies the EMS share only; it is not a confidence interval for national incidence. |

#### S1.2. Cardiac arrest, EMS-treated, non-traumatic (Tier A; US source count or rate with measured shares)

| Item | Value |
| --- | --- |
| Anchor | CARES 2024: 137,119 EMS-treated non-traumatic out-of-hospital cardiac arrests reported from a catchment of 174.1 million (52% of the US population); crude incidence 78.7 per 100,000; national extrapolation 263,711 on the population CARES used for 2024 (about 335 million) (CARES 2024 Annual Report, p. 32). |
| Shares applied | Registry-based; no share applied. The published crude incidence is carried as the point because it is a rate and does not depend on the population year. |
| Arithmetic | 78.7 per 100,000 as published. Low value: the national extrapolation divided by the 2024 population, 263,711 / 3,401 = 77.5, which is lower only because CARES extrapolated on a smaller population than the one used here. |
| Check calculation (machine-checked) | 78.7 |
| Other definition | EMS-assessed arrests (including cases without attempted resuscitation) are a broader denominator than this EMS-treated row; the older ROC-based estimate is retained as context only. |
| Time-to-outcome evidence | Collapse-to-CPR and collapse-to-defibrillation intervals and survival (Larsen 1993); response interval of 8 minutes or less and survival in a multinational prospective registry (Tanaka 2018) [9,10]. |
| Point and range (per 100,000) | 78.7 (77.5 to 78.7) |
| Counting unit and union coefficients | Counting unit patient events or a population rate; retention 1.0 in every scenario. Eligibility 1, 1, 1 (lower-count, reference, higher-count). Intersection with the union of the preceding rows: overlap fraction 0.00, 0.00, 0.00 (lower-count, reference, higher-count). Reference: qualifying 78.70, deduction 0.00, net 78.70 per 100,000. First accounting category: EMS-treated nontraumatic arrest. It is a clinical state, not an initiating-cause label. |

| Item | Value |
|---|---|
| Sources | [11] (data used: Page 32: 137,119 events; 174.1 million catchment; 78.7 per 100,000; national extrapolation 263,711); [9] (data used: Survival falls with each minute of delay to CPR, defibrillation, and advanced care); [10] (data used: Pan-Asian Resuscitation Outcomes Study, multinational prospective registry: EMS response time of 8 minutes or less associated with survival to discharge or 30 days (OR 1.52; 95% CI 1.35 to 1.71); bystander CPR OR 1.43; out-of-hospital defibrillation OR 2.31); [12] (data used: Chapter 18: EMS-assessed out-of-hospital cardiac arrest 110.8 per 100,000 in 2015 (356,461; ROC extrapolation, unpublished data); EMS-treated 66 per 100,000. Later updates carry no EMS-assessed figure) |
| Caveats | Narrow definition: worked arrests only. Overlaps the opioid, drowning, and hemorrhagic stroke rows where those conditions arrest; the ordered union carries the opioid deduction. The 263,711 extrapolation and the 340.1 million denominator are from different population years, so the rate, not the count, is carried. Larsen 1993 relates the intervals from collapse to CPR and to defibrillation to survival. Tanaka 2018 relates the call-to-arrival response interval to survival and reports that US studies of response time alone have not been consistent. |

### S1.3. Septic shock, EMS-attended (Tier A; assumption-dependent scenario)

| Item | Value |
|---|---|
| Anchor | About 1.7 million adult sepsis cases a year (CDC About Sepsis). |
| Shares applied | Septic shock share of adult sepsis 15% assumed (10% to 20%); EMS arrival share 40% (Seymour 2012: more than 40% of severe sepsis patients arrive by EMS). |
| Arithmetic | 1,700,000 x 0.15 x 0.40 = 102,000; / 3,401 = 30. Low and high values: shock share 10% gives 20, 20% gives 40. |
| Check calculation (machine-checked) | 1700000 * 0.15 * 0.40 / 3401 |
| Other definition | Severe sepsis in place of septic shock: 1,700,000 x 0.375 to 0.60 x 0.40 = 75 to 120 per 100,000 (97.5 at the midpoint). |
| Time-to-outcome evidence | Time from emergency department arrival to completion of the sepsis bundle and in-hospital mortality (Seymour 2017, New York State mandate cohort) [13]. |
| Point and range (per 100,000) | 30 (20 to 40) |
| Counting unit and union coefficients | Counting unit patient events or a population rate; retention 1.0 in every scenario. Eligibility 0.8, 0.879, 1 (lower-count, reference, higher-count). Intersection with the union of the preceding rows: overlap fraction 0.05, 0.02, 0.00 (lower-count, reference, higher-count). Reference: qualifying 26.37, deduction 0.53, net 25.84 per 100,000. Shock-share source scenarios 10%,15%,20%. Rhee reports 83,620/95,154 (87.9%) community-onset sepsis cases. Applying this to the EMS shock subset is a conditional transport assumption: community onset includes criteria before hospital day 3, not necessarily shock before EMS. The 80%/100% eligibility values and 0%/2%/5% preceding-presentation intersections are assumptions. |
| Sources | [14] (data used: 1.7 million adult cases per year); [15]; [13] (data used: New York State sepsis mandate cohort; longer time to bundle completion associated with higher in-hospital mortality) |
| Caveats | The CDC anchor includes hospital-onset sepsis. Shock and EMS shares form a planning scenario; the source does not directly enumerate community-onset EMS septic shock. |

The severe-sepsis shares 0.375 and 0.60 used in the sensitivity line are assumed; the community-onset eligibility (87.9%) is applied in the ordered union (S3.6), not here.

### S1.4. STEMI, EMS-transported (Tier A; assumption-dependent scenario)

| Item | Value |
|---|---|
| Anchor | 994,300 weighted non-elective adult STEMI admissions in the National Inpatient Sample 2016 to 2020, about 199,000 a year or 58.5 per 100,000 (Kwok 2024). |
| Shares applied | EMS arrival share 60% (Mathews 2011, NCDR ACTION-GWTG 2007 to 2009); admissions discounted to 50 per 100,000 at the point because NIS admissions include interhospital transfers. |
| Arithmetic | 50 x 0.60 = 30; the low value is the point, and the high value is 58.5 x 0.60 = 35 (the 2016 to 2020 mean without the transfer discount). |

| Item | Value |
|---|---|
| Check calculation (machine-checked) | 50 * 0.60 |
| Other definition | This row is limited to STEMI; other acute coronary syndromes are not quantified. |
| Time-to-outcome evidence | Symptom-onset-to-balloon time and one-year mortality in primary angioplasty (De Luca 2004) [16]. |
| Point and range (per 100,000) | 30 (30 to 35) |
| Counting unit and union coefficients | Counting unit patient events or a population rate; retention 1.0 in every scenario. Eligibility 1, 1, 1 (lower-count, reference, higher-count). Intersection with the union of the preceding rows: overlap fraction 0.10, 0.05, 0.00 (lower-count, reference, higher-count). Reference: qualifying 30.00, deduction 1.50, net 28.50 per 100,000. Input already discounts transfers. Assumed 5% reference intersection with preceding arrest; stress 0% to 10%. |
| Sources | [17]; [18]; [16] (data used: Symptom-onset-to-balloon time and one-year mortality) |
| Caveats | The EMS share is dated (2007 to 2009). The transfer discount is a judgment. |

### S1.5. Presumed status epilepticus: EMS benzodiazepine-treated seizures (Tier A; US source-derived proxy)

| Item | Value |
|---|---|
| Anchor | 156,539 benzodiazepine-treated seizure encounters among 93.7 million NEMSIS events 2010 to 2014, a derived share of 0.167% (Shtull-Leber 2017). |
| Shares applied | Share applied to NEMSIS 2024 activations (60,298,684). |
| Arithmetic | (156,539 / 93,700,000) x 60,298,684 = 100,737; / 3,401 = 29.6. |
| Check calculation (machine-checked) | 156539 / 93700000 * 60298684 / 3401 |
| Other definition | Published clinical status-epilepticus incidence estimates of 12.5 to 41 per 100,000 use different populations and definitions; they are not uncertainty bounds for this treated-seizure proxy. |
| Time-to-outcome evidence | Seizure-onset-to-benzodiazepine timing and outcome in pediatric refractory convulsive status epilepticus (Gainza-Lein 2018). The evidence population is narrower than the incidence proxy [19]. |
| Point and range (per 100,000) | 29.6; carried as a point without a range |
| Counting unit and union coefficients | Counting unit activation records; retention 1.0 in the reference, 0.75 in the lower-count scenario, 0.85 in one sensitivity line. Eligibility 0.5, 1, 1 (lower-count, reference, higher-count). Intersection with the union of the preceding rows: overlap fraction 0.05, 0.02, 0.00 (lower-count, reference, higher-count). Reference: qualifying 29.62, deduction 0.59, net 29.03 per 100,000. Reference retains benzodiazepine-treated presumed-status presentation; a half-eligibility value tests narrower onset criteria. Overlap is with preceding neurological/arrest presentations. |
| Sources | [20] (data used: NEMSIS 2010-2014: 93.7 million events; 1.9 million seizure encounters; 156,539 benzodiazepine-treated); [21]; [19]; [22] |
| Caveats | The source operationalizes presumed status epilepticus using benzodiazepine treatment. Duration was unavailable; the cohort may include shorter or less severe seizures. A historical treatment share is transferred to 2024 activation volume. |

### S1.6. Intracerebral hemorrhage, EMS-transported (Tier A; US source-derived proxy)

| Item | Value |
|---|---|
| Anchor | NIS intracerebral hemorrhage hospitalization rate 26.29 per 100,000 in 2016 to 2018; longer-period mean 23.15 (Bako 2022). |
| Shares applied | Hemorrhagic-stroke EMS share 75.3%; annual Coverdell shares 74.6% to 76.0% (Asaithambi 2022). |
| Arithmetic | 26.29 x 0.753 = 19.8; low 23.15 x 0.746 = 17.3; high 26.29 x 0.760 = 20.0. |
| Check calculation (machine-checked) | 26.29 * 0.753 |
| Other definition | Hemorrhages presenting as treated arrest may overlap the cardiac-arrest row. |
| Time-to-outcome evidence | Earlier sustained systolic blood-pressure control and functional outcome in a pooled ICH trial cohort (Wang 2024). The timing association does not estimate EMS response-time causality. |
| Point and range (per 100,000) | 19.8 (17.3 to 20.0) |

| Item | Value |
|---|---|
| Counting unit and union coefficients | Counting unit admissions; retention 1.0 in the reference, 0.80 in the lower-count scenario, 0.90 in one sensitivity line. Eligibility 1, 1, 1 (lower-count, reference, higher-count). Intersection with the union of the preceding rows: overlap fraction 0.05, 0.02, 0.00 (lower-count, reference, higher-count). Reference: qualifying 19.80, deduction 0.40, net 19.40 per 100,000. Assumed fraction of ICH events intersecting preceding arrest or neurological events. CARES exsanguination is not used as a hemorrhagic-stroke count. |
| Sources | [23]; [8]; [24]; [25] |
| Caveats | Hospitalizations are an event proxy and may include transfers. The hemorrhagic-stroke EMS share is applied to the ICH subgroup. |

### S1.7. Blunt trauma with hemorrhagic shock, EMS (Tier A; assumption-dependent scenario)

| Item | Value |
|---|---|
| Anchor | 54,160 EMS trauma patients with SBP below 70 or SBP below 90 with heart rate above 108 in NEMSIS 2019 (Hashmi 2022). |
| Shares applied | Blunt share of hypotensive trauma 70% assumed (PAMPer cohort 82% blunt; 55% to 82% band). The 2019 count is expressed as a share of that year's 34,203,087 activations and applied to the 60,298,684 activations of 2024. |
| Arithmetic | 54,160 x 60,298,684 / 34,203,087 = 95,482 patients at 2024 volume; x 0.70 = 66,837; / 3,401 = 19.7. Low 8.8 is the count as reported at a 55% blunt share (29,788). High 38.0 scales all 89,391 patients with SBP below 90 the same way (157,593) and applies the 82% blunt share (129,226). |
| Check calculation (machine-checked) | 54160 * (60298684 / 34203087) * 0.70 / 3401 |
| Other definition | Penetrating hypotension is in the penetrating trauma row; severe traumatic brain injury is its own row. |
| Time-to-outcome evidence | Time from massive-transfusion activation to arrival of the first cooler and mortality in a randomized transfusion-ratio trial cohort, 680 patients (Meyer 2017); delay to tranexamic acid and the loss of its survival benefit in pooled trial data from 40,138 bleeding patients (Gayet-Ageron 2018) [26,27]. |
| Point and range (per 100,000) | 19.7 (8.8 to 38.0) |
| Counting unit and union coefficients | Counting unit activation records; retention 1.0 in the reference, 0.75 in the lower-count scenario, 0.85 in one sensitivity line. Eligibility 1, 1, 1 (lower-count, reference, higher-count). Intersection with the union of the preceding rows: overlap fraction 0.03, 0.01, 0.00 (lower-count, reference, higher-count). Reference: qualifying 19.65, deduction 0.20, net 19.46 per 100,000. Small assumed mixed-mechanism overlap with preceding penetrating trauma; not a traumatic-arrest deduction from CARES. |
| Sources | [28] (data used: NEMSIS 2019; 54,160 patients with SBP below 70, or SBP below 90 with HR above 108); [26] (data used: Secondary analysis of the PROPPR trial, 680 patients predicted to need massive transfusion at 12 level I trauma centers. Each minute from massive-transfusion activation to arrival of the first cooler raised the odds of death by 5% at 24 hours (OR 1.05; p = 0.035) and at 30 days (OR 1.05; p = 0.016), adjusted for injury severity, physiology, resuscitation intensity, and treatment arm); [27] (data used: Individual patient data from the CRASH-2 (trauma) and WOMAN (postpartum hemorrhage) tranexamic acid trials, 40,138 patients. Treatment delay reduced the benefit. Immediate treatment raised survival from bleeding by more than 70% (OR 1.72; 95% CI 1.42 to 2.10), the benefit fell by 10% for every 15 minutes of delay until 3 hours, and there was no benefit after 3 hours); [29] |
| Caveats | The blunt share is assumed. Arrest is nontraumatic in the separate arrest row, so traumatic-arrest coverage is not established by that row. The activation ratio 1.763 carries the 2019 count to 2024 volume. The admitting evidence comes from two hospital-enrolled trial cohorts: patients predicted to need massive transfusion, and pooled tranexamic acid trials in trauma and postpartum hemorrhage. A systematic review of prehospital time found no consistent association for trauma as a whole (Harmsen 2015). |

### S1.8. Acute heart failure with respiratory failure (CPAP or ventilation), EMS-attended (Tier A; assumption-dependent scenario)

| Item | Value |
|---|---|
| Anchor | 4.9 primary heart failure hospitalizations per 1,000 adults in 2018, about 1.28 million (Salah 2022); invasive ventilation 1.56% and noninvasive ventilation 7.25% of heart failure hospitalizations (NIS 2002 to 2014; Miller 2019). |
| Shares applied | Ventilated share 8.81%; EMS share 50% (22% by ambulance in a trial cohort, Ezekowitz 2016; 61.8% in the EURODEM registry, Harjola 2022). |
| Arithmetic | 1,280,000 x 0.0881 = 112,770; x 0.50 = 56,385; / 3,401 = 16.6. Bounds use EMS shares of 22% (24,810; 7.3) and 75% (84,580; 24.87). |
| Check calculation (machine-checked) | 1280000 * 0.0881 * 0.50 / 3401 |
| Other definition | The broader all-heart-failure hospitalization population is outside this ventilatory-support scenario. |
| Time-to-outcome evidence | Door-to-furosemide time within 60 minutes and in-hospital mortality in acute heart failure (Matsue 2017, REALITY-AHF) [30]. |
| Point and range (per 100,000) | 16.6 (7.3 to 24.9) |
| Counting unit and union coefficients | Counting unit admissions; retention 1.0 in the reference, 0.80 in the lower-count scenario, 0.90 in one sensitivity line. Eligibility 1, 1, 1 (lower-count, reference, higher-count). Intersection with the union of the preceding rows: overlap fraction 0.20, 0.10, 0.00 (lower-count, reference, higher-count). Reference: qualifying 16.60, deduction 1.66, net 14.94 per 100,000. Assumed intersection with preceding STEMI, arrest, and ventilatory presentations. The reference retains the original 10% pulmonary-edema nesting assumption. |
| Sources | [31]; [32]; [33] (data used: 22% arrived by ambulance); [34] (data used: 61.8% arrived by ambulance); [30] |
| Caveats | The EMS share is assumed between two non-US cohorts. STEMI with pulmonary edema can overlap this row; no measured overlap percentage is applied. |

### S1.9. Penetrating trauma, exsanguinating subset, EMS (Tier A; assumption-dependent scenario)

| Item | Value |
|---|---|
| Anchor | 98,040 EMS-attended penetrating trauma patients in NEMSIS 2021 (Huebinger 2023), 28.8 per 100,000. |
| Shares applied | Exsanguinating subset about 15% (10% to 20%) of penetrating trauma. The 2021 count is expressed as a share of that year's 48,982,990 activations and applied to the 60,298,684 activations of 2024. |
| Arithmetic | 98,040 x 60,298,684 / 48,982,990 = 120,688 patients at 2024 volume, 35.5 per 100,000; x 0.15 = 5.3. High 120,688 x 0.20 / 3,401 = 7.1. Low: the count as reported at the 10% share, 98,040 x 0.10 / 3,401 = 2.9. |
| Check calculation (machine-checked) | 98040 * (60298684 / 48982990) * 0.15 / 3401 |
| Other definition | All penetrating trauma: 35.5 per 100,000 at 2024 volume, 28.8 as reported in 2021. |
| Time-to-outcome evidence | Shorter prehospital time associated with lower mortality in hypotensive penetrating trauma (Harmsen 2015 systematic review) [29]. |
| Point and range (per 100,000) | 5.3 (2.9 to 7.1) |
| Counting unit and union coefficients | Counting unit activation records; retention 1.0 in the reference, 0.75 in the lower-count scenario, 0.85 in one sensitivity line. Eligibility 1, 1, 1 (lower-count, reference, higher-count). Intersection with the union of the preceding rows: overlap fraction 0.00, 0.00, 0.00 (lower-count, reference, higher-count). Reference: qualifying 5.32, deduction 0.00, net 5.32 per 100,000. Nontraumatic arrest source excludes traumatic arrests. The input's assumed exsanguinating share selects the modeled critical subset; it is not measured by Huebinger. |
| Sources | [35]; [29]; [3] (data used: 48,982,990 EMS activations from 13,949 agencies in 53 states and territories) |
| Caveats | The exsanguinating share is assumed. Overlaps the blunt trauma row only through mixed-mechanism cases. The activation ratio 1.231 carries the 2021 count to 2024 volume. |

### S1.10. Severe traumatic brain injury (GCS 8 or less) without hypotension, EMS-transported (Tier A; assumption-dependent scenario)

| Item | Value |
|---|---|
| Anchor | 214,110 TBI hospitalizations in 2020 (CDC). |
| Shares applied | Severe share 10% (10% to 20%) assumed; 75% not already counted as hypotensive; 90% EMS-transported. |
| Arithmetic | 214,110 x 0.10 x 0.75 x 0.90 = 14,452; / 3,401 = 4.2. High 8.5 uses a 20% severe share (28,905). |
| Check calculation (machine-checked) | 214110 * 0.10 * 0.75 * 0.90 / 3401 |
| Other definition | All TBI hospitalizations: 63 per 100,000. |
| Time-to-outcome evidence | Shorter prehospital time associated with better outcome in traumatic brain injury (Harmsen 2015 systematic review) [29]. |
| Point and range (per 100,000) | 4.2 (4.2 to 8.5) |
| Counting unit and union coefficients | Counting unit admissions; retention 1.0 in the reference, 0.80 in the lower-count scenario, 0.90 in one sensitivity line. Eligibility 1, 1, 1 (lower-count, reference, higher-count). Intersection with the union of the preceding rows: overlap fraction 0.00, 0.00, 0.00 (lower-count, reference, higher-count). Reference: qualifying 4.20, deduction 0.00, net 4.20 per 100,000. The source input already applies an assumed nonhypotensive fraction to reduce overlap with shock trauma; no second shock deduction. |
| Sources | [36] (data used: 214,110 TBI-related hospitalizations in 2020); [29] |
| Caveats | The severe, nonhypotensive, and EMS shares are assumed. Excluding hypotension is a scenario to reduce overlap, not linked-patient deduplication. |

### S1.11. Drowning, nonfatal, EMS-attended (Tier A; assumption-dependent scenario)

| Item | Value |
|---|---|
| Anchor | 8,111 nonfatal drowning emergency department visits a year in the United States, 2012 to 2021 (CDC Drowning Facts); more than 4,500 drowning deaths a year in 2020 to 2022 (CDC Vital Signs 2024). |
| Shares applied | Every nonfatal drowning that reached an emergency department is taken as EMS-attended; fatal drownings are excluded because those worked by EMS are in the cardiac arrest row and the rest died with no live-patient response, outside the attended count. |
| Arithmetic | 8,111 / 3,401 = 2.4. Low: half the visits EMS-attended, 1.2. High: visits x 1.25 for events treated without an emergency department visit, 3.0. |
| Check calculation (machine-checked) | 8111 / 3401 |
| Other definition | Adding fatal drownings (4,540 in 2024) would give 3.7 per 100,000 and count the worked arrests twice. |
| Time-to-outcome evidence | Submersion duration and neurologic outcome (Quan 2014, open-water drownings in three Western Washington counties) [37]. |
| Point and range (per 100,000) | 2.4 (1.2 to 3.0) |
| Counting unit and union coefficients | Counting unit patient events or a population rate; retention 1.0 in every scenario. Eligibility 0.5, 1, 1 (lower-count, reference, higher-count). Intersection with the union of the preceding rows: overlap fraction 0.30, 0.15, 0.00 (lower-count, reference, higher-count). Reference: qualifying 2.38, deduction 0.36, net 2.03 per 100,000. Nonfatal drowning emergency department visits taken as EMS-attended in the reference (half in the lower-count scenario). Intersection with the preceding arrest row is assumed; resuscitated drowning arrests sit in both sources. |
| Sources | [38] (data used: Nonfatal emergency department visits, 2012-2021); [39]; [37] (data used: Submersion duration and neurologic outcome, open-water drownings in King, Pierce and Snohomish counties, Washington) |
| Caveats | The EMS-attended share of nonfatal drowning emergency department visits is not measured; 100% is the reference assumption and 50% the lower-count value. Resuscitated drowning arrests also appear in the cardiac arrest row. |

### S1.12. Carbon monoxide poisoning, unintentional non-fire, EMS or fire department attended (Tier A; assumption-dependent scenario)

| Item | Value |
|---|---|
| Anchor | Unintentional non-fire carbon monoxide poisoning: 48.3 emergency department visits per million a year, 2007 to 2013 (4.8 per 100,000; Stearns and Sircar 2019); 56.5 per million in 17 states with 74% non-fire (Shin 2023). |
| Shares applied | Attended share 50% assumed (30% to 70%). |
| Arithmetic | Midpoint of 4.2 x 0.50 (2.1) and 4.8 x 0.50 (2.4) = 2.25; low and high values 4.2 x 0.30 = 1.3 and 4.8 x 0.70 = 3.4. |
| Check calculation (machine-checked) | (4.2 + 4.8) / 2 * 0.50 |
| Other definition | Fire-service CO-release records are presented independently in the operational panel. They are not patient counts and are not added to this medical estimate. |
| Time-to-outcome evidence | Exposure-to-hyperbaric-oxygen delay and six-month neurocognitive outcome in 706 selected treated patients (Lee 2021). The comparison uses hours; it does not establish a minute-by-minute response effect [40]. |
| Point and range (per 100,000) | 2.3 (1.3 to 3.4) |
| Counting unit and union coefficients | Counting unit patient events or a population rate; retention 1.0 in every scenario. Eligibility 0.5, 1, 1 (lower-count, reference, higher-count). Intersection with the union of the preceding rows: overlap fraction 0.05, 0.02, 0.00 (lower-count, reference, higher-count). Reference: qualifying 2.25, deduction 0.05, net 2.21 per 100,000. Non-fire CO remains separate from fire toxic inhalation. Reference retains EMS-attended poisoning proxy; half-eligible stress tests acuity. Arrest overlap is assumed. |
| Sources | [41]; [42] (data used: 56.5 ED visits per million, 17 states; 74.0% non-fire); [43] (data used: Oxygen and hyperbaric oxygen timing in carbon monoxide poisoning); [40] (data used: Exposure-to-HBO delay at most 6 hours versus 6 to 24 hours; six-month neurocognitive outcome. Observational treatment-delay evidence, not a two-minute response-time effect) |
| Caveats | The attended share is assumed. NFPA's 80,100 incidents are a 2010 figure; on the 2010 population the rate is 25.9 per 100,000 and on the 2024 population 23.6. |

### S1.13. Adult nonfatal opioid-overdose EMS encounters, selected dispositions (Tier B; US source-derived proxy)

| Item | Value |
|---|---|
| Anchor | 710,263 selected EMS encounters for nonfatal opioid overdose in NEMSIS 2021 to 2022, agency-deduplicated using the final-responding-agency rule (Taylor 2025). |
| Shares applied | The two-year count is expressed as a share of the 100,362,483 activations NEMSIS recorded in 2021 and 2022 and applied to the 60,298,684 activations of 2024. |
| Arithmetic | 710,263 / 100,362,483 x 60,298,684 = 426,732 encounters; / 3,401 = 125.5. Low: the count as reported, 710,263 / 2 / 3,401 = 104.4. |
| Check calculation (machine-checked) | 710263 / (48982990 + 51379493) * 60298684 / 3401 |
| Other definition | Fatal overdoses, patients younger than 18, transfers, and protocol-release dispositions excluded by the study are outside this estimate. No numerical adjustment for these excluded groups is made. |
| Consensus basis | Opioid respiratory depression requires prompt ventilation or reversal. Pre-EMS naloxone is associated with lower fatality at contact (Cano 2026); that contrast does not isolate the effect of a measured treatment delay. In the same study, EMS response-time quartile was not associated with fatality at contact. The authors' supplemental interaction model suggested a difference by urbanicity [44]. |
| Point and range (per 100,000) | 125.5 (104.4 to 125.5) |

| Item | Value |
| --- | --- |
| Counting unit and union coefficients | Counting unit patient events or a population rate; retention 1.0 in every scenario. Eligibility 0.7, 1, 1 (lower-count, reference, higher-count). Intersection with the union of the preceding rows: deduction is a share of the arrest row: 5.3% of treated arrests (the CARES overdose etiology share) times 1.0, 0.5, and 0 across the scenarios. Reference: qualifying 125.47, deduction 2.09, net 123.39 per 100,000. Selected nonfatal adult encounters agency-reconciled using the final-responding-agency rule; residual duplication may remain. A 70% eligibility value tests a narrower acute-threat definition; it is not a measured TCE or naloxone fraction. Arrest intersection is assumed to be half of the CARES-derived overdose-etiology pool in the reference, none/all in the lower-count and higher-count scenarios; it is not the observed surviving-arrest count. The adult etiology share is transported to the all-age worked-arrest rate as a conservative pool scenario. |
| Sources | [45] (data used: NEMSIS 2021-2022; 710,263 encounters over two years); [46]; [44] (data used: NEMSIS 2023: 373,153 opioid overdose activations, 4,865 fatal at EMS contact); [47] (data used: 79,384 drug overdose deaths in 2024; 54,045 involving any opioid; 91.6% unintentional, 5.6% suicide); [3] (data used: 48,982,990 EMS activations from 13,949 agencies in 53 states and territories); [4] (data used: 51,379,493 EMS activations from 13,946 agencies in 54 states and territories) |
| Caveats | Taylor excluded under-18s, deaths, transfers to another EMS unit, and protocol-released encounters. Residual duplicate observations may remain without patient identifiers. The national target remains all residents, so this is an adult-event proxy per total population. Surviving arrests can overlap the arrest row. The incidence basis is now a US source-derived proxy, because the Table 1 notes define that label to include a US count carried across years by a stated ratio. Overdose deaths fell in 2024, so the scaled figure likely overstates 2024 encounters and the count as reported, 104.4, is the low value. |

### S1.14. Adult asthma/COPD encounters with assisted ventilation, EMS treated and transported (Tier B; US source-derived proxy)

| Item | Value |
| --- | --- |
| Anchor | Peters 2023: 1,336,988 asthma/COPD 911 treated-and-transported encounters in NEMSIS 2018 to 2019; 96% adults, of whom 9% received assisted ventilation. |
| Shares applied | Adult-specific ventilation percentage (both source percentages rounded). The two-year count is halved to a year, expressed as a share of the 28,367,988.5 activations a year NEMSIS recorded across 2018 and 2019, and applied to the 60,298,684 activations of 2024. The count as reported is the low value. |
| Arithmetic | 1,336,988 x 0.96 x 0.09 / 2 = 57,758 encounters a year; x 60,298,684 / 28,367,988.5 = 122,770; / 3,401 = 36.1. Low: the count as reported, 57,758 / 3,401 = 17.0. |
| Check calculation (machine-checked) | 1336988 * 0.96 * 0.09 / 2 * (60298684 / 28367988.5) / 3401 |
| Other definition | Count as reported: 17.0 per 100,000 on the fixed reference population, carried as the row's low value. Other causes of noncardiac respiratory failure are not estimated by this row. |
| Consensus basis | Prompt ventilation can stabilize severe respiratory compromise. Goodacre 2014 compares prehospital CPAP with standard care; it does not isolate a treatment-delay effect [48]. |
| Point and range (per 100,000) | 36.1 (17.0 to 36.1) |
| Counting unit and union coefficients | Counting unit activation records; retention 1.0 in the reference, 0.75 in the lower-count scenario, 0.85 in one sensitivity line. Eligibility 1, 1, 1 (lower-count, reference, higher-count). Intersection with the union of the preceding rows: overlap fraction 0.10, 0.03, 0.00 (lower-count, reference, higher-count). Reference: qualifying 36.10, deduction 1.08, net 35.02 per 100,000. Observed-period asthma/COPD assisted-ventilation records; no activation-growth expansion in the joint scenarios. Assumed overlap with preceding arrest/sepsis presentations. |

| Item | Value |
|---|---|
| Sources | [49] (data used: 1,336,988 activations 2018-2019; 9% of adults received assisted ventilation); [2] (data used: NEMSIS national database events by year: 2018, 22,532,890; 2019, 34,203,087); [5] (data used: Acuity from eSituation.13 (initial) and eDisposition.19 (final); 911 filter eResponse.05 = 2205001; 60,298,684 activations in the 2024 public-release dataset; 54,190,579 in the 2023 dataset); [48] (data used: CPAP mortality OR 0.41); [50] (data used: Analysis of the 2008 and 2014 UK national COPD audits. Time is measured from hospital presentation to noninvasive ventilation. In patients acidotic on admission, in-hospital mortality was 15.9% with ventilation within 3 hours, 18.4% at 3 to 24 hours (p = 0.425), and 28.9% after 24 hours (p = 0.002). The mortality comparison is unadjusted: the paper reports no adjusted estimate for time to ventilation, and its authors note that audit data are not collected to the standard of a research study) |
| Caveats | Treated-and-transported asthma/COPD encounters, with no reported exclusion of arrest. Not a national census and not all respiratory failure. The activation ratio 2.126 carries reporting expansion together with workload change. A pair of UK hospital audits relates time from hospital presentation to noninvasive ventilation to in-hospital mortality in COPD exacerbation with acidosis (Jayadev 2019). Its only significant contrast is ventilation after 24 hours, unadjusted, which lies outside the time scale of the definition, so the row stays in Tier B. |

### S1.15. Hypoglycemia treated with dextrose or glucagon (Tier B; US source count or rate with measured shares)

| Item | Value |
|---|---|
| Anchor | 180,561 hypoglycemia EMS events in NEMSIS 2013, projected to 270,945 nationally; 43% not transported (Kaufmann 2019). |
| Shares applied | Kaufmann's mutually exclusive treatment groups: 26.2% dextrose without glucagon plus 5.7% glucagon with or without dextrose; union 31.9%. |
| Arithmetic | 270,945 x (0.262 + 0.057) / 3,401 = 25.4. |
| Check calculation (machine-checked) | 270945 * 0.319 / 3401 |
| Other definition | Other systems report different treatment shares; these are not used to widen this source-specific estimate. |
| Consensus basis | Severe hypoglycemia can progress to seizure, coma, or death; glucagon is the recommended treatment when the person cannot take glucose by mouth (American Diabetes Association 2024). Treatment-defined incidence is broader than this severe subgroup; no delay-effect curve is assigned [51]. |
| Point and range (per 100,000) | 25.4; carried as a point without a range |
| Counting unit and union coefficients | Counting unit activation records; retention 1.0 in the reference, 0.75 in the lower-count scenario, 0.85 in one sensitivity line. Eligibility 0.75, 1, 1 (lower-count, reference, higher-count). Intersection with the union of the preceding rows: overlap fraction 0.02, 0.00, 0.00 (lower-count, reference, higher-count). Reference: qualifying 25.41, deduction 0.00, net 25.41 per 100,000. Reference retains parenteral-treatment presentation. The 75% eligibility and 2% intersection are stricter-definition values; source treatment groups are already a mutually exclusive union. |
| Sources | [52] (data used: 180,561 NEMSIS 2013 events; projected 270,945 nationally; 43% not transported); [53] (data used: IV dextrose 38.0%; IM glucagon 18.3%); [54] (data used: Parenteral glucose 26%; glucagon 3%); [51] (data used: Severe hypoglycemia definition; glucagon when the person cannot take glucose by mouth) |
| Caveats | Treatment-defined hypoglycemia, not a validated severe-hypoglycemia cohort. EMS events were not all transported. |

### S1.16. Acute alcohol intoxication with airway compromise requiring intubation, EMS-attended (Tier B; assumption-dependent scenario)

| Item | Value |
|---|---|
| Anchor | 5.372 million alcohol-specific emergency department visits a year in 2021 to 2022 (NCHS Health E-Stat 109); acute intoxication diagnoses 54.8% of alcohol-related visits (White 2018). |
| Shares applied | Intubation share 2.3% of acute alcohol intoxication ED patients (Sauter 2020, Swiss single center, n = 3,003); EMS arrival share 85% assumed for intubated patients. |

| Item | Value |
|---|---|
| Arithmetic | 5,372,000 x 0.548 = 2,943,900; x 0.023 x 0.85 = 57,550; / 3,401 = 16.9. High 41.5: the GCS-below-9 share of 4.8% (Sauter 2020) at a 100% EMS share (141,310); at 85% it gives 35.3. Low 7.4: 1.0% required critical care in Hennepin County (Klein 2018), x 0.85 (25,020). |
| Check calculation (machine-checked) | 5372000 * 0.548 * 0.023 * 0.85 / 3401 |
| Other definition | Using a depressed-consciousness definition instead of intubation changes eligibility and produces a different workload scenario. |
| Consensus basis | Intubation identifies selected encounters with airway compromise. The included airway guidance supports urgent assessment and airway management; no treatment-delay effect is assigned. |
| Point and range (per 100,000) | 16.9 (7.4 to 41.5) |
| Counting unit and union coefficients | Counting unit patient events or a population rate; retention 1.0 in every scenario. Eligibility 1, 1, 1 (lower-count, reference, higher-count). Intersection with the union of the preceding rows: overlap fraction 0.30, 0.15, 0.00 (lower-count, reference, higher-count). Reference: qualifying 16.90, deduction 2.53, net 14.36 per 100,000. Assumed union intersection with earlier poisoning, trauma and arrest. It is one deduction, not summed pairwise overlaps. |
| Sources | [55] (data used: 5.372 million visits per year); [56] (data used: Acute alcohol diagnoses were 54.8% of alcohol-related visits in 2014); [57] (data used: GCS below 9 without traumatic brain injury in 4.8%; intubation in 2.3% of 3,003 patients); [58] (data used: 1.0% required critical care; 0.31% hypoxic respiratory failure; n = 31,364); [59] |
| Caveats | The intubation share is transferred from a Swiss ED; the EMS share is assumed. Intubation does not establish that every untreated case would have died. Sedative co-ingestion can overlap the poisoning row. |

### S1.17. Non-opioid poisoning with respiratory or cardiovascular compromise, EMS (Tier B; assumption-dependent scenario)

| Item | Value |
|---|---|
| Anchor | 967,615 drug overdose emergency department visits in NEDS 2017, of which 305,623 opioid, leaving about 662,000 non-opioid (Vivolo-Kantor 2020). |
| Shares applied | Critical share 6% assumed; EMS share 60% assumed. The National Poison Data System major-effect table is the intended anchor and was not available. |
| Arithmetic | 662,000 x 0.06 x 0.60 = 23,830; / 3,401 = 7.0. Low 2.9: 3% critical and 50% EMS (9,930). High 15.2: 12% critical and 65% EMS (51,640). |
| Check calculation (machine-checked) | 662000 * 0.06 * 0.60 / 3401 |
| Other definition | Opioid overdose is its own row. |
| Consensus basis | Consensus: 2023 AHA focused update on life-threatening poisoning (Lavonas 2023) [46]. |
| Point and range (per 100,000) | 7.0 (2.9 to 15.2) |
| Counting unit and union coefficients | Counting unit patient events or a population rate; retention 1.0 in every scenario. Eligibility 1, 1, 1 (lower-count, reference, higher-count). Intersection with the union of the preceding rows: overlap fraction 0.10, 0.05, 0.00 (lower-count, reference, higher-count). Reference: qualifying 7.00, deduction 0.35, net 6.65 per 100,000. Source critical and EMS shares already applied; remaining cross-source drug-mixture/arrest overlap is assumed. |
| Sources | [60] (data used: NEDS 2017); [46] |
| Caveats | Both shares are assumed; the National Poison Data System major-effect table would be the preferred anchor and was not available. |

### S1.18. Subarachnoid hemorrhage, EMS-transported (Tier B; US source-derived proxy)

| Item | Value |
|---|---|
| Anchor | NIS subarachnoid hemorrhage hospitalization rate 8.12 per 100,000 in 2020 (Feler); another estimate 11.4 (Xia 2023). |
| Shares applied | Hemorrhagic-stroke EMS share 75.3%; annual Coverdell shares 74.6% to 76.0% (Asaithambi 2022). |
| Arithmetic | 8.12 x 0.753 = 6.1; low 8.12 x 0.746 = 6.1; high 11.4 x 0.760 = 8.7. |

| Item | Value |
|---|---|
| Check calculation (machine-checked) | 8.12 * 0.753 |
| Other definition | SAH with treated arrest may overlap the cardiac-arrest row. |
| Consensus basis | Urgent specialist assessment and stabilization are required (2023 AHA/ASA guideline on aneurysmal subarachnoid hemorrhage, Hoh 2023); no qualifying SAH treatment-timing estimate is used in this synthesis [61]. |
| Point and range (per 100,000) | 6.1 (6.1 to 8.7) |
| Counting unit and union coefficients | Counting unit admissions; retention 1.0 in the reference, 0.80 in the lower-count scenario, 0.90 in one sensitivity line. Eligibility 1, 1, 1 (lower-count, reference, higher-count). Intersection with the union of the preceding rows: overlap fraction 0.05, 0.02, 0.00 (lower-count, reference, higher-count). Reference: qualifying 6.11, deduction 0.12, net 5.99 per 100,000. Assumed fraction of SAH events intersecting preceding arrest or neurological events; one union deduction, not pairwise subtraction. |
| Sources | [62] (data used: 448,655 SAH admissions 2001-2020; 9.51 per 100,000 person-years; 8.12 in 2020); [63] (data used: Spontaneous subarachnoid hemorrhage incidence 11.4 per 100,000 (2007-2017)); [8]; [61] |
| Caveats | Hospitalizations are an event proxy. The hemorrhagic-stroke transport share is transferred to SAH; ICH timing evidence is not assigned to SAH. |

### S1.19. Anaphylaxis, EMS-attended (Tier B; assumption-dependent scenario)

| Item | Value |
|---|---|
| Anchor | All-cause anaphylaxis incidence in North America 43 per 100,000 a year (Puhringer 2023 meta-analysis; global current-window estimate 46, 95% CI 21 to 103). |
| Shares applied | Share of patients who called 911 for their most recent reaction: 10% of 344 self-reported cases, 12% of the 261 confirmed cases (Wood 2014, national telephone survey). |
| Arithmetic | 43 x 0.12 = 5.2, carried as 5. Low: 43 x 0.10 = 4.3. High: 103 x 0.12 = 12.4. |
| Check calculation (machine-checked) | 43 * 0.12 |
| Other definition | Anaphylaxis presenting to any emergency department (about 57% of cases in Wood 2014) is not counted; only EMS-attended events qualify. |
| Consensus basis | Rapid progression to cardiorespiratory compromise and urgency of epinephrine in anaphylaxis guidance. No death or major-functional-outcome timing estimate is used here; hospitalization-only comparisons do not define this tier. |
| Point and range (per 100,000) | 5 (4 to 12) |
| Counting unit and union coefficients | Counting unit patient events or a population rate; retention 1.0 in every scenario. Eligibility 1, 1, 1 (lower-count, reference, higher-count). Intersection with the union of the preceding rows: overlap fraction 0.05, 0.02, 0.00 (lower-count, reference, higher-count). Reference: qualifying 5.00, deduction 0.10, net 4.90 per 100,000. Assumed intersection with preceding arrest/respiratory presentations. |
| Sources | [64]; [65]; [66] (data used: Fatal reactions: median time from exposure to cardiorespiratory arrest 30 minutes for food, 15 for venom, 5 for iatrogenic causes) |
| Caveats | Cross-design join disclosed in Methods: a meta-analytic incidence multiplied by a retrospective self-reported survey share. The incidence is a continent-level average across 37 anaphylaxis definitions. |

### S1.20. Unstable arrhythmia needing pacing or cardioversion, EMS (Tier B; assumption-dependent scenario)

| Item | Value |
|---|---|
| Anchor | 13,270 transcutaneous pacing patients among 31,158,400 ESO encounters 2018 to 2021, 0.0426% (Smida 2025); 34.9% of the 12,188 with documented times followed arrest resuscitation; 20.4% of the rest progressed to arrest. Cardioversion-to-pacing ratio 0.26 (NEMSIS 2018 subset). |
| Shares applied | Pacing rate applied to the 60,298,684 NEMSIS activations of 2024; non-arrest share 65.1%; non-progression share 79.6%; cardioversions 52% post-ROSC excluded (Cowley 2021, synchronized cardioversion, one UK service). |

| Item | Value |
|---|---|
| Arithmetic | 60,298,684 x 0.000426 = 25,687 pacing; x 0.651 x 0.796 = 13,311; cardioversion 25,687 x 0.26 = 6,679, x 0.48 not post-ROSC = 3,206; total 16,517; / 3,401 = 4.9. Low 3.9 drops cardioversion (13,311); high 9.5 keeps every pacing and cardioversion patient (32,366). |
| Check calculation (machine-checked) | (60298684 * 0.000426 * 0.651 * 0.796 + 60298684 * 0.000426 * 0.26 * 0.48) / 3401 |
| Other definition | Arrhythmia patients who arrest are in the arrest row. |
| Consensus basis | Consensus: 2020 AHA adult advanced life support guidance on unstable bradycardia and tachycardia (Panchal 2020) [67]. |
| Point and range (per 100,000) | 4.9 (3.9 to 9.5) |
| Counting unit and union coefficients | Counting unit activation records; retention 1.0 in the reference, 0.75 in the lower-count scenario, 0.85 in one sensitivity line. Eligibility 1, 1, 1 (lower-count, reference, higher-count). Intersection with the union of the preceding rows: overlap fraction 0.20, 0.10, 0.00 (lower-count, reference, higher-count). Reference: qualifying 4.86, deduction 0.49, net 4.37 per 100,000. Source already excludes post-arrest and progression-to-arrest pacing. Residual overlap here is with other preceding cardiac presentations, not a repeat arrest exclusion. |
| Sources | [68] (data used: ESO 2018-2021); [69] (data used: Prehospital synchronized cardioversion; 49 of 94 were post-ROSC); [67] |
| Caveats | Weakest joint: ESO-to-national scaling. |

### S1.21. Pediatric airway-procedure workload proxy (Tier B; assumption-dependent scenario)

| Item | Value |
|---|---|
| Anchor | NEMSIS 2019: 8,075 pediatric airway procedures (bag-valve-mask 3,997, tracheal intubation 3,165, supraglottic airway 582, CPAP or BiPAP 331) among 1,148,943 pediatric encounters; 16.9% in cardiac arrest, 16.6% injured (Hanlin 2022). |
| Shares applied | Scaled to NEMSIS 2024 volume by 60,298,684 / 34,203,087 = 1.763 (14,236, 4.2 per 100,000); arrest and trauma patients removed (about one third). |
| Arithmetic | 8,075 x (60,298,684 / 34,203,087) x (1 - 0.169 - 0.166) / 3,401 = 2.8. This is an explicit disjointness scenario, not a measured non-arrest/non-trauma patient count. |
| Check calculation (machine-checked) | 8075 * (60298684 / 34203087) * (1 - 0.169 - 0.166) / 3401 |
| Other definition | Some pediatric arrests and injuries may also occur in other rows; cross-source linkage is unavailable. |
| Consensus basis | Consensus: 2022 NAEMSP pediatric airway position statement (Harris 2022); Gausche 2000 for outcomes of pediatric respiratory failure managed in the field [70,71]. |
| Point and range (per 100,000) | 2.8 (2.1 to 6.2) |
| Counting unit and union coefficients | Counting unit activation records; retention 1.0 in the reference, 0.75 in the lower-count scenario, 0.85 in one sensitivity line. Eligibility 1, 1, 1 (lower-count, reference, higher-count). Intersection with the union of the preceding rows: overlap fraction 0.20, 0.10, 0.00 (lower-count, reference, higher-count). Reference: qualifying 2.80, deduction 0.28, net 2.52 per 100,000. The input calculation already subtracts the assumed maximum arrest/injury union; this is not a source-verified exclusion. Remaining intersection concerns preceding noninjury presentations; neonatal and maternal events are distinct patients. |
| Sources | [72] (data used: 1,148,943 pediatric encounters; BVM 3,997; tracheal intubation 3,165; supraglottic airway 582; CPAP or BiPAP 331; 16.9% cardiac arrest; 16.6% injured); [73] (data used: ESO 2019 pediatric airway procedure rate; cross-check for the pediatric respiratory failure row (6.2 per 100,000)); [70]; [71] |
| Caveats | Arrest and injury percentages may overlap. The prior subtraction of both as disjoint groups is unsupported. For the reference calculation, their union is set to the conservative maximum 33.5%; an overlapping union of 16.9% gives a larger eligible share. Multiple procedure records per patient and incomplete coverage remain possible. |

### S1.22. Heat stroke, EMS-attended (Tier B; assumption-dependent scenario)

| Item | Value |
|---|---|
| Anchor | 261,687 heat-related EMS activations over five warm seasons 2020 to 2024, 52,337 a season (Saha 2026); heat stroke 17.1% of 34,814 heat-related EMS records (Yeargin 2020). |
| Shares applied | Heat stroke share 17.1%. |
| Arithmetic | 261,687 / 5 x 0.171 / 3,401 = 2.6. High value: a 23% heat-stroke share gives 3.5. Heat mortality is reported separately and is not used as an attended-incidence bound. |
| Check calculation (machine-checked) | 261687 / 5 * 0.171 / 3401 |
| Other definition | All heat-related activations: 15.4 per 100,000. |
| Consensus basis | Clinical guidance supports rapid cooling in heat stroke (Epstein and Yanovich 2019). No quantitative mortality or major-disability timing estimate is used for this row [74]. |
| Point and range (per 100,000) | 2.6 (2.6 to 3.5) |
| Counting unit and union coefficients | Counting unit activation records; retention 1.0 in the reference, 0.75 in the lower-count scenario, 0.85 in one sensitivity line. Eligibility 1, 1, 1 (lower-count, reference, higher-count). Intersection with the union of the preceding rows: overlap fraction 0.03, 0.01, 0.00 (lower-count, reference, higher-count). Reference: qualifying 2.60, deduction 0.03, net 2.57 per 100,000. Heat-stroke share already applied; assumed intersection with preceding arrest. |
| Sources | [75] (data used: NEMSIS 2020-2024; 261,687 heat-related activations; 16.46 per 100,000); [76] (data used: 5,955 of 34,814 heat-related records were heat stroke); [77]; [74] |
| Caveats | The annualization of the five-season count is our arithmetic. |

### S1.23. Acute aortic dissection, EMS-transported (Tier B; assumption-dependent scenario)

| Item | Value |
|---|---|
| Anchor | 8,875 primary-diagnosis aortic dissection admissions in the National Inpatient Sample 2012, about 2.8 per 100,000 on the 2012 population (Zimmerman 2016); pooled population incidence 4.8 per 100,000 (Gouveia e Melo 2022). |
| Shares applied | Hospital-reaching incidence 3.0 to 3.5 per 100,000; EMS share 60% assumed. |
| Arithmetic | 3.3 x 0.60 = 2.0; low and high values 2.2 x 0.60 = 1.3 and 5.0 x 0.60 = 3.0. |
| Check calculation (machine-checked) | 3.3 * 0.60 |
| Other definition | People who die before hospitalization are absent from the hospital anchor. Some, but not all, may appear in an EMS-treated arrest series. |
| Consensus basis | Consensus: untreated type A dissection carries a mortality of 1% to 2% per hour early after onset (IRAD, Hagan 2000; 2022 ACC/AHA guideline), an untreated time course [78,79]. |
| Point and range (per 100,000) | 2.0 (1.3 to 3.0) |
| Counting unit and union coefficients | Counting unit admissions; retention 1.0 in the reference, 0.80 in the lower-count scenario, 0.90 in one sensitivity line. Eligibility 1, 1, 1 (lower-count, reference, higher-count). Intersection with the union of the preceding rows: overlap fraction 0.10, 0.05, 0.00 (lower-count, reference, higher-count). Reference: qualifying 2.00, deduction 0.10, net 1.90 per 100,000. Assumed intersection with preceding arrest or stroke presentations. |
| Sources | [80]; [81]; [79]; [78] (data used: Untreated type A dissection mortality of 1% to 2% per hour early after onset) |
| Caveats | A US EMS-arrival share was not used; 60% is assumed. Other incidence anchors span hospital-reaching and population estimates. |

### S1.24. Gastrointestinal bleeding presenting in hemorrhagic shock, EMS-attended (Tier B; assumption-dependent scenario)

| Item | Value |
|---|---|
| Anchor | Upper gastrointestinal hemorrhage 67 per 100,000 adults in NIS 2012 (Wuerth and Rockey 2018) plus lower gastrointestinal bleeding, assumed about 35 per 100,000 adults; adults are 78% of residents. |

| Item | Value |
|---|---|
| Shares applied | Shock share 5% assumed; EMS share 50% assumed. |
| Arithmetic | (67 + 35) x 0.78 = 80 per 100,000 residents; x 0.05 x 0.50 = 2.0; low 1.4 uses a 3.4% shock share; high 5.6 uses 14%. |
| Check calculation (machine-checked) | (67 + 35) * 0.78 * 0.05 * 0.50 |
| Other definition | Bleeding without the shock criterion is outside this scenario. |
| Consensus basis | Consensus: resuscitation and urgent endoscopy in the ACG upper gastrointestinal bleeding guideline (Laine 2021) [82]. |
| Point and range (per 100,000) | 2.0 (1.4 to 5.6) |
| Counting unit and union coefficients | Counting unit patient events or a population rate; retention 1.0 in every scenario. Eligibility 1, 1, 1 (lower-count, reference, higher-count). Intersection with the union of the preceding rows: overlap fraction 0.05, 0.02, 0.00 (lower-count, reference, higher-count). Reference: qualifying 2.00, deduction 0.04, net 1.96 per 100,000. Shock and EMS shares already applied; assumed preceding-arrest intersection. |
| Sources | [83] (data used: 67 per 100,000 adults, 2012); [84] (data used: Index mortality 1.9%); [82] (data used: Resuscitation and urgent endoscopy in upper gastrointestinal bleeding) |
| Caveats | The lower gastrointestinal rate, shock share, and EMS share are assumptions. Treated mortality in a broad GI-bleeding cohort does not establish untreated mortality in the shock subgroup. |

### S1.25. High-risk pulmonary embolism, EMS-transported (Tier B; assumption-dependent scenario)

| Item | Value |
|---|---|
| Anchor | High-risk (shock or arrest) pulmonary embolism as primary diagnosis 2.3 per 100,000 in 2014 (Stein 2021); 2.4% of pulmonary embolism admissions are high-risk (NRD 2016 to 2019; Sedhom 2022). |
| Shares applied | EMS share 60% assumed for unstable patients (21.5% for all pulmonary embolism in an ED cohort, Rouleau 2023). |
| Arithmetic | 2.3 x 0.60 = 1.4; low and high values 2.3 x 0.45 = 1.0 and 2.3 x 0.80 = 1.8. |
| Check calculation (machine-checked) | 2.3 * 0.60 |
| Other definition | Hemodynamically stable pulmonary embolism is outside this high-risk scenario. |
| Consensus basis | Consensus: reperfusion urgency in high-risk pulmonary embolism (2019 ESC guideline, Konstantinides 2020) [85]. |
| Point and range (per 100,000) | 1.4 (1.0 to 1.8) |
| Counting unit and union coefficients | Counting unit patient events or a population rate; retention 1.0 in every scenario. Eligibility 1, 1, 1 (lower-count, reference, higher-count). Intersection with the union of the preceding rows: overlap fraction 0.40, 0.20, 0.00 (lower-count, reference, higher-count). Reference: qualifying 1.40, deduction 0.28, net 1.12 per 100,000. High-risk source includes shock or arrest; assumed arrest-overlap scenarios. |
| Sources | [86]; [87]; [88]; [85] (data used: Reperfusion urgency in high-risk pulmonary embolism) |
| Caveats | Treated nontraumatic PE arrests may overlap the arrest row. The EMS share is assumed. |

### S1.26. Obstetric and neonatal emergencies attended in the field (delivery complications, high-acuity intrapartum or postpartum hemorrhage, eclamptic seizure, neonatal resuscitation) (Tier B; assumption-dependent scenario)

| Item | Value |
|---|---|
| Anchor | NEMSIS 2018 (one year, 22,532,890 activations; Cash 2021): 629 intrapartum or postpartum hemorrhage activations (55% high acuity) and about 1,000 hypertensive or eclampsia activations. NEMSIS 2018 to 2019 (two years, 56,735,977 activations; Cash 2024): 8,614 EMS-attended deliveries, 1,712 delivery complications, 21 cord prolapse, 37 non-cephalic presentations. NEMSIS 2018 to 2019 (Peters 2026): 580 neonatal resuscitations. |
| Shares applied | Scaled to 2024 volume (60,298,684 activations) by the ratio of annual activations: x2.676 for 2018 counts; x2.126 a year for two-year 2018 to 2019 counts. Eclamptic seizures assumed to be half of the hypertensive or eclampsia activations. |

| Item | Value |
| --- | --- |
| Arithmetic | Complications 1,712 / 2 x 2.126 = 1,820 (0.54); high-acuity hemorrhage 629 x 0.55 x 2.676 = 926 (0.27); eclamptic seizures 1,000 x 0.50 x 2.676 = 1,338 (0.39); neonatal resuscitation 580 / 2 x 2.126 = 617 (0.18). Sum 1.4 per 100,000. Low 1.0 omits the eclampsia component. High 4.2 counts every field delivery (8,614 / 2 x 2.126 = 9,157, 2.69), all hemorrhage activations (1,683, 0.49), all hypertensive or eclampsia activations (2,676, 0.79), and neonatal resuscitations (0.18). |
| Check calculation (machine-checked) | (1712 / 2 * (60298684 / 28367988.5) + 629 * 0.55 * (60298684 / 22532890) + 1000 * 0.50 * (60298684 / 22532890) + 580 / 2 * (60298684 / 28367988.5)) / 3401 |
| Other definition | Every EMS field delivery counted as an event, with the other components at their point values: 3.5 per 100,000 (inclusive bound). Deliveries that reach hospital before birth are excluded. |
| Consensus basis | Consensus: the 2020 AHA neonatal resuscitation guideline (Aziz 2020) and the ACOG postpartum hemorrhage bulletin (2017) [89,90]. |
| Point and range (per 100,000) | 1.4 (1.0 to 4.2) |
| Counting unit and union coefficients | Counting unit activation records; retention 1.0 in the reference, 0.75 in the lower-count scenario, 0.85 in one sensitivity line. Eligibility 1, 1, 1 (lower-count, reference, higher-count). Intersection with the union of the preceding rows: overlap fraction 0.40, 0.20, 0.00 (lower-count, reference, higher-count). Reference: qualifying 1.40, deduction 0.28, net 1.12 per 100,000. Composite-source maternal overlap and intersection with preceding seizure/neonatal respiratory presentations; assumed residual-union approximation. |
| Sources | [91] (data used: NEMSIS 2018: 107,771 obstetric activations, 3,489 out-of-hospital deliveries, 629 intrapartum or postpartum hemorrhage activations (55% high acuity), about 1,000 hypertensive or eclampsia activations, 332 ectopic pregnancy activations (47% high acuity)); [92] (data used: 8,614 deliveries; 1,712 delivery complications; 5,749 newborns); [93] (data used: 580 neonatal resuscitations; 31.7% cardiac arrest); [2] (data used: NEMSIS national database events by year: 2018, 22,532,890; 2019, 34,203,087); [89] (data used: Neonatal resuscitation rationale); [90] (data used: Postpartum hemorrhage management) |
| Caveats | The eclamptic share is assumed. Delivery complications, maternal hemorrhage, and hypertensive events may overlap within this composite; neonatal resuscitation concerns the infant, a different patient. |

### S1.27. Ruptured abdominal or thoracic aortic aneurysm, EMS-transported (Tier B; assumption-dependent scenario)

| Item | Value |
| --- | --- |
| Anchor | 4,625 ruptured abdominal aortic aneurysm admissions in NIS 2019 and about 1,230 ruptured thoracoabdominal and thoracic aneurysm admissions a year (Conroy 2025). |
| Shares applied | EMS share 75% assumed. |
| Arithmetic | 4,625 x 0.75 = 3,470; / 3,401 = 1.0. Low 0.7 uses a 50% EMS share (2,313); high 1.55 adds the 1,230 thoracic ruptures and uses a 90% share (5,270). |
| Check calculation (machine-checked) | 4625 * 0.75 / 3401 |
| Other definition | Prehospital deaths are not represented by admission counts; they cannot all be assigned to the EMS-treated arrest row. |
| Consensus basis | Consensus: door-to-intervention targets for ruptured aneurysm in the Society for Vascular Surgery guideline (Chaikof 2018) [94]. |
| Point and range (per 100,000) | 1.0 (0.7 to 1.6) |
| Counting unit and union coefficients | Counting unit admissions; retention 1.0 in the reference, 0.80 in the lower-count scenario, 0.90 in one sensitivity line. Eligibility 1, 1, 1 (lower-count, reference, higher-count). Intersection with the union of the preceding rows: overlap fraction 0.10, 0.05, 0.00 (lower-count, reference, higher-count). Reference: qualifying 1.00, deduction 0.05, net 0.95 per 100,000. Assumed intersection with preceding nontraumatic arrest. |
| Sources | [95] (data used: NIS 2004-2019); [96]; [94] (data used: Door-to-intervention targets for ruptured aneurysm) |
| Caveats | A US EMS-arrival share was not used; 75% is assumed. The expanded high value adds thoracic aneurysm ruptures. |

### S1.28. Major burn with inhalation injury or airway involvement, EMS-transported (Tier B; assumption-dependent scenario)

| Item | Value |
|---|---|
| Anchor | 29,165 burn admissions in NIS 2020 (American Burn Association fact sheet). |
| Shares applied | Inhalation injury share 7.5% (2.2% to 14% across NIS-based series, Veeravagu 2015; up to 10.3%, Foncerrada 2018); EMS share 85%. |
| Arithmetic | 29,165 x 0.075 x 0.85 = 1,859; / 3,401 = 0.547. Bounds use inhalation shares of 4.4% and 11.5%. |
| Check calculation (machine-checked) | 29165 * 0.075 * 0.85 / 3401 |
| Other definition | Burn admissions without this inhalation or airway marker are outside the row. |
| Consensus basis | Clinical reasoning: early airway control before supraglottic edema, as described in the burn literature (Foncerrada 2018) [97]. |
| Point and range (per 100,000) | 0.5 (0.3 to 0.8) |
| Counting unit and union coefficients | Counting unit patient events or a population rate; retention 1.0 in every scenario. Eligibility 1, 1, 1 (lower-count, reference, higher-count). Intersection with the union of the preceding rows: overlap fraction 0.05, 0.02, 0.00 (lower-count, reference, higher-count). Reference: qualifying 0.55, deduction 0.01, net 0.54 per 100,000. Hospital admissions with inhalation injury and an EMS share; burn patients treated at structure or vehicle fires are also in the fire rows, which carry that deduction because they follow this row in the order. |
| Sources | [98] (data used: 29,165 burn admissions, HCUP NIS 2020); [97] (data used: Inhalation injury in up to 10.3% of burn admissions; supraglottic edema within hours); [99] |
| Caveats | This hospital-based inhalation-injury proxy is broader than an immediately obstructed airway. It may arise from incidents in the separate fire panel, which is not added to the medical sum. |

### S1.29. Ruptured ectopic pregnancy with hemodynamic instability, EMS-attended (Tier B; assumption-dependent scenario)

| Item | Value |
|---|---|
| Anchor | Ectopic pregnancy 1.40% of pregnancies among Medicaid-insured women 2004 to 2008 (Stulberg 2014); 11% with complications in the same population (Stulberg 2016); ruptured ectopic 22.9% of 606 pregnancy-related hemorrhage deaths 2012 to 2019 (Hollier 2024). |
| Shares applied | Unstable share 30% and EMS share 40% assumed. |
| Arithmetic | About 5.8 million pregnancies x 0.014 x 0.11 x 0.30 x 0.40 = 1,070; / 3,401 = 0.3. Low 0.1: 332 ectopic EMS activations in NEMSIS 2018 x 2.676 x 47% high acuity = 418. High 1.0 treats every complicated ectopic pregnancy as unstable (3,570). |
| Check calculation (machine-checked) | 5800000 * 0.014 * 0.11 * 0.30 * 0.40 / 3401 |
| Other definition | All ectopic pregnancies (about 24 per 100,000) are excluded. |
| Consensus basis | Consensus: surgical management of ruptured ectopic pregnancy in the ACOG bulletin (2018) [100]. |
| Point and range (per 100,000) | 0.3 (0.1 to 1.0) |
| Counting unit and union coefficients | Counting unit patient events or a population rate; retention 1.0 in every scenario. Eligibility 1, 1, 1 (lower-count, reference, higher-count). Intersection with the union of the preceding rows: overlap fraction 0.05, 0.02, 0.00 (lower-count, reference, higher-count). Reference: qualifying 0.30, deduction 0.01, net 0.29 per 100,000. Unstable and EMS shares already applied; assumed preceding hemorrhage/arrest intersection. |
| Sources | [101] (data used: 1.40% of pregnancies); [102] (data used: 11% with complications); [103] (data used: Ruptured ectopic pregnancy 22.9% of 606 hemorrhage deaths); [91] (data used: NEMSIS 2018: 107,771 obstetric activations, 3,489 out-of-hospital deliveries, 629 intrapartum or postpartum hemorrhage activations (55% high acuity), about 1,000 hypertensive or eclampsia activations, 332 ectopic pregnancy activations (47% high acuity)); [100] (data used: Surgical management of ruptured ectopic pregnancy) |
| Caveats | The pregnancy denominator, the unstable share, and the EMS share are assumptions. |

#### S1.30. Upper airway obstruction other than foreign body (angioedema, epiglottitis, deep neck infection) requiring airway intervention (Tier B; assumption-dependent scenario)

| Item | Value |
|---|---|
| Anchor | 108,816 angioedema emergency department visits a year 2001 to 2009 (Kelly 2013); intubation within 14 days in 1.30% of propensity-matched angioedema patients without a substance use disorder and 3.50% of those with one, among 147,806 patients in all (Bogart 2025, Tables I and III); 4,063 epiglottitis admissions a year, 13.2% intubated and 3.6% tracheotomy (Shah and Stocks 2010). |
| Shares applied | EMS share 40% for angioedema and 50% for epiglottitis assumed; deep neck infection bounded at 0.1 or less. |
| Arithmetic | 108,816 x 0.013 x 0.40 = 566; 4,063 x 0.168 x 0.50 = 341; total 907; / 3,401 = 0.27, carried as 0.3. Low 0.13 halves the EMS shares (454). High 0.72 uses full EMS shares (1,415 + 683 = 2,098, 0.62) and adds deep neck infections at 0.1. |
| Check calculation (machine-checked) | (108816 * 0.013 * 0.40 + 4063 * (0.132 + 0.036) * 0.50) / 3401 |
| Other definition | None. |
| Consensus basis | Progression to complete airway obstruction motivates urgent airway assessment and intervention. No uniform fatality or timing effect is assigned to the included diagnoses. |
| Point and range (per 100,000) | 0.3 (0.1 to 0.7) |
| Counting unit and union coefficients | Counting unit patient events or a population rate; retention 1.0 in every scenario. Eligibility 1, 1, 1 (lower-count, reference, higher-count). Intersection with the union of the preceding rows: overlap fraction 0.20, 0.10, 0.00 (lower-count, reference, higher-count). Reference: qualifying 0.30, deduction 0.03, net 0.27 per 100,000. Assumed preceding anaphylaxis and respiratory-presentation intersection. |
| Sources | [104] (data used: 108,816 angioedema visits per year, 2001-2009); [105] (data used: Intubation within 14 days in 1.30% of the propensity-matched cohort without substance use disorder and 3.50% of the cohort with one (Table III); 147,806 patients in Table I); [106] (data used: 4,063 admissions per year; 13.2% intubated; 3.6% tracheotomy; mortality 0.89%) |
| Caveats | Treated epiglottitis mortality 0.89% (Shah and Stocks 2010). |

### S2. Fire and rescue derivation

Fire and rescue rows were counted from NERIS Public, the record-level public feed of the National Emergency Response Information System [107]. Aggregate counts were queried for the most recent complete twelve months of incidents; no record-level data were extracted. Counts were scaled to NFPA national totals by incident share; no completeness adjustment was applied. NERIS incident types are the reporting department's classification of the incident, and counts by incident type are described that way throughout. Two cross-checks sit beside the NERIS rows: the NFIRS 2024 public data release, counted independently (S2.7), and, for the fire rows, NFPA casualty counts with the USFA serious-injury share (S2.6). The block enters the sum after the medical rows in the ordered union (S3.6), so each row also carries an assumed intersection with the medical rows before it.

#### S2.1. NERIS provenance

##### Table S3. NERIS Public provenance

| Item | Value |
|---|---|
| Source | NERIS Public Incident Basics, ArcGIS feature service, layer 0 |
| Feature service item id | 241c7985c3b14da39ae67d2925a677e0 |
| Query date | 4 September 2026 |
| Window | call_create >= 2025-08-01 AND call_create < 2026-08-01 (most recent complete 12 months) |
| Records in the service at query time | 17,047,939 (call_create 2025-04-07 to 2026-08-28) |
| Records in the window | 16,997,003 |
| Departments reporting in the window | 17,103 |
| Department type mix, reporting departments | career 2,242; combination 3,623; volunteer 9,307; type not recorded 1,922 (sum 17,094; 9 departments carry another or unparsed type value) |

| Item | Value |
|---|---|
| Method | Aggregate outStatistics (count grouped by type_1/type_2/type_3 and by neris_id_dept) and returnCountOnly queries, chunked by ObjectId in 500,000-record blocks. No record-level export. |
| Departments with population_protected greater than 0 | 6,168 |
| Sum of population_protected, those departments | 169,866,192 |
| Incidents from those departments | 7,764,767 |

NERIS has a population_protected field for departments, and 6,168 of the 17,103 reporting departments carry a value above zero. Those departments reported 7,764,767 incidents against a summed population of 169,866,192, which is 4,571 per 100,000, against a national rate of about 12,083. Jurisdictions overlap and reporting is partial, so population_protected was not used as a denominator. Incident share scales the counts instead (S2.3). No national comparator is given for the department type mix because the NFPA department profile counts departments on a different basis from the NERIS reporting-department field.

### S2.2. Incident types included and the tactic filter

An incident entered a fire or rescue row when its type_1 value placed a person in immediate danger and its action_and_tactics field contained the token EMERGENCY_MEDICAL_CARE, which we took as evidence that a patient was present and treated. The filter matched that token in the action_and_tactics field on incidents selected by type_1; the search filter used for the context rows matched any search tactic token. Table S4 lists the type_1 values feeding each row, the number of type_1 incidents in the window (the denominator), and the number carrying the tactic. The motor vehicle crash row is built from extrications performed and a published entrapment share (S2.5); the type_1 and tactic counts shown for it here set its upper bound. Carbon monoxide incidents were counted on the medical side and are shown as context in Table S10.

**Table S4. Type_1 values counted in each fire and rescue row**

| Row | type_1 values | type_1 incidents | With emergency medical care tactic | Share |
|---|---|---:|---:|---:|
| Motor vehicle crash with a physically trapped patient | RESCUE / TRANSPORTATION / MOTOR_VEHICLE_EXTRICATION_ENTRAPPED | 42,637 | 12,659 | 29.7% |
| Structure fire with a patient treated | FIRE / STRUCTURE_FIRE / STRUCTURAL_INVOLVEMENT_FIRE; FIRE / STRUCTURE_FIRE / CONFINED_COOKING_APPLIANCE_FIRE; FIRE / STRUCTURE_FIRE / ROOM_AND_CONTENTS_FIRE; FIRE / STRUCTURE_FIRE / CHIMNEY_FIRE | 229,796 | 3,351 | 1.5% |
| Other extrication or entrapment (structure, machinery, outside) with a patient treated | RESCUE / OUTSIDE / EXTRICATION_ENTRAPPED; RESCUE / STRUCTURE / EXTRICATION_ENTRAPPED | 11,899 | 2,194 | 18.4% |
| Vehicle fire with a patient treated | FIRE / TRANSPORTATION_FIRE / VEHICLE_FIRE_PASSENGER; FIRE / TRANSPORTATION_FIRE / VEHICLE_FIRE_COMMERCIAL; FIRE / TRANSPORTATION_FIRE / VEHICLE_FIRE_RV; FIRE / TRANSPORTATION_FIRE / POWERED_MOBILITY_DEVICE_FIRE; FIRE / TRANSPORTATION_FIRE / AIRCRAFT_FIRE; FIRE / TRANSPORTATION_FIRE / TRAIN_RAIL_FIRE; FIRE / TRANSPORTATION_FIRE / BOAT_PERSONAL_WATERCRAFT_BARGE_FIRE; FIRE / TRANSPORTATION_FIRE / VEHICLE_FIRE_FOOD_TRUCK | 82,950 | 1,095 | 1.3% |
| Person in water (standing or swift) with a patient treated | RESCUE / WATER / PERSON_IN_WATER_STANDING; RESCUE / WATER / PERSON_IN_WATER_SWIFTWATER | 10,973 | 1,028 | 9.4% |
| Backcountry rescue with a patient treated | RESCUE / OUTSIDE / BACKCOUNTRY_RESCUE | 2,617 | 747 | 28.5% |

| Row | type_1 values | type_1 incidents | With emergency medical care tactic | Share |
|---|---|---:|---:|---:|
| Rope rescue (high, steep, low angle) with a patient treated | RESCUE / OUTSIDE / HIGH_ANGLE_RESCUE; RESCUE / OUTSIDE / LOW_ANGLE_RESCUE; RESCUE / OUTSIDE / STEEP_ANGLE_RESCUE | 2,427 | 506 | 20.8% |
| Aviation crash with a patient treated | RESCUE / TRANSPORTATION / AVIATION_COLLISION_CRASH | 1,340 | 274 | 20.4% |
| Hazardous materials release with a patient treated | HAZSIT / HAZARDOUS_MATERIALS / HAZMAT_RELEASE_TRANSPORT; HAZSIT / HAZARDOUS_MATERIALS / HAZMAT_RELEASE_FACILITY | 5,944 | 218 | 3.7% |
| Structural collapse with a patient treated | RESCUE / STRUCTURE / BUILDING_STRUCTURE_COLLAPSE | 2,959 | 194 | 6.6% |
| Watercraft in distress with a patient treated | RESCUE / WATER / WATERCRAFT_IN_DISTRESS | 3,765 | 157 | 4.2% |
| Confined space or trench rescue with a patient treated | RESCUE / OUTSIDE / CONFINED_SPACE_RESCUE; RESCUE / STRUCTURE / CONFINED_SPACE_RESCUE; RESCUE / OUTSIDE / TRENCH | 1,243 | 130 | 10.5% |
| Explosion or overpressure rupture with a patient treated | FIRE / SPECIAL_FIRE / EXPLOSION; HAZSIT / OVERPRESSURE / RUPTURE_WITHOUT_FIRE | 4,194 | 130 | 3.1% |

### Table S5. All 85 type_1 values in the window, sorted by count

| Rank | Category | Group | Type | Incidents | Share of records |
|---:|---|---|---|---:|---:|
| 1 | MEDICAL | | | 9,740,467 | 57.31% |
| 2 | NOEMERG | CANCELLED | | 1,160,474 | 6.83% |
| 3 | MEDICAL | INJURY | MOTOR_VEHICLE_COLLISION | 728,136 | 4.28% |
| 4 | PUBSERV | CITIZEN_ASSIST | LIFT_ASSIST | 685,879 | 4.04% |
| 5 | PUBSERV | CITIZEN_ASSIST | CITIZEN_ASSIST_SERVICE_CALL | 513,846 | 3.02% |
| 6 | PUBSERV | ALARMS_NONMED | FIRE_ALARM | 503,897 | 2.96% |
| 7 | NOEMERG | FALSE_ALARM | ACCIDENTAL_ALARM | 449,649 | 2.65% |
| 8 | NOEMERG | GOOD_INTENT | NO_INCIDENT_FOUND_LOCATION_ERROR | 349,479 | 2.06% |
| 9 | NOEMERG | FALSE_ALARM | MALFUNCTIONING_ALARM | 339,083 | 1.99% |
| 10 | HAZSIT | HAZARD_NONCHEM | MOTOR_VEHICLE_COLLISION | 240,868 | 1.42% |
| 11 | HAZSIT | HAZARDOUS_MATERIALS | GAS_LEAK_ODOR | 168,279 | 0.99% |
| 12 | FIRE | OUTSIDE_FIRE | VEGETATION_GRASS_FIRE | 150,711 | 0.89% |
| 13 | HAZSIT | HAZARD_NONCHEM | ELEC_POWER_LINE_DOWN_ARCHING_MALFUNC | 145,605 | 0.86% |
| 14 | FIRE | STRUCTURE_FIRE | STRUCTURAL_INVOLVEMENT_FIRE | 120,823 | 0.71% |
| 15 | NOEMERG | FALSE_ALARM | OTHER_FALSE_CALL | 115,179 | 0.68% |
| 16 | FIRE | OUTSIDE_FIRE | TRASH_RUBBISH_FIRE | 111,954 | 0.66% |
| 17 | HAZSIT | INVESTIGATION | SMOKE_INVESTIGATION | 108,262 | 0.64% |
| 18 | HAZSIT | INVESTIGATION | ODOR | 96,538 | 0.57% |
| 19 | PUBSERV | ALARMS_NONMED | CO_ALARM | 84,840 | 0.50% |
| 20 | FIRE | OUTSIDE_FIRE | OTHER_OUTSIDE_FIRE | 82,231 | 0.48% |
| 21 | RESCUE | STRUCTURE | ELEVATOR_ESCALATOR_RESCUE | 78,318 | 0.46% |
| 22 | LAWENFORCE | | | 73,873 | 0.43% |
| 23 | NOEMERG | GOOD_INTENT | SMOKE_FROM_NONHOSTILE_SOURCE | 72,167 | 0.42% |
| 24 | FIRE | STRUCTURE_FIRE | CONFINED_COOKING_APPLIANCE_FIRE | 66,516 | 0.39% |
| 25 | PUBSERV | OTHER | STANDBY | 66,234 | 0.39% |
| 26 | FIRE | TRANSPORTATION_FIRE | VEHICLE_FIRE_PASSENGER | 58,685 | 0.35% |
| 27 | PUBSERV | ALARMS_NONMED | OTHER_ALARM | 49,544 | 0.29% |
| 28 | NOEMERG | GOOD_INTENT | INVESTIGATE_HAZARDOUS_RELEASE | 47,904 | 0.28% |

| Rank | Category | Group | Type | Incidents | Share of records |
|---:|---|---|---|---:|---:|
| 29 | NOEMERG | FALSE_ALARM | INTENTIONAL_FALSE_ALARM | 47,643 | 0.28% |
| 30 | HAZSIT | HAZARD_NONCHEM | ELEC_HAZARD_SHORT_CIRCUIT | 47,330 | 0.28% |
| 31 | PUBSERV | CITIZEN_ASSIST | PERSON_IN_DISTRESS | 45,153 | 0.27% |
| 32 | RESCUE | TRANSPORTATION | MOTOR_VEHICLE_EXTRICATION_ENTRAPPED | 42,637 | 0.25% |
| 33 | NOEMERG | GOOD_INTENT | CONTROLLED_BURNING_AUTHORIZED | 41,077 | 0.24% |
| 34 | PUBSERV | DISASTER_WEATHER | WEATHER_RESPONSE | 36,422 | 0.21% |
| 35 | HAZSIT | HAZARDOUS_MATERIALS | CARBON_MONOXIDE_RELEASE | 34,472 | 0.20% |
| 36 | FIRE | STRUCTURE_FIRE | ROOM_AND_CONTENTS_FIRE | 33,558 | 0.20% |
| 37 | PUBSERV | OTHER | MOVE_UP | 29,279 | 0.17% |
| 38 | HAZSIT | HAZARDOUS_MATERIALS | FUEL_SPILL_ODOR | 23,622 | 0.14% |
| 39 | FIRE | OUTSIDE_FIRE | WILDFIRE_WILDLAND | 21,945 | 0.13% |
| 40 | FIRE | OUTSIDE_FIRE | DUMPSTER_OUTDOOR_CONTAINER_FIRE | 20,984 | 0.12% |
| 41 | PUBSERV | DISASTER_WEATHER | DAMAGE_ASSESSMENT | 17,461 | 0.10% |
| 42 | FIRE | TRANSPORTATION_FIRE | VEHICLE_FIRE_COMMERCIAL | 15,137 | 0.09% |
| 43 | RESCUE | TRANSPORTATION | AVIATION_STANDBY | 12,885 | 0.08% |
| 44 | FIRE | OUTSIDE_FIRE | UTILITY_INFRASTRUCTURE_FIRE | 11,292 | 0.07% |
| 45 | FIRE | STRUCTURE_FIRE | CHIMNEY_FIRE | 8,899 | 0.05% |
| 46 | RESCUE | OUTSIDE | EXTRICATION_ENTRAPPED | 8,631 | 0.05% |
| 47 | PUBSERV | ALARMS_NONMED | GAS_ALARM | 6,907 | 0.04% |
| 48 | RESCUE | WATER | PERSON_IN_WATER_STANDING | 6,595 | 0.04% |
| 49 | PUBSERV | CITIZEN_ASSIST | LOST_PERSON | 6,499 | 0.04% |
| 50 | PUBSERV | OTHER | DAMAGED_HYDRANT | 6,298 | 0.04% |
| 51 | FIRE | OUTSIDE_FIRE | CONSTRUCTION_WASTE | 6,060 | 0.04% |
| 52 | FIRE | OUTSIDE_FIRE | WILDFIRE_URBAN_INTERFACE | 5,176 | 0.03% |
| 53 | FIRE | TRANSPORTATION_FIRE | VEHICLE_FIRE_RV | 4,624 | 0.03% |
| 54 | RESCUE | WATER | PERSON_IN_WATER_SWIFTWATER | 4,378 | 0.03% |
| 55 | RESCUE | WATER | WATERCRAFT_IN_DISTRESS | 3,765 | 0.02% |
| 56 | RESCUE | STRUCTURE | EXTRICATION_ENTRAPPED | 3,268 | 0.02% |
| 57 | HAZSIT | HAZARDOUS_MATERIALS | HAZMAT_RELEASE_TRANSPORT | 3,152 | 0.02% |
| 58 | RESCUE | STRUCTURE | BUILDING_STRUCTURE_COLLAPSE | 2,959 | 0.02% |
| 59 | HAZSIT | HAZARDOUS_MATERIALS | HAZMAT_RELEASE_FACILITY | 2,792 | 0.02% |
| 60 | HAZSIT | OVERPRESSURE | RUPTURE_WITHOUT_FIRE | 2,774 | 0.02% |
| 61 | RESCUE | OUTSIDE | BACKCOUNTRY_RESCUE | 2,617 | 0.02% |
| 62 | HAZSIT | HAZARD_NONCHEM | BOMB_THREAT_RESPONSE_SUSPICIOUS_PACKAGE | 2,174 | 0.01% |
| 63 | RESCUE | OUTSIDE | LIMITED_NO_ACCESS | 1,590 | 0.01% |
| 64 | FIRE | SPECIAL_FIRE | EXPLOSION | 1,420 | 0.01% |
| 65 | RESCUE | TRANSPORTATION | AVIATION_COLLISION_CRASH | 1,340 | 0.01% |
| 66 | FIRE | TRANSPORTATION_FIRE | POWERED_MOBILITY_DEVICE_FIRE | 1,299 | 0.01% |
| 67 | RESCUE | OUTSIDE | HIGH_ANGLE_RESCUE | 1,165 | 0.01% |
| 68 | FIRE | OUTSIDE_FIRE | OUTSIDE_TANK_FIRE | 1,158 | 0.01% |
| 69 | HAZSIT | HAZARDOUS_MATERIALS | BIOLOGICAL_RELEASE_INCIDENT | 1,132 | 0.01% |
| 70 | HAZSIT | OVERPRESSURE | NO_RUPTURE | 1,110 | 0.01% |
| 71 | FIRE | TRANSPORTATION_FIRE | AIRCRAFT_FIRE | 994 | 0.01% |
| 72 | FIRE | TRANSPORTATION_FIRE | TRAIN_RAIL_FIRE | 972 | 0.01% |
| 73 | FIRE | SPECIAL_FIRE | ESS_FIRE | 938 | 0.01% |
| 74 | FIRE | TRANSPORTATION_FIRE | BOAT_PERSONAL_WATERCRAFT_BARGE_FIRE | 868 | 0.01% |
| 75 | RESCUE | OUTSIDE | LOW_ANGLE_RESCUE | 797 | 0.00% |

| Rank | Category | Group | Type | Incidents | Share of records |
|---|---|---|---|---|---|
| 76 | FIRE | SPECIAL_FIRE | INFRASTRUCTURE_FIRE | 787 | 0.00% |
| 77 | RESCUE | OUTSIDE | CONFINED_SPACE_RESCUE | 766 | 0.00% |
| 78 | NOEMERG | FALSE_ALARM | BOMB_SCARE | 738 | 0.00% |
| 79 | RESCUE | TRANSPORTATION | TRAIN_RAIL_COLLISION_DERAILMENT | 490 | 0.00% |
| 80 | RESCUE | OUTSIDE | STEEP_ANGLE_RESCUE | 465 | 0.00% |
| 81 | FIRE | TRANSPORTATION_FIRE | VEHICLE_FIRE_FOOD_TRUCK | 371 | 0.00% |
| 82 | RESCUE | STRUCTURE | CONFINED_SPACE_RESCUE | 291 | 0.00% |
| 83 | RESCUE | OUTSIDE | TRENCH | 186 | 0.00% |
| 84 | NULL | | | 138 | 0.00% |
| 85 | HAZSIT | HAZARDOUS_MATERIALS | RADIOACTIVE_RELEASE_INCIDENT | 82 | 0.00% |
| | Total | | | 16,997,003 | 100.00% |

#### Table S6. Incident counts by type token in any type position

Each count sums the token's appearances in type_1, type_2, and type_3; an incident can carry up to three types. The ledger treats the sum as a count of incidents, and an incident that carried the same token in two positions would be counted twice.

| Type token | Incidents (type_1 + type_2 + type_3) |
|---|---|
| MOTOR_VEHICLE_EXTRICATION_ENTRAPPED | 45,517 |
| PERSON_IN_WATER | 11,586 |
| WATERCRAFT_IN_DISTRESS | 3,989 |
| OUTSIDE_EXTRICATION_ENTRAPPED | 10,166 |
| STRUCTURE_EXTRICATION_ENTRAPPED | 3,954 |
| BUILDING_STRUCTURE_COLLAPSE | 3,473 |
| CONFINED_SPACE_RESCUE | 1,156 |
| TRENCH | 202 |
| ROPE_RESCUE_HIGH_STEEP_LOW_ANGLE | 2,940 |
| BACKCOUNTRY_RESCUE | 2,966 |
| LIMITED_NO_ACCESS | 1,964 |
| CARBON_MONOXIDE_RELEASE | 35,776 |
| EXPLOSION | 1,636 |
| RUPTURE_WITHOUT_FIRE | 2,969 |
| AVIATION_COLLISION_CRASH | 1,426 |
| TRAIN_RAIL_COLLISION_DERAILMENT | 539 |
| STRUCTURE_FIRE | 239,889 |
| HAZMAT_RELEASE | 6,915 |

#### Table S7. Tactic counts on the type_1 basis

| Key | Incidents |
|---|---|
| structure_fires_all | 229,796 |
| structure_fires_search | 17,680 |
| structure_fires_emc | 3,351 |
| structure_fires_search_or_emc | 20,345 |
| structure_fires_displacement_gt0 | 23,697 |
| structure_fires_location_use_residential | 116,943 |
| vehicle_fires_all | 82,950 |
| vehicle_fires_emc | 1,095 |
| vehicle_fires_with_extrication_type2or3 | 90 |
| co_release_emc | 783 |

| Key | Incidents |
|---|---|
| mvc_extrication_entrapped_emc | 12,659 |
| person_in_water_emc | 1,028 |
| collapse_emc | 194 |
| confined_space_emc | 90 |
| trench_emc | 40 |
| rope_rescue_emc | 506 |
| backcountry_emc | 747 |
| outside_extrication_entrapped_emc | 1,820 |
| structure_extrication_entrapped_emc | 374 |
| explosion_emc | 96 |
| rupture_without_fire_emc | 34 |
| hazmat_release_emc | 218 |
| watercraft_in_distress_emc | 157 |
| aviation_crash_emc | 274 |
| structure_fires_residential_search | 11,099 |
| medical_type1_emc_check | 6,870,747 |

The emergency-medical-care tactic was present on 6,870,747 of 9,740,467 incidents typed MEDICAL (70.5%). Tactic-based patient counts on other types may therefore undercount, and no adjustment is applied for it: whether crews record the tactic at the same rate at a structure fire as on a medical call is not known. The NFIRS cross-check, which uses a different patient marker, is the check on the undercount.

### S2.3. Scaling factors

NERIS participation is incomplete, so counts were scaled to NFPA 2024 national totals by incident share [108,109]. Two factors bracket the scaling: the structure-fire share and the all-incident share. The point estimate uses their midpoint and the range spans both. NFPA's totals are for calendar 2024 and the NERIS window is 1 August 2025 to 31 July 2026; the scaling assumes that national incident volume was similar in the two periods.

**Table S8. Scaling factors**

| Factor | Arithmetic | Value |
|---|---|---|
| Structure-fire share (low) | 470,500 NFPA structure fires / 229,796 NERIS structure fires | 2.047 |
| All-incident share (high) | 41,095,000 NFPA incidents net of mutual aid / 16,997,003 NERIS incidents | 2.418 |
| Midpoint (point estimate) | (2.047 + 2.418) / 2 | 2.233 |
| Emergency-medical-care tactic share on MEDICAL incidents (reported, not applied) | 6,870,747 / 9,740,467 | 70.5% |
| Per 100,000 | scaled count / 3,401 | |

The factors are printed rounded (2.047, 2.418, 2.233) here and in the arithmetic cells below; the stored values are unrounded, so a product recomputed from the printed factors can differ from the stored value in the second decimal.

### S2.4. Fire and rescue rows

The group is fire and rescue events with a patient treated or trapped. Twelve of the thirteen rows count a patient treated at an incident whose type places a person in immediate danger. The motor vehicle crash row counts physically trapped crash occupants and is built differently (S2.5). Table S9 gives every row with the arithmetic from the ledger and the intersection fraction the ordered union applies to it (S3.6).

**Table S9. Fire and rescue events with a patient treated or trapped, NERIS Public 1 August 2025 to 31 July 2026, scaled**

| Row | NERIS incidents (12 months) | Arithmetic | Per 100,000 | Range | Overlap fraction (lower, reference, higher) | Net after overlap | Incidence basis | Sources | Notes |
|---|---|---|---|---|---|---|---|---|---|
| Motor vehicle crash with a physically trapped patient | 12,659 | Extrications (15.6 + 29.9) / 2 = 22.7; x 0.10 = 2.27. Low 15.6 x 0.10 = 1.56. High 12,659 x 2.418 / 3,401 = 9.00 (every typed incident with a patient treated, all-incident scaling). | 2.3 | 1.6 to 9.0 | 0.75, 0.50, 0.00 | 1.14 | Assumption-dependent scenario | [107,110,111] | A patient treated at an incident typed as extrication is not shown to have been trapped, and NFIRS and NERIS record a patient on 28% and 30% of their extrication incidents, so the treated-patient counts measure typing breadth, not entrapment. Overlaps the blunt trauma and TBI rows; the union deduction carries that. |
| Structure fire with a patient treated | 3,351 | 3,351 x 2.233 / 3,401 = 2.20; low 3,351 x 2.047 / 3,401 = 2.02; high 3,351 x 2.418 / 3,401 = 2.38. | 2.2 | 2.0 to 2.4 | 0.20, 0.10, 0.00 | 1.98 | US operational count, scaled by incident share | [107,109,111,112] | Cross-checks: NFIRS 2024 structure fires with a civilian casualty, 4,942 x 1.485 / 3,401 = 2.2 per 100,000; NFPA 2024 structure-fire casualties (3,170 deaths, 10,335 injuries) with the USFA serious-injury share of 16%, (3,170 + 10,335 x 0.16) / 3,401 = 1.4. |

| Row | NERIS incidents (12 months) | Arithmetic | Per 100,000 | Range | Overlap fraction (lower, reference, higher) | Net after overlap | Incidence basis | Sources | Notes |
|---|---|---|---|---|---|---|---|---|---|
| Other extrication or entrapment (structure, machinery, outside) with a patient treated | 2,194 | 2,194 x 2.233 / 3,401 = 1.44; low 2,194 x 2.047 / 3,401 = 1.32; high 2,194 x 2.418 / 3,401 = 1.56. | 1.4 | 1.3 to 1.6 | 0.50, 0.25, 0.00 | 1.08 | US operational count, scaled by incident share | [107,111] | |
| Vehicle fire with a patient treated | 1,095 | 1,095 x 2.233 / 3,401 = 0.72; low 1,095 x 2.047 / 3,401 = 0.66; high 1,095 x 2.418 / 3,401 = 0.78. | 0.7 | 0.7 to 0.8 | 0.20, 0.10, 0.00 | 0.65 | US operational count, scaled by incident share | [107,109,111,112,113] | Cross-checks: NFIRS 2024 vehicle fires with an EMS action, 1,778 x 1.439 / 3,401 = 0.75; NFPA 2024 vehicle-fire casualties (510 deaths, 1,070 injuries) at the 16% serious share, 0.20. |
| Person in water (standing or swift) with a patient treated | 1,028 | 1,028 x 2.233 / 3,401 = 0.67; low 1,028 x 2.047 / 3,401 = 0.62; high 1,028 x 2.418 / 3,401 = 0.73. | 0.7 | 0.6 to 0.7 | 0.75, 0.50, 0.00 | 0.34 | US operational count, scaled by incident share | [107,111] | Incidents typed person in water scale to 7.0 to 8.2 per 100,000; NFIRS 2024 water rescues with an EMS action 0.73. |
| Backcountry rescue with a patient treated | 747 | 747 x 2.233 / 3,401 = 0.49; low 747 x 2.047 / 3,401 = 0.45; high 747 x 2.418 / 3,401 = 0.53. | 0.5 | 0.4 to 0.5 | 0.50, 0.25, 0.00 | 0.37 | US operational count, scaled by incident share | [107] | |
| Rope rescue (high, steep, low angle) with a patient treated | 506 | 506 x 2.233 / 3,401 = 0.33; low 506 x 2.047 / 3,401 = 0.30; high 506 x 2.418 / 3,401 = 0.36. | 0.3 | 0.3 to 0.4 | 0.50, 0.25, 0.00 | 0.25 | US operational count, scaled by incident share | [107] | |

| Row | NERIS incidents (12 months) | Arithmetic | Per 100,000 | Range | Overlap fraction (lower, reference, higher) | Net after overlap | Incidence basis | Sources | Notes |
|---|---|---|---|---|---|---|---|---|---|
| Aviation crash with a patient treated | 274 | 274 x 2.233 / 3,401 = 0.18; low 274 x 2.047 / 3,401 = 0.16; high 274 x 2.418 / 3,401 = 0.19. | 0.2 | 0.2 to 0.2 | 0.50, 0.25, 0.00 | 0.13 | US operational count, scaled by incident share | [107] | |
| Hazardous materials release with a patient treated | 218 | 218 x 2.233 / 3,401 = 0.14; low 218 x 2.047 / 3,401 = 0.13; high 218 x 2.418 / 3,401 = 0.15. | 0.1 | 0.1 to 0.2 | 0.50, 0.25, 0.00 | 0.11 | US operational count, scaled by incident share | [107] | |
| Structural collapse with a patient treated | 194 | 194 x 2.233 / 3,401 = 0.13; low 194 x 2.047 / 3,401 = 0.12; high 194 x 2.418 / 3,401 = 0.14. | 0.1 | 0.1 to 0.1 | 0.50, 0.25, 0.00 | 0.10 | US operational count, scaled by incident share | [107] | |
| Watercraft in distress with a patient treated | 157 | 157 x 2.233 / 3,401 = 0.10; low 157 x 2.047 / 3,401 = 0.09; high 157 x 2.418 / 3,401 = 0.11. | 0.1 | 0.1 to 0.1 | 0.50, 0.25, 0.00 | 0.08 | US operational count, scaled by incident share | [107] | |
| Confined space or trench rescue with a patient treated | 130 | 130 x 2.233 / 3,401 = 0.09; low 130 x 2.047 / 3,401 = 0.08; high 130 x 2.418 / 3,401 = 0.09. | 0.1 | 0.1 to 0.1 | 0.50, 0.25, 0.00 | 0.06 | US operational count, scaled by incident share | [107] | |
| Explosion or overpressure rupture with a patient treated | 130 | 130 x 2.233 / 3,401 = 0.09; low 130 x 2.047 / 3,401 = 0.08; high 130 x 2.418 / 3,401 = 0.09. | 0.1 | 0.1 to 0.1 | 0.50, 0.25, 0.00 | 0.06 | US operational count, scaled by incident share | [107] | |
| Subtotal | 22,683 | | 9 | 8 to 16 | | 6.3 | | | |

The NERIS count shown for the motor vehicle crash row (12,659) is the emergency-medical-care count that sets its high value; its point is built from extrications performed and the physical-entrapment share (S2.5). The unrounded gross subtotal is 8.9 (7.6 to 16.1), rounded half up; after the union deductions the block adds 6.3 (5.6 to 10.1). The overlap fractions are the assumed intersection of each fire and rescue row with the union of the preceding medical rows (burn, trauma, arrest, drowning). Vehicle entrapment and water rescue carry the largest fractions because trapped patients in shock or with head injury are in the trauma rows and rescued swimmers who reach an emergency department are in the drowning row. The reference takes half of those two rows as already counted and a quarter of the other rescue rows; the lower-count set takes three quarters and a half; the higher-count set takes none, as for the medical rows.

#### S2.5. The motor vehicle crash row

The motor vehicle crash row is built differently from the other twelve, and it counts physically trapped crash occupants. Extrications performed per 100,000 residents lie between two readings. NFIRS 2024 extrication actions recorded on motor vehicle collision incidents (types 322 to 324) or vehicle extrication incidents (type 352) give 40,853 x 1.296 / 3,401 = 15.6; action codes are optional, so this is a floor [111]. NERIS incidents typed as extrication or entrapment in any type position give 45,517 x 2.233 / 3,401 = 29.9; typing precedes arrival, so this is a ceiling [107]. The point takes the central value, (15.6 + 29.9) / 2 = 22.7, and applies the physical-entrapment share from the Nutbeam scoping review, a share of extricated patients (about one in ten is physically trapped): 22.7 x 0.10 = 2.27, displayed as 2.3 [110]. The low value applies the share to the floor, 15.6 x 0.10 = 1.56. The high value is every typed incident with a patient treated at the all-incident scaling: 12,659 x 2.418 / 3,401 = 9.00, displayed as 9.0. The Nutbeam share is a clinical share borrowed from the UK and European extrication literature; the paper's transfer rule allows it because both incidence readings are US counts and the row says what was borrowed and why.

A patient treated at an incident typed as extrication is not shown to have been trapped. NFIRS records an EMS action on 6,646 of its 23,936 type 352 incidents (28%) and NERIS records emergency medical care on 12,659 of its 42,637 incidents typed extrication or entrapment in the first position (30%), so the two systems agree on the treated-patient share and their treated-patient counts measure typing breadth: NFIRS types 9.1 per 100,000 as extrication and NERIS 27 to 32 (Table S10). Counting every NERIS typed incident with a patient treated, without the entrapment share, gives 8.3 per 100,000; the main text's sensitivity table carries that reading as a variant (593). The row's incidence basis is assumption-dependent because its point rests on a central value between two readings and a borrowed share. It overlaps the blunt trauma and TBI rows, and the ordered union deducts 50% of it in the reference (S3.6).

**Table S10. Context rows, reported and not counted**

| Row | NERIS incidents | Basis | Arithmetic | Per 100,000 (low to high) | Notes |
|---|---|---|---|---|---|
| Structure fires, all (first type position) | 229,796 | type_1 = structure fire (first type position), the count the structure-fire scaling factor is built on | low 229,796 x 2.047 / 3,401 = 138.34; high 229,796 x 2.418 / 3,401 = 163.36. | 138.3 to 163.4 | NFPA 2024: 470,500 = 138 per 100,000; the low bound reproduces it by construction |
| Structure fires with a search tactic and no patient treated | 16,994 | type_1 structure fires with a search tactic token and without the emergency-medical-care token (20,345 with either token minus 3,351 with the medical token) | low 16,994 x 2.047 / 3,401 = 10.23; high 16,994 x 2.418 / 3,401 = 12.08. | 10.2 to 12.1 | |
| Structure fires with occupants displaced (occupied proxy) | 23,697 | type_1 structure fires with displacement_count greater than 0 (structure_fires_displacement_gt0) | low 23,697 x 2.047 / 3,401 = 14.27; high 23,697 x 2.418 / 3,401 = 16.85. | 14.3 to 16.8 | |
| Motor vehicle extrication or entrapment, by incident type | 45,517 | MOTOR_VEHICLE_EXTRICATION_ENTRAPPED in any type position (Table S6) | low 45,517 x 2.047 / 3,401 = 27.40; high 45,517 x 2.418 / 3,401 = 32.36. | 27.4 to 32.4 | |
| Carbon monoxide incidents with CO found, by incident type (CO alarms excluded) | 35,776 | CARBON_MONOXIDE_RELEASE in any type position (Table S6); the PUBSERV CO_ALARM type is a separate value and is excluded | low 35,776 x 2.047 / 3,401 = 21.54; high 35,776 x 2.418 / 3,401 = 25.43. | 21.5 to 25.4 | NFPA 2010 estimate 80,100 = 23.6 per 100,000 on the 2024 population, 25.9 on the 2010 population |

| Row | NERIS incidents | Basis | Arithmetic | Per 100,000 (low to high) | Notes |
|---|---|---|---|---|---|
| Carbon monoxide incidents with a patient treated | 783 | type_1 CARBON_MONOXIDE_RELEASE with the emergency-medical-care tactic (co_release_emc) | low 783 x 2.047 / 3,401 = 0.47; high 783 x 2.418 / 3,401 = 0.56. | 0.5 to 0.6 | Emergency-medical-care tactic recorded on 2.3% of CO release incidents; not counted here because carbon monoxide poisoning is counted once, on the medical side (Table 1). |

The two incident-share factors scale the context rows by incident type. Both carbon monoxide rows are shown because the ledger counts carbon monoxide poisoning once, on the medical side (S1): the tactic appears on 2.3% of type_1 carbon monoxide incidents (783 of 34,472). The search context row excludes the 686 structure fires with both a search and a patient treated, which are in the counted structure fire row.

### S2.6. Cross-checks on the fire rows

Table S11 sets each observed fire and rescue event beside its NFIRS analogue and, for the fire rows, beside an estimate built from NFPA casualty counts. The three readings use different patient markers (a tactic token in NERIS, a casualty field or an action code in NFIRS, a person counted as a casualty in NFPA's survey), so agreement within a factor of two is the most the comparison can show.

**Table S11. Fire and rescue operational events per 100,000: NERIS rows, NFIRS analogues, and NFPA casualty-based estimates**

| Event | NERIS incidents (12 months) | NERIS scaled per 100,000 (low to high) | NFIRS 2024 marker and count | NFIRS scaled per 100,000 | NFPA casualties x 16% serious | In the sum as |
|---|---|---|---|---|---|---|
| Structure fire with a patient treated | 3,351 | 2.20 (2.02 to 2.38) | structure fires with a civilian casualty, 4,942 | 2.16 | 1.42 | f_structure |
| Vehicle fire with a patient treated | 1,095 | 0.72 (0.66 to 0.78) | vehicle fires with an EMS action, 1,778 | 0.75 | 0.20 | f_vehicle |
| Motor vehicle extrication or entrapment with a patient treated | 12,659 | 8.31 (7.62 to 9.00) | vehicle extrication (352) with an EMS action, 6,646 | 2.53 | | f_mvc (10% physically trapped) |
| Extrication actions performed at collisions or extrications | 45,517 | 29.88 (27.40 to 32.36) | extrication action (23) on 322 to 324 or 352 incidents, 40,853 | 15.56 | | f_mvc reading (NERIS typed, any position) |
| Water rescue with a patient treated | 1,028 | 0.67 (0.62 to 0.73) | water rescue (360 to 365) with an EMS action, 1,908 | 0.73 | | f_water |
| Carbon monoxide incident with a patient treated | 783 | 0.51 (0.47 to 0.56) | carbon monoxide incidents (424) with an EMS action, 1,316 | 0.50 | | counted on the medical side (co, Table 1) |

**Casualties from NFPA counts.** NFPA reports 3,920 civilian fire deaths and 11,780 civilian fire injuries in 2024, of which 3,170 deaths and 10,335 injuries were in structure fires and 510 and 1,070 in vehicle fires [109]. USFA's residential topical report classifies 16% of civilian fire injuries as serious or life-threatening [112]. Counting every death and 16% of injuries as a time-critical patient gives (3,170 + 10,335 x 0.16) / 3,401 = 1.42 per 100,000 for structure fires and (510 + 1,070 x 0.16) / 3,401 = 0.20 for vehicle fires. The USFA share is residential and is transferred to all fires; it is a borrowed clinical share and serves here as a cross-check only. The structure-fire figure sits below the NERIS row (2.2, 2.0 to 2.4) and the NFIRS count of structure fires with a civilian casualty (2.16); a patient treated at a fire is a broader marker than a casualty counted by the department, and every fire death in the NFPA count is an attended incident with a dead victim, so the casualty-based estimate is the narrower reading.

**Casualty fires from NFPA counts.** A second reading converts persons to fires with USFA's residential ratios of 1.44 deaths per fatal fire and 1.62 injuries per injury fire [114]: 3,920 / 1.44 = about 2,720 fatal fires and 11,780 / 1.62 = about 7,270 injury fires, together about 9,990 casualty fires or 2.9 per 100,000 for fires of all types. Fires with both a death and an injury are counted twice, which pushes the figure up; the ratios are residential and non-residential casualty fires are more often single-person, which pulls it down. The NFIRS 2024 count of fires of all types with a civilian casualty is 2.60 per 100,000 (Table S16); neither all-fires figure has a ledger row.

**Carbon monoxide incidents against NFPA.** NERIS recorded 35,776 carbon monoxide incidents with CO found in any type position (alarm activations are a separate type and are excluded). Scaled by the two incident-share factors: 35,776 x 2.047 / 3,401 = 21.5 and 35,776 x 2.418 / 3,401 = 25.4 per 100,000. NFPA's independent estimate for 2010 was 80,100 non-fire incidents in which CO was found [115]; on the 2024 population that is 80,100 / 3,401 = 23.6 per 100,000, and on the 2010 population 25.9. The 2024-population figure falls inside the scaled range and the 2010-population figure sits at its upper edge.

### S2.7. NFIRS 2024 cross-check

Source: NFIRS Public Data Release 2024 (NFIRS_2024_050125, basicincident.txt), downloaded from OpenFEMA [111]. Method: the file was read once by the counting script; aid-given records (AID 3 or 4) were dropped so each incident counts once; a casualty is OTH_DEATH or OTH_INJ greater than 0; an EMS action is any of ACT_TAK1 to ACT_TAK3 in 31 to 34; search, rescue, extrication, and recovery are actions 21 to 24. Table S12 gives the record counts and the scaling to NFPA totals.

**Table S12. NFIRS 2024 records, coverage, and scaling**

| Item | Count | Share of NFPA 2024 total |
|---|---:|---|
| Download date | 4 September 2026 | |
| Records in basicincident.txt | 33,312,139 | |
| Aid-given records dropped (AID 3 or 4) | 1,596,850 | |
| Incidents counted | 31,715,289 | 77.2% of 41,095,000 |
| Fires (incident type 100 to 173) | 964,522 | 69.5% of 1,388,000 |
| Structure fires (incident type 111 to 123) | 316,807 | 67.3% of 470,500 |
| Scaling, all incidents | 41,095,000 / 31,715,289 | x1.296 |
| Scaling, fires | 1,388,000 / 964,522 | x1.439 |
| Scaling, structure fires | 470,500 / 316,807 | x1.485 |

**Patient markers.** NFIRS fire modules populate the civilian casualty fields (OTH_DEATH, OTH_INJ) routinely, and EMS action codes on fire incidents are optional, so the casualty field is the patient marker for fires. On non-fire types the casualty fields are close to empty (4 casualties on 82,265 carbon monoxide incidents), so for rescue types the patient marker is the EMS action codes (ACT_TAK1 to ACT_TAK3 in 31 to 34), the analogue of the NERIS emergency-medical-care tactic. Both structure-fire figures are given in Table S16: 2.16 per 100,000 with a civilian casualty and 1.02 with an EMS action. NFIRS type 352 (extrication of victims from a vehicle) is coded far less often than the NERIS motor vehicle extrication or entrapment type; the closer NFIRS analogue of an extrication performed is an extrication action (23) recorded on a motor vehicle collision incident (types 322 to 324) or on a type 352 incident, and S2.5 uses both code groups.

**Table S13. Field positions in basicincident.txt used by the counting script**

The file is caret-delimited with quoted strings; the header row is skipped and quotes are stripped before the fields are read.

| Field | Position |
|---|---:|
| INC_TYPE (incident type) | $9 |
| AID (aid given or received) | $11 |
| ACT_TAK1 (action taken 1) | $19 |
| ACT_TAK2 (action taken 2) | $20 |
| ACT_TAK3 (action taken 3) | $21 |
| OTH_DEATH (civilian deaths) | $35 |
| OTH_INJ (civilian injuries) | $37 |
| PROP_USE (property use) | $41 |

**Table S14. Definitions applied by the counting script**

The short keys in the left column are the prefixes and suffixes of the count keys listed in Table S15.

| Key or term | Definition |
|---|---|
| Incident counted | AID not 3 and not 4 (aid-given records dropped so each incident counts once) |

| Key or term | Definition |
|---|---|
| Casualty (cas) | OTH_DEATH greater than 0 or OTH_INJ greater than 0 |
| Death | OTH_DEATH greater than 0 |
| Injury (inj) | OTH_INJ greater than 0 |
| EMS action (ems) | any of ACT_TAK1 to ACT_TAK3 in 31 to 34 |
| Search | any of ACT_TAK1 to ACT_TAK3 equal to 21 |
| Rescue | any of ACT_TAK1 to ACT_TAK3 equal to 22 |
| Extrication (extric) | any of ACT_TAK1 to ACT_TAK3 equal to 23 |
| Recovery (recover) | any of ACT_TAK1 to ACT_TAK3 equal to 24 |
| sr | search, rescue, extrication, or recovery (any action 21 to 24) |
| f (fires) | incident type 100 to 173 |
| sf (structure fires) | incident type 111 to 123 |
| rsf (residential structure fires) | structure fires with PROP_USE 400 to 499 |
| vf (vehicle fires) | incident type 130 to 138 |
| co (carbon monoxide incidents) | incident type 424 |
| wr (water rescue) | incident type 360 to 365; wr_any = casualty, EMS, rescue, or recovery |
| mvx (vehicle extrication) | incident type 352; mvx_any = casualty, EMS, or extrication |
| tr (technical rescue) | incident type 351, 353, 354, 355, 356, or 357; tr_any = casualty, EMS, rescue, or extrication |
| el (electrical rescue) | incident type 370 to 372 |
| op (overpressure) | incident type 240 to 243 |
| hz (hazardous condition) | incident type 400 to 431 excluding 424 |
| ems_all_300_323 | incident type 300 to 323 |
| ems_medical_311_323 | incident type 311 to 323 excluding 322 |
| mvc_322_324 | incident type 322, 323, or 324 (motor vehicle collisions) |

### Table S15. All counts in the NFIRS 2024 count file

| Key | Meaning | Count |
|---|---|---:|
| co_all | Carbon monoxide incidents (424), all incidents | 82,265 |
| co_cas | Carbon monoxide incidents (424), with a civilian casualty (OTH_DEATH or OTH_INJ greater than 0) | 4 |
| co_ems | Carbon monoxide incidents (424), with an EMS action (31 to 34) | 1,316 |
| el_all | Electrical rescue (370 to 372), all incidents | 2,639 |
| el_cas | Electrical rescue (370 to 372), with a civilian casualty (OTH_DEATH or OTH_INJ greater than 0) | 2 |
| el_ems | Electrical rescue (370 to 372), with an EMS action (31 to 34) | 251 |
| ems_all_300_323 | EMS and rescue incident types 300 to 323, all | 20,060,996 |
| ems_medical_311_323 | EMS incident types 311 to 323 excluding 322, all | 18,425,833 |
| f_all | Fires (100 to 173), all incidents | 964,522 |
| f_cas | Fires (100 to 173), with a civilian casualty (OTH_DEATH or OTH_INJ greater than 0) | 6,133 |
| f_cas_or_rescue | Fires (100 to 173), with a casualty or a rescue action | 6,849 |
| f_death | Fires (100 to 173), with a civilian death | 1,652 |
| f_ems | Fires (100 to 173), with an EMS action (31 to 34) | 5,165 |
| f_inj | Fires (100 to 173), with a civilian injury | 4,652 |
| f_rescue | Fires (100 to 173), with a rescue action (22) | 1,077 |

| Key | Meaning | Count |
|---|---|---|
| f_search | Fires (100 to 173), with a search action (21) | 10,270 |
| f_sr | Fires (100 to 173), with any search, rescue, extrication, or recovery action (21 to 24) | 11,744 |
| hz_all | Hazardous condition (400 to 431 excluding 424), all incidents | 360,283 |
| hz_cas | Hazardous condition (400 to 431 excluding 424), with a civilian casualty (OTH_DEATH or OTH_INJ greater than 0) | 6 |
| hz_ems | Hazardous condition (400 to 431 excluding 424), with an EMS action (31 to 34) | 2,186 |
| mvc_322_324_all | Motor vehicle collisions (322 to 324), all incidents | 1,742,718 |
| mvc_cas | Motor vehicle collisions (322 to 324), with a civilian casualty (OTH_DEATH or OTH_INJ greater than 0) | 14 |
| mvc_extric | Motor vehicle collisions (322 to 324), with an extrication action (23) | 26,274 |
| mvx_all | Vehicle extrication (352), all incidents | 23,936 |
| mvx_any | Vehicle extrication (352), with a casualty, EMS action, or extrication action | 17,098 |
| mvx_cas | Vehicle extrication (352), with a civilian casualty (OTH_DEATH or OTH_INJ greater than 0) | 3 |
| mvx_ems | Vehicle extrication (352), with an EMS action (31 to 34) | 6,646 |
| mvx_extric | Vehicle extrication (352), with an extrication action (23) | 14,579 |
| n | Incidents counted | 31,715,289 |
| n_aidgiven_dropped | Aid-given records dropped | 1,596,850 |
| n_all | Records in basicincident.txt | 33,312,139 |
| op_all | Overpressure (240 to 243), all incidents | 3,919 |
| op_cas | Overpressure (240 to 243), with a civilian casualty (OTH_DEATH or OTH_INJ greater than 0) | 5 |
| op_ems | Overpressure (240 to 243), with an EMS action (31 to 34) | 124 |
| rsf_all | Residential structure fires (111 to 123, property use 400 to 499), all incidents | 225,955 |
| rsf_cas | Residential structure fires (111 to 123, property use 400 to 499), with a civilian casualty (OTH_DEATH or OTH_INJ greater than 0) | 4,469 |
| rsf_death | Residential structure fires (111 to 123, property use 400 to 499), with a civilian death | 1,174 |
| rsf_ems | Residential structure fires (111 to 123, property use 400 to 499), with an EMS action (31 to 34) | 2,011 |
| rsf_inj | Residential structure fires (111 to 123, property use 400 to 499), with a civilian injury | 3,445 |
| rsf_rescue | Residential structure fires (111 to 123, property use 400 to 499), with a rescue action (22) | 908 |
| rsf_search | Residential structure fires (111 to 123, property use 400 to 499), with a search action (21) | 8,154 |
| rsf_sr | Residential structure fires (111 to 123, property use 400 to 499), with any search, rescue, extrication, or recovery action (21 to 24) | 9,161 |
| sf_all | Structure fires (111 to 123), all incidents | 316,807 |

| Key | Meaning | Count |
|---|---|---|
| sf_cas | Structure fires (111 to 123), with a civilian casualty (OTH_DEATH or OTH_INJ greater than 0) | 4,942 |
| sf_cas_or_rescue | Structure fires (111 to 123), with a casualty or a rescue action | 5,560 |
| sf_death | Structure fires (111 to 123), with a civilian death | 1,258 |
| sf_ems | Structure fires (111 to 123), with an EMS action (31 to 34) | 2,343 |
| sf_inj | Structure fires (111 to 123), with a civilian injury | 3,842 |
| sf_rescue | Structure fires (111 to 123), with a rescue action (22) | 960 |
| sf_search | Structure fires (111 to 123), with a search action (21) | 9,255 |
| sf_sr | Structure fires (111 to 123), with any search, rescue, extrication, or recovery action (21 to 24) | 10,326 |
| tr_all | Technical rescue (351, 353 to 357), all incidents | 110,796 |
| tr_any | Technical rescue (351, 353 to 357), with a casualty, EMS, rescue, or extrication action | 65,058 |
| tr_cas | Technical rescue (351, 353 to 357), with a civilian casualty (OTH_DEATH or OTH_INJ greater than 0) | 3 |
| tr_ems | Technical rescue (351, 353 to 357), with an EMS action (31 to 34) | 2,944 |
| tr_extric | Technical rescue (351, 353 to 357), with an extrication action (23) | 12,257 |
| tr_rescue | Technical rescue (351, 353 to 357), with a rescue action (22) | 51,263 |
| tr_type_351 | Technical rescue, incident type 351 | 3,445 |
| tr_type_353 | Technical rescue, incident type 353 | 101,569 |
| tr_type_354 | Technical rescue, incident type 354 | 647 |
| tr_type_355 | Technical rescue, incident type 355 | 696 |
| tr_type_356 | Technical rescue, incident type 356 | 2,244 |
| tr_type_357 | Technical rescue, incident type 357 | 2,195 |
| vf_all | Vehicle fires (130 to 138), all incidents | 139,328 |
| vf_cas | Vehicle fires (130 to 138), with a civilian casualty (OTH_DEATH or OTH_INJ greater than 0) | 738 |
| vf_death | Vehicle fires (130 to 138), with a civilian death | 326 |
| vf_ems | Vehicle fires (130 to 138), with an EMS action (31 to 34) | 1,778 |
| vf_extric | Vehicle fires (130 to 138), with an extrication action (23) | 188 |
| vf_inj | Vehicle fires (130 to 138), with a civilian injury | 424 |
| vf_rescue | Vehicle fires (130 to 138), with a rescue action (22) | 66 |
| wr_all | Water rescue (360 to 365), all incidents | 20,335 |
| wr_any | Water rescue (360 to 365), with a casualty, EMS, rescue, or recovery action | 9,382 |
| wr_cas | Water rescue (360 to 365), with a civilian casualty (OTH_DEATH or OTH_INJ greater than 0) | 0 |
| wr_ems | Water rescue (360 to 365), with an EMS action (31 to 34) | 1,908 |

| Key | Meaning | Count |
|---|---|---|
| wr_recover | Water rescue (360 to 365), with a recovery action (24) | 387 |
| wr_rescue | Water rescue (360 to 365), with a rescue action (22) | 7,616 |

#### Table S16. NFIRS 2024 cross-check rows, scaled to NFPA totals

| Row | NFIRS 2024 | Scaling | Scaled | Per 100,000 | Ledger comparison |
|---|---|---|---|---|---|
| Fires with a civilian casualty | 6,133 | fires x1.439 | 8,826 | 2.60 | structure fire with a patient treated 2.2; NFPA casualty-based 1.4 |
| Structure fires with a civilian casualty | 4,942 | structure fires x1.485 | 7,340 | 2.16 | 2.2 (2.0 to 2.4) |
| Structure fires with an EMS action (31 to 34) | 2,343 | structure fires x1.485 | 3,480 | 1.02 | casualty field is the better NFIRS marker for fires |
| Structure fires with a search action (21) | 9,255 | structure fires x1.485 | 13,745 | 4.04 | search-only context 10.2 to 12.1 (different tactic vocabularies) |
| Vehicle fires with an EMS action | 1,778 | fires x1.439 | 2,559 | 0.75 | 0.7 (0.7 to 0.8) |
| Carbon monoxide incidents (424) with an EMS action | 1,316 | all incidents x1.296 | 1,705 | 0.50 | CO with a patient 0.5 to 0.6 (context; counted on the medical side) |
| Water rescue (360 to 365) with an EMS action | 1,908 | all incidents x1.296 | 2,472 | 0.73 | 0.7 (0.6 to 0.7) |
| Vehicle extrication (352) with an EMS action | 6,646 | all incidents x1.296 | 8,612 | 2.53 | 2.3 (1.6 to 9.0) |
| Extrication action (23) on motor vehicle collisions (322 to 324) or vehicle extrications (352) | 40,853 | all incidents x1.296 | 52,935 | 15.56 | NFIRS reading of extrications performed for the entrapment row |
| Motor vehicle collisions (322 to 324) with an extrication action (23) | 26,274 | all incidents x1.296 | 34,044 | 10.01 | collision-typed incidents only |
| Vehicle extrication (352), all | 23,936 | all incidents x1.296 | 31,015 | 9.12 | typed extrication or entrapment 27 to 32 (context) |
| Water rescue (360 to 365), all | 20,335 | all incidents x1.296 | 26,349 | 7.75 | person in water by incident type 7 to 8 (context) |
| Technical rescue (351, 353 to 357) with an EMS action | 2,944 | all incidents x1.296 | 3,815 | 1.12 | other extrication or entrapment with a patient treated 1.4 |

On the marker each system populates, the NFIRS figure falls within or beside the ledger ranges for structure fires (civilian casualty, 2.16 per 100,000), vehicle fires (EMS action, 0.75), water rescues (EMS action, 0.73), and the carbon monoxide context row (EMS action, 0.50). Fires of all types with a civilian casualty (2.60) have no ledger row; the NFPA-derived 2.9 in S2.6 is the same all-fires quantity. For vehicle extrication, NFIRS type 352 with an EMS action gives 2.53, a treated-patient count under NFIRS's narrower typing; extrication actions on collision or extrication incidents give 15.6, the floor for extrications performed used in S2.5 (collision-typed incidents alone give 10.0); the ledger's motor vehicle crash row is 2.3 (1.6 to 9.0).

### S3. Counting bases and triangulation

#### S3.1. Fire-department responses and incidents

NFPA's Fire Experience Survey counts fire-department responses, and a mutual-aid response to another department's incident is counted again by the aiding department. The 2024 survey reports 42,687,000 responses, of which 1,592,000 were mutual aid [108]. The net figure, 42,687,000 less 1,592,000 = 41,095,000 incidents, or 41,095,000 / 3,401 = 12,083 per 100,000, is the fire-department layer of the funnel and the numerator of the NERIS all-incident scaling factor.

#### S3.2. EMS activations and patient events

A NEMSIS activation is one EMS unit's response record. A patient event generates more than one activation when several units respond or when a patient is handed between units. In the opioid overdose row [45], the authors kept the final responding agency's record and acknowledged residual duplication; this does not establish a correction for the other activation-based rows. In New York City assault data, Morrison and colleagues reconciled 32,202 responses to 26,451 presumed patient encounters (82.1% retained) [116]. This single-city, single-call-type result supports testing response-to-encounter retention; it is not a national correction or an admission-duplication estimate. The reference counts activation-based rows at face value (retention 1.0), while the lower-count scenario and one sensitivity line discount them (S3.6). The NEMSIS acuity layers count activations.

#### S3.3. NEMSIS acuity elements and codes

NEMSIS records acuity twice on each activation: Initial Patient Acuity (eSituation.13) and Final Patient Acuity (eDisposition.19) [5]. Critical is coded 2813001 initially and 4219001 finally; Emergent is coded 2813003 initially and 4219003 finally. Responses to 911 calls were selected with eResponse.05 (Type of Service Requested) = 2205001. All counts are from the 2024 public-release research dataset, and all rates are per 100,000 residents. The original code-frequency extraction and its source URL are preserved in nemsis-acuity-inputs.json. Code 7701001 (Not Applicable) is excluded from the adjustment pool. Only code 7701003 (Not Recorded) is imputed. Both elements permit these two NOT values; 7701005 (Not Reporting) is not permitted. All recorded categories remain in the observed denominator, including dead and routine acuity. Initial code 2813009 means Non-Acute/Routine in the NEMSIS v3.5 dictionary; its 2,216,783 records are observed noncritical acuity.

**Table S17. NEMSIS 2024 acuity-code eligibility and missingness by extraction scope**

| Scope | All activations N | Not Applicable A (7701001) | Not Recorded R (7701003) | Eligible E = N - A | Observed O = E - R | R / E |
|---|---|---|---|---|---|---|
| Initial acuity, all service types | 60,298,684 | 4,220,975 | 14,873,499 | 56,077,709 | 41,204,210 | 26.5% |
| Final acuity, all service types | 60,298,684 | 5,550,043 | 19,586,250 | 54,748,641 | 35,162,391 | 35.8% |
| Final acuity, 911 responses only | 46,733,668 | 4,955,766 | 16,368,893 | 41,777,902 | 25,409,009 | 39.2% |

**Missing-at-random arithmetic.** For each recorded count C, the observed rate is C / 3,401 and the adjusted rate is (C / 3,401) x E / O, equivalently (C / 3,401) / (1 - R / E). Not Applicable records receive no imputed cases. The scenario assumes equal acuity distributions among Not Recorded and observed records within each eligible pool. Acuity coding defines eligibility here; the extraction cannot confirm patient contact. The scenario is neither an upper bound nor a patient-level estimate.

**Table S18. Recorded rates and missing-at-random scenarios excluding Not Applicable**

| Measure | Recorded count C | Recorded per 100,000 | Multiplier E / O | MAR scenario per 100,000 |
|---|---|---|---|---|
| Initial acuity Critical, all service types | 1,564,207 | 459.9 | 56,077,709 / 41,204,210 | 626 |
| Final acuity Critical, all service types | 990,158 | 291.1 | 54,748,641 / 35,162,391 | 453 |
| Final acuity Critical, 911 responses only | 816,411 | 240.1 | 41,777,902 / 25,409,009 | 395 |
| Final acuity Emergent, all service types | 6,818,691 | 2,004.9 | 54,748,641 / 35,162,391 | 3,122 |
| Final acuity Emergent, 911 responses only | 5,796,511 | 1,704.4 | 41,777,902 / 25,409,009 | 2,802 |
| Final acuity Emergent or Critical, 911 responses only | 6,612,922 | 1,944.4 | 41,777,902 / 25,409,009 | 3,197 |

Rates use the unrounded recorded count and exact integer multiplier; displayed rates are rounded only at the end. **Reading the acuity figures.** Acuity counts activations, and activations outnumber patient events (S3.2), which pushes the acuity figures up. Not Recorded accounts for 26.5% to 39.2% of acuity-applicable records, and a patient resuscitated or stabilized in the field is often downgraded by the end of the call, both of which pull the recorded Critical counts down. Recorded Critical acuity (240 to 460 per 100,000 residents) and the adjusted values (395 to 626) frame the medical estimate of 583 net after overlap; the Emergent-or-Critical 911 layer (1,944 as recorded, 3,197 adjusted) sits above it. The frame is not independent of the estimate: several medical rows are themselves NEMSIS-derived, and the adjustment assumes that applicable records without acuity carry the same Critical share as observed records.

#### S3.4. Rate derived from the Jarvis inputs

Jarvis and colleagues report a service population of about 650,000, about 40,000 incidents a year, and a time-critical intervention on 6.9% to 7.6% of incidents [117]. The per-population rate is our arithmetic on those inputs: 40,000 x 0.069 / 650,000 x 100,000 = 425, and 40,000 x 0.076 / 650,000 x 100,000 = 468 per 100,000 a year. The definition (an intervention performed) and the denominator (one agency's incidents, which excludes cancelled calls) differ from the ledger's, and the intervention list includes stroke and STEMI alerts, which are notifications with no treatment performed; the two figures are compared for order of magnitude only.

#### S3.5. Funnel and shares

**Table S19. Funnel per 100,000 residents, bases labeled**

| Component | Basis | Per 100,000 |
|---|---|---:|
| Fire-department incidents net of mutual aid (NFPA 2024: 42,687,000 responses less 1,592,000 mutual aid) | incidents | 12,083 |
| EMS activations, all service types (NEMSIS 2024) | activations | 17,730 |
| EMS activations, 911 responses only | activations | 13,741 |
| 911 activations Emergent or Critical at final acuity | activations, as recorded | 1,944 |
| Structure fires (NFPA 2024) | incidents | 138 |
| Attended time-critical events, net after overlap | patient events | 589 |
| Tier A medical, net | patient events | 350 |

Shares, computed from the unrounded ledger values: attended events (589) are 4.3% of 911 activations (589.5 / 13,741.2), and the medical rows alone (583) are 4.2% (583.2 / 13,741.2). Attended events are 3.3% of all activations (589.5 / 17,729.7) and 30% of 911 activations coded Emergent or Critical at final acuity as recorded (589.5 / 1,944.4). Tier A alone (350) is 2.5% of 911 activations. The acuity layers count activations and the ledger counts events, so the event-basis shares are higher than shown.

#### S3.6. The ordered union

Rows are not mutually exclusive: a poisoning that arrests appears in the opioid source and in CARES, a trapped crash occupant in shock appears in the blunt trauma source and in the entrapment count. The ledger combines the rows by an ordered union. Rows enter in a fixed order, cardiac arrest first, then the other medical rows, then the fire and rescue block. For each row the qualifying events are the rate times a retention factor for the counting unit times an eligibility share; the deduction is an assumed fraction of the qualifying events taken to lie inside the union of the rows already counted, capped at that union; the net is what the row adds. The opioid row's deduction is a share of the arrest row instead: 5.3% of treated arrests carry an overdose etiology in CARES, and the reference takes half of that pool as already inside the opioid source (all of it in the lower-count scenario, none in the higher-count). Table S20 gives every step at the reference coefficients.

Retention is 1.0 for every counting unit in the reference: no transferable national duplicate-record rate was established. Taylor reconciled agencies for the opioid source [45]; Morrison's New York City assault study supports testing activation retention but does not supply a general correction [116]. The lower-count scenario takes 75% of activation records and 80% of admissions as distinct events; two sensitivity lines use 85% and 90%. Eligibility is 1.0 for most rows; the septic shock row applies the 87.9% community-onset share from Rhee [118] because the CDC sepsis count includes hospital-onset cases a crew cannot attend, with the caveat that community onset means criteria met before hospital day 3, a wider net than shock before EMS arrival. The other lower-count eligibility values (opioid 70%, status epilepticus 50%, carbon monoxide 50%, hypoglycemia 75%, nonfatal drowning 50% in the lower-count scenario) test narrower case definitions. Every fraction is a stated assumption; the lower-count and higher-count sets are coherent assumption sets with no probability attached.

**Table S20. Ordered union at the reference coefficients (per 100,000)**

| Order | Row | Rate | Retention | Eligibility | Qualifying | Overlap fraction | Deduction | Net |
|---|---|---|---|---|---|---|---|---|
| 1 | Cardiac arrest, EMS-treated, non-traumatic | 78.70 | 1.00 | 1.000 | 78.70 | 0.00 | 0.00 | 78.70 |
| 2 | Ischemic stroke, EMS-transported | 120.19 | 1.00 | 1.000 | 120.19 | 0.00 | 0.00 | 120.19 |
| 3 | Adult nonfatal opioid-overdose EMS encounters, selected dispositions | 125.47 | 1.00 | 1.000 | 125.47 | share of the arrest pool | 2.09 | 123.39 |
| 4 | STEMI, EMS-transported | 30.00 | 1.00 | 1.000 | 30.00 | 0.05 | 1.50 | 28.50 |
| 5 | Intracerebral hemorrhage, EMS-transported | 19.80 | 1.00 | 1.000 | 19.80 | 0.02 | 0.40 | 19.40 |
| 6 | Subarachnoid hemorrhage, EMS-transported | 6.11 | 1.00 | 1.000 | 6.11 | 0.02 | 0.12 | 5.99 |
| 7 | Septic shock, EMS-attended | 30.00 | 1.00 | 0.879 | 26.37 | 0.02 | 0.53 | 25.84 |
| 8 | Adult asthma/COPD encounters with assisted ventilation, EMS treated and transported | 36.10 | 1.00 | 1.000 | 36.10 | 0.03 | 1.08 | 35.02 |
| 9 | Presumed status epilepticus: EMS benzodiazepine-treated seizures | 29.62 | 1.00 | 1.000 | 29.62 | 0.02 | 0.59 | 29.03 |
| 10 | Penetrating trauma, exsanguinating subset, EMS | 5.32 | 1.00 | 1.000 | 5.32 | 0.00 | 0.00 | 5.32 |
| 11 | Blunt trauma with hemorrhagic shock, EMS | 19.65 | 1.00 | 1.000 | 19.65 | 0.01 | 0.20 | 19.46 |
| 12 | Severe traumatic brain injury (GCS 8 or less) without hypotension, EMS-transported | 4.20 | 1.00 | 1.000 | 4.20 | 0.00 | 0.00 | 4.20 |
| 13 | Acute heart failure with respiratory failure (CPAP or ventilation), EMS-attended | 16.60 | 1.00 | 1.000 | 16.60 | 0.10 | 1.66 | 14.94 |
| 14 | Anaphylaxis, EMS-attended | 5.00 | 1.00 | 1.000 | 5.00 | 0.02 | 0.10 | 4.90 |
| 15 | Acute aortic dissection, EMS-transported | 2.00 | 1.00 | 1.000 | 2.00 | 0.05 | 0.10 | 1.90 |

| Order | Row | Rate | Retention | Eligibility | Qualifying | Overlap fraction | Deduction | Net |
|---|---|---|---|---|---|---|---|---|
| 16 | Unstable arrhythmia needing pacing or cardioversion, EMS | 4.86 | 1.00 | 1.000 | 4.86 | 0.10 | 0.49 | 4.37 |
| 17 | Ruptured abdominal or thoracic aortic aneurysm, EMS-transported | 1.00 | 1.00 | 1.000 | 1.00 | 0.05 | 0.05 | 0.95 |
| 18 | High-risk pulmonary embolism, EMS-transported | 1.40 | 1.00 | 1.000 | 1.40 | 0.20 | 0.28 | 1.12 |
| 19 | Carbon monoxide poisoning, unintentional non-fire, EMS or fire department attended | 2.25 | 1.00 | 1.000 | 2.25 | 0.02 | 0.05 | 2.21 |
| 20 | Non-opioid poisoning with respiratory or cardiovascular compromise, EMS | 7.00 | 1.00 | 1.000 | 7.00 | 0.05 | 0.35 | 6.65 |
| 21 | Heat stroke, EMS-attended | 2.60 | 1.00 | 1.000 | 2.60 | 0.01 | 0.03 | 2.57 |
| 22 | Hypoglycemia treated with dextrose or glucagon | 25.41 | 1.00 | 1.000 | 25.41 | 0.00 | 0.00 | 25.41 |
| 23 | Acute alcohol intoxication with airway compromise requiring intubation, EMS-attended | 16.90 | 1.00 | 1.000 | 16.90 | 0.15 | 2.53 | 14.36 |
| 24 | Pediatric airway-procedure workload proxy | 2.80 | 1.00 | 1.000 | 2.80 | 0.10 | 0.28 | 2.52 |
| 25 | Obstetric and neonatal emergencies attended in the field (delivery complications, high-acuity intrapartum or postpartum hemorrhage, eclamptic seizure, neonatal resuscitation) | 1.40 | 1.00 | 1.000 | 1.40 | 0.20 | 0.28 | 1.12 |

| Order | Row | Rate | Retention | Eligibility | Qualifying | Overlap fraction | Deduction | Net |
|---|---|---|---|---|---|---|---|---|
| 26 | Gastrointestinal bleeding presenting in hemorrhagic shock, EMS-attended | 2.00 | 1.00 | 1.000 | 2.00 | 0.02 | 0.04 | 1.96 |
| 27 | Ruptured ectopic pregnancy with hemodynamic instability, EMS-attended | 0.30 | 1.00 | 1.000 | 0.30 | 0.02 | 0.01 | 0.29 |
| 28 | Upper airway obstruction other than foreign body (angioedema, epiglottitis, deep neck infection) requiring airway intervention | 0.30 | 1.00 | 1.000 | 0.30 | 0.10 | 0.03 | 0.27 |
| 29 | Major burn with inhalation injury or airway involvement, EMS-transported | 0.55 | 1.00 | 1.000 | 0.55 | 0.02 | 0.01 | 0.54 |
| 30 | Drowning, nonfatal, EMS-attended | 2.38 | 1.00 | 1.000 | 2.38 | 0.15 | 0.36 | 2.03 |
| 31 | Structure fire with a patient treated | 2.20 | 1.00 | 1.000 | 2.20 | 0.10 | 0.22 | 1.98 |
| 32 | Vehicle fire with a patient treated | 0.72 | 1.00 | 1.000 | 0.72 | 0.10 | 0.07 | 0.65 |
| 33 | Motor vehicle crash with a physically trapped patient | 2.27 | 1.00 | 1.000 | 2.27 | 0.50 | 1.14 | 1.14 |
| 34 | Other extrication or entrapment (structure, machinery, outside) with a patient treated | 1.44 | 1.00 | 1.000 | 1.44 | 0.25 | 0.36 | 1.08 |
| 35 | Person in water (standing or swift) with a patient treated | 0.67 | 1.00 | 1.000 | 0.67 | 0.50 | 0.34 | 0.34 |
| 36 | Watercraft in distress with a patient treated | 0.10 | 1.00 | 1.000 | 0.10 | 0.25 | 0.03 | 0.08 |
| 37 | Structural collapse with a patient treated | 0.13 | 1.00 | 1.000 | 0.13 | 0.25 | 0.03 | 0.10 |
| 38 | Confined space or trench rescue with a patient treated | 0.09 | 1.00 | 1.000 | 0.09 | 0.25 | 0.02 | 0.06 |

| Order | Row | Rate | Retention | Eligibility | Qualifying | Overlap fraction | Deduction | Net |
|---|---|---|---|---|---|---|---|---|
| 39 | Rope rescue (high, steep, low angle) with a patient treated | 0.33 | 1.00 | 1.000 | 0.33 | 0.25 | 0.08 | 0.25 |
| 40 | Backcountry rescue with a patient treated | 0.49 | 1.00 | 1.000 | 0.49 | 0.25 | 0.12 | 0.37 |
| 41 | Explosion or overpressure rupture with a patient treated | 0.09 | 1.00 | 1.000 | 0.09 | 0.25 | 0.02 | 0.06 |
| 42 | Hazardous materials release with a patient treated | 0.14 | 1.00 | 1.000 | 0.14 | 0.25 | 0.04 | 0.11 |
| 43 | Aviation crash with a patient treated | 0.18 | 1.00 | 1.000 | 0.18 | 0.25 | 0.04 | 0.13 |
| | Medical rows | | | | 596.3 | | 13.1 | 583.2 |
| | Fire and rescue rows | | | | 8.9 | | 2.5 | 6.3 |
| | Attended time-critical events | | | | 605.1 | | 15.7 | 589.5 |

The rows sum to 608.8 per 100,000; eligibility brings the qualifying events to 605.1; the deductions total 15.7, 2.6% of the qualifying sum. The joint lower-count scenario gives 474 and the joint higher-count scenario 609; at the rows' low and high values with the reference coefficients the total is 494 to 697 (Table S22).

### S3.7. Retained pathways

Five pathways meet the time-critical definition and have no US incidence anchor. They are kept in view with scenario values built from the sources in the table, and they enter the total in one sensitivity line only. Table S21 gives each with the reason it stays outside the sum. The severe foreign-body airway obstruction value is the Alberta intervention-defined rate; the paper's transfer rule admits a borrowed clinical share with a stated reason and excludes a whole foreign incidence rate from the sum, so the Alberta rate is shown as a scenario and not transferred.

**Table S21. Retained pathways: no US incidence anchor; scenario values shown outside the sum**

| Pathway | Tier | Scenario value | Range | Anchor | Arithmetic | Why outside the sum | Evidence | Sources |
|---|---|---|---|---|---|---|---|---|
| Severe foreign-body airway obstruction requiring intervention, EMS-attended | B | 4.0 | 1.6 to 12.4 | Alberta 2018 to 2021 (population about 4.4 million): 3,677 EMS encounters for foreign-body airway obstruction (20.9 per 100,000 a year), 709 requiring an intervention (4.0 per 100,000 a year); 42.4% died when the obstruction was unrelieved at paramedic arrival, 0.4% when relieved before arrival (Dunne 2024). | 4.0 per 100,000 as published. Cross-check: 5,553 US choking deaths (NSC 2022) / 0.424 = 13,100 = 3.9 per 100,000. | The only intervention-defined incidence is Canadian (Alberta); a whole foreign incidence rate is not transferred into the sum. | No measured interval; treatment before and after EMS arrival compared: neurologic outcome by the stage at which the foreign body was removed (Igarashi 2017); case fatality 42.4% when unrelieved at paramedic arrival against 0.4% when relieved before (Dunne 2024). | [119,120,121,122] |
| Severe accidental hypothermia, EMS-attended | B | 0.9 | 0.3 to 1.8 | Age-adjusted cold-related death rate 0.92 per 100,000 in 2022 (Liu 2025); 1,024 deaths attributed to excessive cold or hypothermia in 2023 (CDC QuickStats 2025). | 0.92 carried as 0.9; low 0.3 from the strict 1,024 deaths (1,024 / 3,401); high 1.8 allows nonfatal severe cases at twice the death rate (2 x 0.92). | A US death rate stands in for attended severe cases; no US EMS or hospitalization count exists. | Consensus: rewarming and arrest prevention in the Wilderness Medical Society guideline (Dow 2019); no time-to-outcome study located. | [123,124,125] |
| Electrical injury (non-arrest), EMS-attended | B | 0.5 | 0.2 to 1.5 | About 1,000 electrocution deaths and at least 30,000 nonfatal shock injuries a year (textbook figures as summarized in StatPearls and Karray 2025). | 30,000 x 0.10 x 0.60 = 1,800; / 3,401 = 0.5. Bounds 0.2 to 1.5 span serious shares of 4% to 28%. | Textbook injury figures with two assumed shares; no US dataset count of serious non-arrest cases. | Clinical reasoning: arrhythmia and airway compromise after high-voltage injury; no guideline or time-to-outcome study cited. | [126,127] |
| Hemodialysis vascular-access hemorrhage, EMS-attended | B | 0.2 | 0.1 to 0.7 | Fatal vascular-access hemorrhage fewer than 1 per 1,000 hemodialysis patient-years (Jose 2017); 78% of fatal cases at home (Ellingson 2012); about 560,000 US hemodialysis patients. | 560,000 x 0.001 = 560 fatal events; / 3,401 = 0.16, carried as 0.2. Low 0.1 uses half that rate; high 0.7 allows three nonfatal attended events per fatal one. | A fatal-event rate gives a floor; the attended nonfatal count is unknown. | Clinical reasoning: direct pressure and hemorrhage control; fatal within minutes untreated (Ellingson 2012); no guideline or time-to-outcome study cited. | [128,129] |

| Pathway | Tier | Scenario value | Range | Anchor | Arithmetic | Why outside the sum | Evidence | Sources |
|---|---|---|---|---|---|---|---|---|
| Device-dependent patient emergencies (tracheostomy obstruction or dislodgement, home ventilator failure, LVAD failure), EMS-attended | B | 0.3 | 0.1 to 0.5 | 28,029 continuous-flow LVADs implanted (Intermacs 2025); device alarms 15% of 75 EMS transports of LVAD patients (Larson 2026); 44% of pediatric tracheostomy patients had an emergency department visit within 90 days, 36% airway-related (Tarfa 2021). US home-ventilator and home-tracheostomy populations are unregistered. | Bounded at 0.3 (0.1 to 0.5) per 100,000 from device populations and cohort event rates. | No national device population or emergency count. | Clinical reasoning: tracheostomy obstruction, ventilator failure, and LVAD pump failure are fatal within minutes untreated and reliably managed by a crew (Goebel 2019; Larson 2026); no guideline or time-to-outcome study cited. | [130,131,132,133] |
| Sum of scenario values | | 5.9 | 2.3 to 16.9 | | | Added in one sensitivity line: 595 in place of 589 | | |

### S3.8. Sensitivity

Table S22 reproduces the main text's sensitivity table with the medical and fire components of each variant. The first two variants after the reference are the summed bounds: every row at its low or high value with the reference coefficients. The next two are the joint coefficient scenarios. Each remaining line changes one thing.

**Table S22. Sensitivity of the attended count (net after overlap, per 100,000)**

| Variant | Kind | Medical | Fire and rescue | Attended events | Change |
|---|---|---:|---:|---:|---:|
| Reference: every row at its point, retention 1.0, reference eligibility and overlap | reference | 583.2 | 6.3 | 589 | +0 |
| Every row at its low value (summed bounds) | bounds | 488.5 | 5.6 | 494 | -95 |
| Every row at its high value (summed bounds) | bounds | 686.9 | 10.1 | 697 | +108 |
| Joint lower-count coefficients: activation records 75% and admissions 80% distinct events, stricter eligibility, higher overlap | joint | 469.9 | 4.6 | 474 | -115 |
| Joint higher-count coefficients: retention 1.0, every row fully eligible, every overlap fraction zero | joint | 599.9 | 8.9 | 609 | +19 |

| Variant | Kind | Medical | Fire and rescue | Attended events | Change |
|---|---|---|---|---|---|
| Activation records at 85% distinct events | single | 564.4 | 6.3 | 571 | -19 |
| Admissions at 90% distinct events | single | 578.4 | 6.3 | 585 | -5 |
| Stricter eligibility on six rows (opioid, septic shock, status epilepticus, carbon monoxide, hypoglycemia, nonfatal drowning) | single | 520.2 | 6.3 | 527 | -63 |
| Overlap fractions at their high values | single | 565.7 | 4.6 | 570 | -19 |
| Every overlap fraction zero | single | 596.3 | 8.9 | 605 | +16 |
| Eight NEMSIS or ESO sample rows at their source-year counts, no scaling to 2024 activation volume (a floor) | single | 508.6 | 6.3 | 515 | -75 |
| Severe sepsis (97.5, the midpoint of 75 to 120) in place of septic shock (30) | single | 641.3 | 6.3 | 648 | +58 |
| EMS-assessed cardiac arrest (110.8, the 2015 ROC extrapolation) in place of EMS-treated | single | 614.4 | 6.3 | 621 | +31 |
| Retained pathways added at their scenario values | single | 583.2 | 6.3 | 595 | +6 |
| Vehicle entrapment at every typed extrication with a patient treated | single | 583.2 | 9.4 | 593 | +3 |
| Structure fires with a search performed and no patient treated added | single | 583.2 | 17.5 | 601 | +11 |
| Nonfatal drowning row removed | single | 581.1 | 6.3 | 587 | -2 |

### S4. Coverage and representativeness of NEMSIS and NERIS

#### S4.1. NEMSIS

The 2024 National EMS Data Report describes the dataset as a "near complete census" covering "99% of U.S. counties in 2024"; the 2023 report says records come from "nearly 95 percent of all EMS agencies nationwide"; and the public-release research dataset user manual states "THE PUBLIC RELEASE RESEARCH DATASET IS NOT A POPULATION BASED DATA SET." [5] The statements are consistent: agency coverage is near complete, and the public-release file is an activation file with no population denominator, which is why rates here use the Census population. Following the agency-coverage statement, NEMSIS-derived event counts are treated as slight undercounts. NEMSIS acuity is read as described in S3.3, with 26.5% to 39.2% of acuity-applicable records coded Not Recorded (Table S17); Not Applicable records are excluded from the adjustment pool.

#### S4.2. NERIS

NERIS participation is voluntary and incomplete. In the twelve-month window 17,103 departments reported at least one incident [107]. By recorded department type they were 2,242 career, 3,623 combination, 9,307 volunteer, and 1,922 with no type recorded;

those four groups sum to 17,094, which leaves 9 departments with another or unparsed type value. No national comparator is given for the mix because the NFPA department profile counts departments on a different basis. The public data carry no civilian casualty, injury, death, or patient fields, so a recorded emergency-medical-care tactic is the marker of a patient treated; that tactic appears on 70.5% of incidents typed MEDICAL, so the fire and rescue counts may understate patients treated, and no adjustment is applied because the recording rate at a fire is not known (S2.2). Only 6,168 departments carry a population_protected value and their jurisdictions overlap, so scaling is by incident share against NFPA national totals (S2.3). Participation skews toward departments that adopted the system early, and the scaling factors carry that department mix with them: the structure-fire and all-incident factors differ, and the range of every fire and rescue row spans both.

#### S4.3. NFIRS

NFIRS is not a census of US fire departments. After dropping aid-given records, the 2024 public release holds 31,715,289 incidents, 77.2% of NFPA's 41,095,000; 964,522 fires, 69.5% of 1,388,000; and 316,807 structure fires, 67.3% of 470,500 [108,109,111]. Those shares are the inverses of the NFIRS scaling factors in Table S12.

### S5. Considered and excluded

The authors reviewed candidates by mechanism (airway, circulation, brain, environment, trauma), by population (children, older adults, pregnant women, patients dependent on medical devices), by responder (EMS, fire suppression, technical rescue, bystanders), and by calls routine enough to escape a disease-based list. Table S23 lists the medical and operational candidates excluded with the reason for each, and Table S24 lists the NERIS incident types considered for the fire and rescue rows and excluded.

**Table S23. Candidates considered and excluded**

| Candidate | Reason for exclusion | Where it appears instead |
|---|---|---|
| Behavioral emergencies (suicide attempts, psychiatric crises) | Excluded as a category; the time-critical cases are already counted by mechanism | Poisoning (opioid and non-opioid rows), asphyxia (airway obstruction and cardiac arrest rows), trauma (penetrating and blunt rows) |
| Interfacility critical-care transfers | Hospital-to-hospital, not community events | Nowhere |
| Severe sepsis without shock | Hours-scale | Broader definition of the septic shock row (S1); sensitivity variant gives 648 in place of 589 |
| Non-ST-elevation myocardial infarction | Hours-scale | Broader definition of the STEMI row (all acute myocardial infarction), not carried |
| Gastrointestinal bleeding without shock | Hours-scale | Broader definition of the gastrointestinal bleeding row, not carried |
| Diabetic ketoacidosis | Hours-scale | Nowhere |
| Hip fracture | Hours-scale | Nowhere |
| Spinal cord injury stabilization | Hours-scale | Nowhere; injured patients in shock or coma are in the blunt trauma row |
| Envenomation | Hours-scale | Anaphylaxis to venom falls within the all-cause anaphylaxis row |
| Button-battery ingestion | Hours-scale | Nowhere |
| Sudden infant death | Response cannot change the outcome | Nowhere; infants on whom resuscitation was attempted are in the EMS-treated cardiac arrest row |
| Patients dead at discovery | No responder reached a live patient; outside the attended count by construction | Not estimated in this paper; the cardiac arrest row counts worked arrests only |
| Long lie after a fall | Discovery time, not response time | Not estimated; no registry measures how often a responder reaches the person alive |
| Severe foreign-body airway obstruction, severe hypothermia, electrical injury, dialysis-access hemorrhage, device-dependent emergencies | Time-critical, with no US incidence anchor | Retained pathways (Table S21); one sensitivity line adds their scenario values |
| Lift assists | No time-critical patient | Nowhere; 685,879 NERIS incidents typed lift assist in the window (Table S5) |
| Structure fires with a search but no patient treated | A search does not establish that a person was inside | Context row (Table S10); sensitivity variant adds them and gives 601 |
| Residential structure fires as a class | Most are found with no one in danger | Nowhere as a class; 116,943 of the 229,796 first-position structure fires had a residential location use (Table S24), and residential fires enter only through the structure fire row when a patient was treated |

| Candidate | Reason for exclusion | Where it appears instead |
|---|---|---|
| Carbon monoxide alarm activations | No patient; alarm without CO found | Nowhere; 84,840 NERIS incidents typed CO alarm in the window are excluded from the CO context row (Table S10) |
| Wildfire civilian entrapment | Meets the test; no dataset counts it | Named as a gap; not counted (Table S24) |
| Mass-casualty and active-shooter events | Their patients are counted individually by injury | Penetrating trauma, blunt trauma, and burn rows |
| Anaphylaxis presenting to an emergency department without EMS | Not EMS-attended | Broader definition of the anaphylaxis row (S1), not carried |

**Table S24. NERIS incident types considered for the fire and rescue rows and excluded**

| NERIS type | Incidents in the window | Reason for exclusion | Where it appears instead |
|---|---:|---|---|
| RESCUE / STRUCTURE / ELEVATOR_ESCALATOR_RESCUE | 78,318 | No time-critical patient in the ordinary case | Nowhere |
| PUBSERV / CITIZEN_ASSIST / LOST_PERSON | 6,499 | Discovery-time problem; patient status unknown | Nowhere |
| PUBSERV / CITIZEN_ASSIST / PERSON_IN_DISTRESS | 45,153 | Discovery-time problem; patient status unknown | Nowhere |
| RESCUE / TRANSPORTATION / TRAIN_RAIL_COLLISION_DERAILMENT | 490 | No patient marker separable from the type | Inside the trauma rows when a person is injured |
| FIRE / OUTSIDE_FIRE / WILDFIRE_WILDLAND | 21,945 | Civilian entrapment meets the test; no field records it | Named as a gap |
| FIRE / OUTSIDE_FIRE / WILDFIRE_URBAN_INTERFACE | 5,176 | Civilian entrapment meets the test; no field records it | Named as a gap |
| MEDICAL / INJURY / MOTOR_VEHICLE_COLLISION | 728,136 | Injured occupants without entrapment are EMS patients | Trauma rows, where time-critical; the motor vehicle crash row counts physical entrapment |
| FIRE / STRUCTURE_FIRE with a residential location use (first type position) | 116,943 | Most are found with no one in danger | Structure fire row, only when a patient was treated |

### Natural-history notes

Several Tier B rows enter on clinical reasoning about the untreated course: the condition is lethal untreated and a responding crew reverses it, so the row qualifies whatever its outcomes look like once a service exists. Table S25 gives the treated and untreated outcomes for those rows, for two retained pathways that meet the same test, and for other rows whose untreated course explains their membership; where no study of the untreated course exists, the statement is clinical reasoning and is labeled so.

**Table S25. Natural-history notes for rows admitted on the untreated course and related rows**

| Row | With treatment or under current response | Untreated or unrelieved | Basis |
|---|---|---|---|
| Hypoglycemia treated with dextrose or glucagon (S1) | Reversed by a crew with dextrose or glucagon; outcomes under a functioning service show no time gradient | Severe hypoglycemia progresses to seizure, coma, and death | Clinical reasoning; American Diabetes Association Standards of Care [51] |
| Acute alcohol intoxication with airway compromise requiring intubation (S1) | 2,221 alcohol poisoning deaths a year in the United States in 2010 to 2012 under current response | Loss of airway protection is fatal without airway management | Deaths: [134]; untreated course: clinical reasoning, NAEMSP airway guideline [59] |
| Pediatric airway-procedure workload proxy (S1) | 30% survival in a mixed, arrest-heavy treated cohort | Progresses to hypoxic arrest | Treated cohort: [71]; untreated course: clinical reasoning |
| Device-dependent patient emergencies (retained pathway, Table S21) | Reliably managed by a crew (tracheostomy replacement, manual ventilation, device troubleshooting) | Tracheostomy obstruction, ventilator failure, and LVAD pump failure are fatal within minutes | Clinical reasoning |
| Severe foreign-body airway obstruction (retained pathway, Table S21) | 0.4% died when the obstruction was relieved before paramedic arrival | 42.4% died when the obstruction was unrelieved at paramedic arrival; neurologic outcome worsens with the stage at which the body is removed | [119,122] |
| Adult asthma/COPD encounters with assisted ventilation (S1) | Mortality 9% with noninvasive ventilation in acute exacerbations of chronic obstructive pulmonary disease | Mortality 29% with standard care in the same trial | [135] |

| Row | With treatment or under current response | Untreated or unrelieved | Basis |
|---|---|---|---|
| Gastrointestinal bleeding in hemorrhagic shock (S1) | Index mortality 1.9% for treated non-variceal upper gastrointestinal hemorrhage | Shock without hemostasis is fatal in most cases | Treated: [84]; untreated course: clinical reasoning |
| Upper airway obstruction other than foreign body (S1) | Treated epiglottitis mortality 0.89% | Complete obstruction is fatal within minutes | Treated: [106]; untreated course: clinical reasoning |
| Major burn with inhalation injury (S1) | Early airway control and burn-center care | Airway edema without airway control is fatal in most cases | Clinical reasoning |
| Ruptured ectopic pregnancy (S1) | Surgical hemorrhage control | Majority fatal in the pre-surgical era | Clinical reasoning; ACOG bulletin [100] |
| Obstetric and neonatal emergencies (S1) | Positive-pressure ventilation by the responding crew | Neonates needing positive-pressure ventilation face high mortality without it | Clinical reasoning; neonatal resuscitation guideline [89] |
